# Cross-Country Differences in Self-Rated Health: Decomposing the Influence of Depressive Symptoms

**DOI:** 10.1101/2025.10.23.25338675

**Authors:** Jacques Wels, Jane Maddock, Praveetha Patalay

## Abstract

**Background:** Despite extensive research on self-rated health (SRH), the extent to which mental health is a component part of SRH assessments and whether this relationship varies by country remains underexplored. This study quantifies the contribution of depressive symptoms to self-rated health and assesses cross-national variation in this association.

**Methods:** Using general population data from older adults (age 55 years+) in 18 countries in the Survey of Health, Ageing and Retirement in Europe (SHARE, 2011-2015), we employed four approaches: (1) country-specific pseudo R² models assessing the contribution of depressive symptoms (EURO-D) to SRH, adjusting for socio-demographics, physical health, and functional limitations; (2) partial correlation networks to visualize relationships between SRH, depressive symptoms, and physical health indicators; (3) Blinder-Oaxaca decompositions to quantify cross-country differences in SRH attributable to depressive symptoms and (4) stratification of the estimates by gender.

**Results:** Depressive symptoms explained nearly as much variance in SRH as physical health. Partial correlation networks revealed SRH as central to all health measures, and correlations with depressive symptoms ranging from 0.15 (Israel) to 0.35 (Hungary). Decomposition analyses identified three distinct patterns: Nordic countries (e.g., Denmark, Sweden) had lower depression prevalence but stronger depression-SRH associations; Eastern European countries (e.g., Estonia, Hungary) showed lower depression levels but weaker depression-SRH associations; and Mediterranean countries (e.g., Greece, Spain) exhibited higher depressive symptoms prevalence but weaker depression-SRH associations. Gender disparities in SRH were largely explained by differences in depressive symptom levels rather than differential weighting of depressive symptoms in self-assessments.

**Discussion:** Depressive symptoms contribute to self-rated health independently of physical health, confirming that mental health is integral to how individuals perceive their health. However, the extent to which this is the case varies cross-nationally highlighting that the meaning of ‘health’ is shaped by national and cultural contexts and this should be considered in any within-country or cross-national research using self-rated health measures.

**Highlights:**

- Mental health is a component as important as physical health in self-rated health.
- The contribution of depressive symptoms to SRH varies across countries
- Depressive symptoms levels partly explain country differences in self-rated health.
- Gender gaps in self-rated health are largely explained by differences in extent of depressive symptoms rather than differences in its weighting

## Background

Self-rated health (SRH) is widely used in public health research, particularly in studies utilising a comparative framework (e.g., Mak et al., 2023) and is considered as a reliable measure of overall health and a strong predictor of morbidity (Jylha, 2011; Liu et al., 2021; Mossey & Shapiro, 1982; Wu et al., 2013).

With few exceptions, SRH – also known as self-reported health or general health – is an ordinal variable capturing, on Likert scale, the way respondents rate their own health over a timeframe that could be the year prior the interview, the interview week, or the interview time. Response options are usually on a scale from very good to very poor and contain, in most cases, five options. SRH is often a key health variable in population data and is often repeated within surveys more than other health measures (Van Doorslaer & Jones, 2003). It is a convenient measure with response options that are often similar across countries, easing comparison and avoiding the need to retrospectively harmonise variables. However, little attention has been paid to what exactly constitutes self-rated health and the mental health dimension of self-rated health is underappreciated.

It has long been established that SRH is a strong predicator of mortality. Since the late 1970s, a growing corpus of studies has flourished aiming at estimating the relationship between SRH, the presence of health conditions (HC) and mortality. (Mossey & Shapiro, 1982) first examined in-depth the hypothesis that self-rated health is a predictor of mortality that is independent of "objective health status" (even though such an intuition was already present in (Singer et al., 1976) but focusing principally on the relationship between mental health, hospitalization and mortality). They carried out their analysis on a population aged 65 and over in Manitoba and found that the association between a poor SRH and risk of death is much higher than that associated with objective health status, age, sex, life satisfaction, income and whether respondents live in an urban or rural area. The study clearly provides support for considering further the way respondents view their own health as it is highly related with further health outcomes. (Idler & Benyamini, 1997) carried out a review of twenty-seven community studies between 1982 and 1996 and concluded that SRH is an independent predictor of mortality in nearly all studies. One interesting result from the review is that gender plays a key role in five of the seven studies included in the review that carried out the analyses separately for men and women: the association between SRH and mortality is much stronger for men than it is for women (Jylha et al., 1998). Though, one of the studies observed the opposite relationship (McCallum et al., 1994).

But SRH goes beyond the absence or presence of morbidities and mortality. As stated by the pioneers on the research on SRH, the variable measures ‘something more – and something less – than objective medical ratings’ (Maddox & Douglass, 1973). In that sense, health, as captured by SRH is not defined solely as the absence of diseases, especially those that could be life-threatening but is in line with the WHO definition of health where health is a ‘a state of complete physical, mental and social wellbeing and not merely the absence of disease and infirmity’ (Quesnel-Vallée, 2007). Nevertheless, SRH is constantly and positively related to objective evaluations of health status (Maddox & Douglass, 1973).

The interpretation of SRH varies across population sub-groups and countries. Several studies have found that there are constant gender differences in SRH with women’s health significantly worse than men’s. It was estimated that about 30 percent of this gap is attributable to differences in material conditions such as employment, education, or marital status (Hosseinpoor et al., 2012). However, the size of the gap varies across countries, and a significant portion remains unexplained by these observable gender-based inequalities. Cross-country differences also exist when looking at the associations between SRH and the risk of short-term mortality among older adults as SRH better predicts, for instance, older male mortality in the US (Assari, 2016) and female mortality in Israel (Benyamini et al., 2003).

Education, employment and wealth also play a key role in explaining inequalities in self-rated health (Van Doorslaer & Jones, 2003). Association between self-rated health and objective measurement of health differ by socio-economic group. For instance, looking at the same health conditions and comparing working and non-working respondents, it was shown that non-working respondents were more likely to reports a poor SRH (Butler et al., 1987). Education is also seen as a main driver of self-report health inequalities (Schuring et al., 2015) that is independent on national setting, region or gender (Subramanian et al., 2010). However, low functional health literacy is associated with higher propensities of poor self-rated health and is a higher predictor compared with education (Baker et al., 1997). Similarly, age is associated with self-rated health with poorer values observed in old age (Gunzelmann & Hinz, 2006).

Differences exist in the adverse events individuals face, how they experience their health conditions and the way they translate health conditions into self-rated health reports. Such a difference also exists when comparing nations. Cross-national comparison of subjective health measurements has been widely used showing important difference across countries. These studies underline the subjective dimension of self-report health, such as the propensity to report poor health and illness behaviours (Kunst et al., 1995). Again, population characteristics such as education, employment or age, on the one hand, and the way self-rated health is understood might explain why gaps are observed across countries. Self-rated health, particularly within a cross-national setting, exemplifies particularly well the difference between individual susceptibility to poor health and the underlying distribution (Rose, 1985), questioning what would be the good threshold to distinguish health from non-healthy respondents and whether such a threshold has any validity in an international setting.

Yet, whilst these dimensions have been explored by previous studies, one important dimension that has been often omitted is the role mental health plays in partly explaining or as a key component part of self-rated health. Poor self-rated health is constantly associated with poor mental health outcomes (Chun et al., 2015). Recently, it was observed using the Survey of Health, Ageing and Retirement in Europe (SHARE) that higher levels of depressive symptoms (measured on the EURO-D scale) are associated with a poorer self-perception of physical health and economic difficulties (Portellano-Ortiz et al., 2018). It was shown that such relationship varies with age as the relationship between mental health and SRH is stronger within younger cohorts (Spuling et al., 2015). One issue is that studies usually address mental health as a confounder of self-rated health (Lorem et al., 2017), not as a dimension of it. No study has so far examined to what extent mental health, or more specifically depressive symptoms, might be considered a component part or dimension within SRH.

To address this missing link, this study uses cross-national data from SHARE and aims to answer the following research questions:

**[RQ1]** To what extent self-rated health encompasses depressive symptoms and does this vary by country.
**[RQ2]** How do self-rated health, depressive symptoms, activities of daily living, health conditions correlate with each other, and does this vary by country?
**[RQ3]** Whether these observed differences are due to differences in country-level population composition including age, income levels, gender or education and what is the share of correspondence between self-rated health that is independent of population composition?
**[RQ4]** To what extent do the relationships explored in RQ1-RQ3 – specifically, the association between self-rated health and depressive symptoms, and its cross-national variation – differ by gender?

## Data and methods

### The Survey of Health, Ageing and Retirement in Europe (SHARE)

We have pooled three waves from the Survey of Health, Ageing and Retirement in Europe (SHARE). Wave 4 was collected in 2011, wave 5 was collected in 2013 and wave 6 was collected in 2015. The sample includes all respondents aged 55 and over in 18 countries. Sample sizes, shown in <u>table 1</u>, were as follow: 11,201 in Austria; 13,713 in Belgium; 2,130 in Croatia; 14,395 in Czech Republic; 8,272 in Denmark; 15,936 in Estonia; 12,216 in France; 9,832 in Germany; 2,595 in Hungary; 4,289 in Israel; 11,809 in Italy; 2,690 in Luxembourg; 6,018 in the Netherlands; 8,717 in Slovenia; 14,318 in Spain; 9,864 in Sweden; 8,381 in Switzerland. Data was pooled over all three waves in most countries except Croatia and Greece (wave 6 only), Hungary (wave 4) and Israel and Luxembourg (waves 5 and 6).

**Table 1.** Country descriptives statistics.

| Country | N | Available waves |  |  | SRH |  |  | EUROD |  |  | Health condition |  |  | ADL |  |  |
| --- | --- | --- | --- | --- | --- | --- | --- | --- | --- | --- | --- | --- | --- | --- | --- | --- |
|  |  | 4 | 5 | 6 | Mean | Median | SD | Mean | Median | SD | Mean | Median | SD | Mean | Median | SD |
| Austria | 11,201 | x | x | x | 3.04 | 3.00 | 1.05 | 2.04 | 2.00 | 2.02 | 0.66 | 1.00 | 0.47 | 0.11 | 0.00 | 0.32 |
| Belgium | 13,713 | x | x | x | 3.05 | 3.00 | 0.97 | 2.56 | 2.00 | 2.22 | 0.68 | 1.00 | 0.47 | 0.17 | 0.00 | 0.38 |
| Croatia | 2,130 |  |  | x | 3.38 | 3.00 | 1.17 | 2.61 | 2.00 | 2.46 | 0.68 | 1.00 | 0.47 | 0.11 | 0.00 | 0.32 |
| Czech Republic | 14,395 | x | x | x | 3.44 | 3.00 | 0.98 | 2.33 | 2.00 | 2.22 | 0.74 | 1.00 | 0.44 | 0.13 | 0.00 | 0.33 |
| Denmark | 8,272 | x | x | x | 2.57 | 2.00 | 1.16 | 1.75 | 1.00 | 1.84 | 0.61 | 1.00 | 0.49 | 0.09 | 0.00 | 0.28 |
| Estonia | 15,936 | x | x | x | 3.93 | 4.00 | 0.82 | 3.13 | 3.00 | 2.29 | 0.73 | 1.00 | 0.44 | 0.18 | 0.00 | 0.38 |
| France | 12,216 | x | x | x | 3.27 | 3.00 | 1.02 | 2.86 | 2.00 | 2.29 | 0.65 | 1.00 | 0.48 | 0.14 | 0.00 | 0.35 |
| Germany | 9,832 | x | x | x | 3.30 | 3.00 | 0.98 | 2.26 | 2.00 | 1.99 | 0.69 | 1.00 | 0.46 | 0.12 | 0.00 | 0.32 |
| Greece | 4,230 |  |  | x | 3.11 | 3.00 | 1.01 | 2.64 | 2.00 | 2.62 | 0.72 | 1.00 | 0.45 | 0.09 | 0.00 | 0.28 |
| Hungary | 2,595 | x |  |  | 3.75 | 4.00 | 1.02 | 3.17 | 3.00 | 2.64 | 0.75 | 1.00 | 0.43 | 0.14 | 0.00 | 0.34 |
| Israel | 4,289 |  | x | x | 3.20 | 3.00 | 1.16 | 2.38 | 2.00 | 2.46 | 0.70 | 1.00 | 0.46 | 0.15 | 0.00 | 0.36 |
| Italy | 11,809 | x | x | x | 3.33 | 3.00 | 1.05 | 2.90 | 2.00 | 2.57 | 0.65 | 1.00 | 0.48 | 0.13 | 0.00 | 0.33 |
| Luxembourg | 2,690 |  | x | x | 3.10 | 3.00 | 1.04 | 2.48 | 2.00 | 2.21 | 0.70 | 1.00 | 0.46 | 0.12 | 0.00 | 0.32 |
| Netherlands | 6,018 | x | x |  | 2.98 | 3.00 | 1.03 | 1.85 | 1.00 | 1.93 | 0.57 | 1.00 | 0.49 | 0.08 | 0.00 | 0.27 |
| Slovenia | 8,717 | x | x | x | 3.38 | 3.00 | 1.00 | 2.39 | 2.00 | 2.08 | 0.70 | 1.00 | 0.46 | 0.12 | 0.00 | 0.32 |
| Spain | 14,318 | x | x | x | 3.42 | 3.00 | 1.02 | 2.62 | 2.00 | 2.65 | 0.70 | 1.00 | 0.46 | 0.14 | 0.00 | 0.35 |
| Sweden | 9,864 | x | x | x | 2.78 | 3.00 | 1.14 | 1.97 | 2.00 | 1.85 | 0.62 | 1.00 | 0.49 | 0.09 | 0.00 | 0.29 |
| Switzerland | 8,381 | x | x | x | 2.72 | 3.00 | 0.95 | 1.91 | 2.00 | 1.80 | 0.54 | 1.00 | 0.50 | 0.07 | 0.00 | 0.25 |

### Self-rated health

Self-rated health (SRH) is the answer to the following question: “How is your health in general”. It contains five response options: excellent, very good, good, fair and poor. An important methodological feature of SHARE is that this question is randomly placed – either at the beginning or the end of the physical health questionnaire module – and later combined into a single variable. This randomization helps address potential biases, as research shows that there is a relatively high propensity (28 percent) for respondents to change their health self-assessment when the question is asked twice within the survey (Clarke & Ryan, 2006) and questions order might have an effect on response (particularly extreme and non-extreme values) (Crossley & Kennedy, 2002). The way the question is phrased does not specify physical health but rather target health in general. Our main analyses use SRH as a numeric variable, SRH forming a continuum from good to poor health (Manderbacka et al., 1998). However, the underlying information is inherently categorical and other methods can be used such as probit regression or interval regression (Van Doorslaer & Jones, 2003). As the interval regression was estimated to outperform other types of modelling (Van Doorslaer & Jones, 2003), we replicate the main analyses using an interval regression modelling (on point estimates) as a sensitivity check.

### Depressive symptoms

Depressive symptoms are examined using the EURO-D variable (Prince, 2002) that is homogeneously collected throughout SHARE (Guerra et al., 2015). EURO-D was originally developed to harmonize data on depression in late-life (Castro-Costa et al., 2008) and is among the most used survey measures to address depressive symptoms in older population studies (Portellano-Ortiz et al., 2018). EURO-D assesses the frequency and severity with which symptoms of depression were experienced over the past month with respondents asked to answer yes-no about how they experienced each symptom. Symptoms include depressed mood, pessimism, suicidality, guilt, sleep, interest, irritability, appetite, fatigue, concentration, enjoyment, and tearfulness. Scores range from zero to 12, higher scores indicating higher levels of depressive symptoms (Gana et al., 2023). Cross-national variations in depression prevalence across country exist but they are not due to differences in the nature or validity of reporting depressive disorders (Simon et al., 2002).

### Control variables

We derived several sets of harmonized control variables from SHARE.

Health condition reflect whether the respondents report at least one chronic health condition rated by a medical doctor. Medical conditions include heart attack; high blood pressure or hypertension; high blood cholesterol; stroke or cerebral vascular disease; diabetes or high blood sugar; chronic lung disease; arthritis, including osteoarthritis, or rheumatism; cancer or malignant tumour; stomach or duodenal ulcer, peptic ulcer; Parkinson disease; cataracts; hip fracture or femoral fracture. The variable is dichotomised distinguishing those reporting at least one condition (=1) from those reporting no listed condition (=0). We select this variable to address physical health issues because they reflect both the existence of specific health conditions that might translate into poorer self-rated health scores and the experience of daily living impairment that might also affect SRH.

We additionally include information on Activities of Daily Living (ADL), i.e., the difficulty performing activities of daily living to maintain one’s person (Espinoza et al., 2018). ADL scores information on tasks of dressing, bathing/showering, eating, cutting up food, walking across a room and getting in or out of bed with values from zero to five, five indicating the highest level of difficulty (Hajek & König, 2016). The variable is coded as binary distinguishing those who report at least one issue with ADL. ADL is distinguished from health conditions in the sense that all conditions do not translate into ADL impairment and ADL could exist independently of health conditions (Espinoza et al., 2018) and is a strong confounder of wellbeing indicators within elderly populations (Barile et al., 2012).

Demographic information includes gender (male is the reference category), age-group (55 to 65 or above) and marital status (married and living together with spouse or registered partnership (reference), not married or divorced, and widowed). Information on respondents’ socio-economic status (SES) include household incomes equivalized to reflect household composition (log) (Anyaegbu, 2010), the highest level of education (based on the International Standard Classification of Education – ISCED classification and distinguishing respondent with no formal education or ISCED 1, respondents reporting ISCED 5 to 6 (reference) and those reporting another type of education not included in the ISCED nomenclature) and housing tenure (owner (reference), not owner).

### Methods

#### Explained variance

To estimate the variance in SRH attributed to depressive symptoms, we estimated a series of weighted linear regression models separately for each country. SRH was the dependent variable, and depressive symptoms were measured using the EURO-D scale. We used a stepwise modelling strategy to assess how the explanatory power of depressive symptoms changed with the inclusion of different sets of variables. Model 1 included socio-demographic and socio-economic factors such as age, gender, education, employment status, income, and household characteristics. Model 2 added physical health indicators, including the presence of chronic conditions and limitations in activities of daily living (ADL). Model 3 further included the EURO-D score to assess its additional contribution to explaining SRH. This approach allowed us to isolate the role of depressive symptoms after accounting for other relevant factors and to compare their influence across countries. The models controlled for the wave of data collection when several waves were available by country and weighted using cross-sectional survey weights. To estimate the proportion of variance explained by each model, we computed a weighted pseudo R-squared based on the deviance of the fitted and null models. To assess the uncertainty around R-squared, we used non-parametric bootstrapping. For each country and each model, we generated 100 bootstrap samples by resampling the data with replacement. A weighted linear regression model was fitted to each resample, and the pseudo R-squared value was extracted. We then calculated the 95% confidence intervals using the percentile method based on the bootstrap distribution. This method provides a robust and assumption-free way to estimate confidence intervals for R-squared, which does not have a standard analytical distribution (Wagstaff et al., 2009). By comparing the pseudo R-squared values and their confidence intervals across the three models, we were able to quantify how much depressive symptoms contributed to explaining SRH in each country.

#### Partial correlations networks

To examine the associations between Self-Rated Health (SRH, numeric), Activities of Daily Living (ADL), the presence of chronic health conditions (binary), and the EURO-D scale (treated as binary) we used partial correlation networks to generate separate plots for each selected country. In each network plot, nodes represent observed variables, while edges represent regularized partial correlations between pairs of variables, controlling for all others (Epskamp et al., 2018). To ensure sparsity, we applied Lasso regularization using the graphical Lasso method. The optimal tuning parameter was selected using the Extended Bayesian Information Criterion (EBIC), as proposed by Epskamp and Fried (2016), which balances model fit and complexity to yield a balanced network structure. The network layouts were visualized using a Fruchterman-Reingold algorithm, adapted for weighted networks. This layout places nodes with stronger connections closer together, enhancing interpretability. In the final figures, edge thickness reflects the magnitude of the partial correlation, with thicker edges indicating stronger conditional associations. 95% confidence intervals (CI) were approximated using Fisher’s z-transformation and are shown in the supplementary files.

#### Blinder-Oaxaca decomposition

To assess how much of the cross-country variation in self-rated health can be attributed to differences in population composition versus differences in the social and health contexts that shape how these characteristics influence health, we employed a Blinder-Oaxaca decomposition technique. Decomposition techniques are widely used in the social sciences (Oaxaca & Sierminska, 2023) and Blinder-Oaxaca types of decomposition are popular in economics. Although not often used to address public health issues, the advantages of this method to address health inequalities were recently emphasised (Rahimi & Hashemi Nazari, 2021). In this study, the Blinder-Oaxaca decomposition model was employed to analyse the differences in SRH scores across countries. Each of the 18 countries were compared to a reference category that is the sum of all countries weighted for each country to have identical sample size based on the original population weights. In other words, we compared each country (i) to a reference group (m) that corresponds to the mean self-rated health observed within the 18 selected countries (formula [a]). By doing so, we generated estimates that do not compare countries to a single reference country which would make the estimates highly dependent on the value of the reference group.

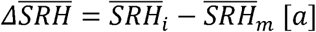

The Blinder Oaxaca decomposition estimated the mean difference in the outcome variable between the two groups, i.e., the country of interest versus the reference groups. The model decomposed the observed mean difference in SRH into two main components. The *explained component* is the part of the outcome difference due to differences in the average levels of depressive symptoms (EURO-D) between the two groups, weighted by the coefficients from the reference group as can be seen in formula [b] where ‘m’ corresponds to the reference group, ‘i’ is the group of interest and ‘I’ marks the set of control variables. It represents the portion of the difference in self-rated health that can be attributed to differences in observable characteristics, such as depression or incomes, between the countries. If the explained component for depression is positive, it suggests that part of the difference in self-rated health between the reference group and the other countries can be explained by differences in the levels of depression between these countries.

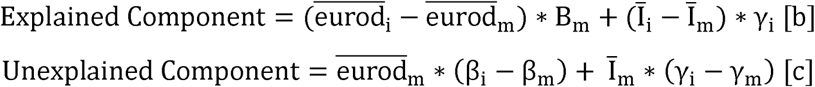

By contrast, the *unexplained component* is the part of the outcome difference that is due to differences in the coefficients of EURO-D and the other control variables between the two groups, as can be seen in formula [c]. The unexplained component captures the portion of the difference in self-rated health that cannot be explained by observable characteristics. This includes differences in coefficients (how depression associates self-rated health) and other unmeasured factors. If the unexplained component is significant for depression, it suggests that the association between depression and SRH is different between the country of interest and the reference group. This could be due to differences in how depression affects people’s health in different cultural or healthcare contexts, or due to the influence of unobserved factors that differ between the countries. The total mean difference is the sum of the explained and unexplained components.

Due to the nonlinear nature of interval regression models, it was not possible to directly compute the explained and unexplained components of group differences by covariate, as in linear Blinder-Oaxaca decompositions. In nonlinear settings, the contributions of individual variables are not separable in an additive way. Therefore, our main analyses rely on linear models, which allow for a full decomposition, while interval regression were used in sensitivity analyses to examine overall group differences only.

### Weights

We used complete case analysis and included cross-sectional weights provided by SHARE.

### Stratification

We first conduct analysis in the full sample and subsequently all analyses were stratified by gender to allow examination of gender differences.

### Sensitivity analysis

To increase statistical power, we conducted the main analyses on a pooled sample that included all available waves for each country, applying population weights and adjusting for wave fixed effects. As a sensitivity check, we replicated the analyses using a restricted sample that considered only respondents’ participation in the last available wave. No significant differences were observed between the two approaches. The main model included SRH as a numeric variable. To address normal distributiveness issues and the ordinal nature of SRH, we also replicated the main analyses using an interval regression modelling.

### Software

Data cleaning, partial correlations and network analyses were made using the R software. Blinder-Oaxaca models were calculated using Stata v.17 and specifically, the *oaxaca* and *nldecompose* commands.

## Results

### Descriptive results

Table 1 presents descriptive statistics for self-rated health (SRH), depressive symptoms (EURO-D), health conditions, and limitations in activities of daily living (ADL) across European countries. On average, self-rated health was highest in Denmark (mean = 2.57) and Switzerland (2.72) and lowest in Estonia (3.93) and Hungary (3.75). Depression scores varied considerably, with the lowest levels in Denmark (1.75) and Switzerland (1.91) and the highest in Estonia (3.13) and Hungary (3.17). The prevalence of health conditions was generally high, particularly in the Czech Republic (0.74) and Hungary (0.75), while Switzerland (0.54) and the Netherlands (0.57) rated the lowest means. ADL limitations were least common in Switzerland (0.07) and the Netherlands (0.08) and most frequent in Estonia (0.18) and Belgium (0.17).

### Explained variance

**<u>Figure 1</u>** exhibits the explanatory power of three sequentially adjusted regression models for self-rated health across the 18 countries. The full results are shown supplementary file S.1. The left panel displays pseudo R² values with 95% confidence intervals for: (1) a baseline socio-economic model, (2) an intermediate model incorporating physical health measures (chronic conditions and functional limitations), and (3) a full model additionally including depressive symptoms. Countries are ordered by the R² values of the full model. The right panel illustrates the marginal gains in explanatory power achieved by adding physical health indicators and subsequently depressive symptoms, with a dashed zero-line reference indicating no improvement. A first observation is that in all countries except Greece and Israel, both health conditions/ADL and EURO-D add explanatory power to the model. For instance, in Switzerland, the model that includes only socio-demographic characteristics explains 11.7 percent of the variance. When health conditions and ADL are added, the explained variance increases to 19.6 percent—an improvement of about 8 percentage points. Adding depressive symptoms (EURO-D) further increases the explained variance to 27.5 percent—another gain of approximately 8 percentage points. Another observation is that there appears to be no clear correlation between the marginal gains from including health conditions and ADL and those from including depressive symptoms. In other words, physical health and depressive symptoms variables are not interchangeable.

We replicated the analyses using an interval regression and a linear model stratified by gender, with results in <u>Supplementary Files S.2 and S.3</u>.

**Figure 1.**
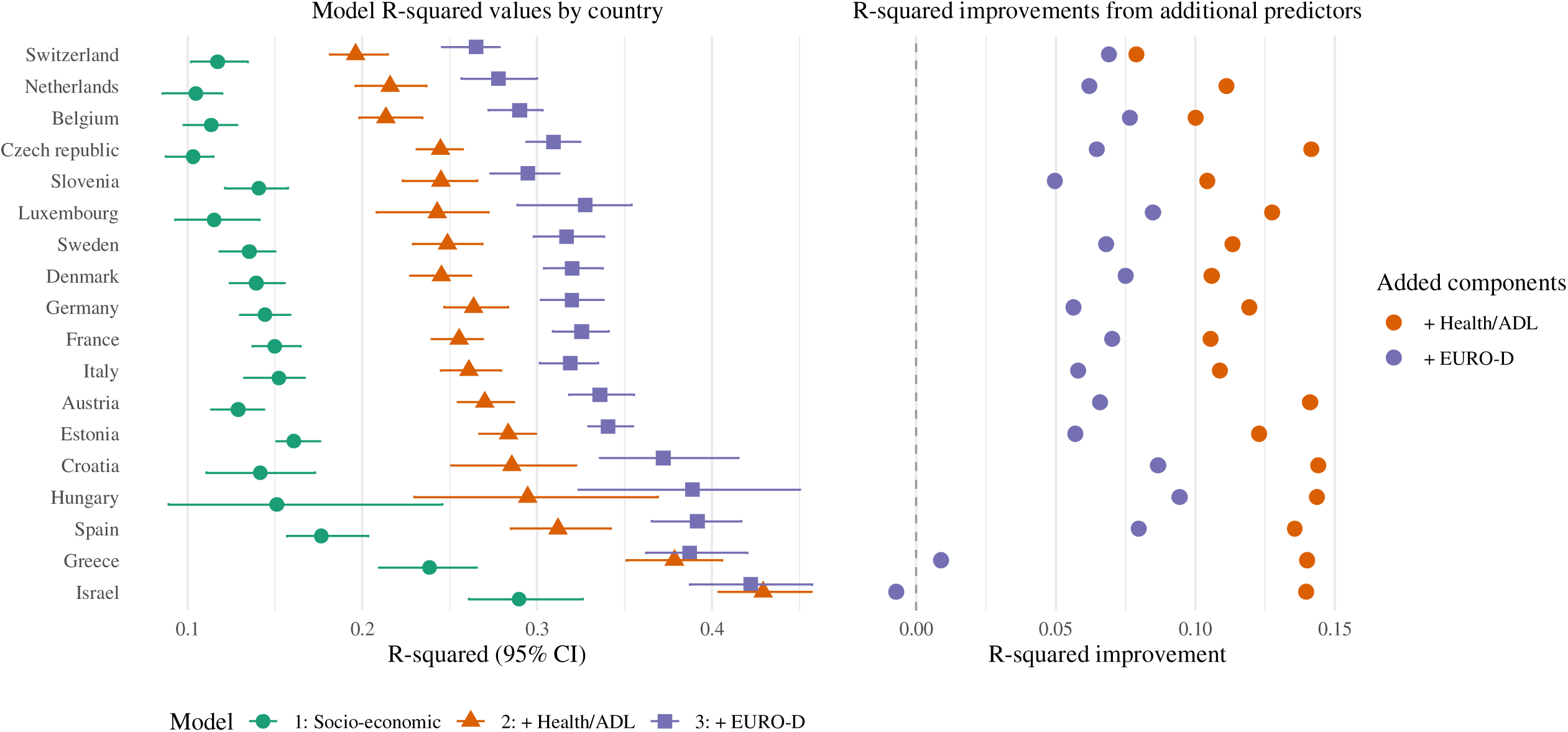
Country variations in Pseudo R-squared explained by socio-economic variables, chronic conditions (health) and activity of daily living (ADL) and depressive symptoms (EUROD-D) (linear model)

### Partial correlations networks

<u>Figure 2</u> shows the partial correlation networks by country. Full estimates and 95%CI are shown in supplementary file S.5. The graphs show the relative centrality of SRH among the other health-related variables. In all countries, self-rated health is independently associated with the presence of a health condition, ADL impairment and depressive symptoms. However, the nature of such relationships vary from one country to another. Looking at depressive symptoms (EURO-D), the partial correlation ranges from 0.15 (95%CI: 0.12; 0.18) in Israel to 0.35 (95%CI: 0.31; 0.38) in Hungary. What can also be observed is that each variable is independently correlated with SRH. The correlation estimates across EURO-D, ADL and COND are lower than their correlations with SRH. For instance, we observe that the partial correlation between EURO-D and the presence of a health condition is near zero in most countries. There is no correlation in Austria, Czech Republic, Denmark, Estonia, Germany, Greece, Luxembourg, the Netherlands, Slovenia and Spain. Croatia exhibits the highest degree of correlation with a coefficient of 0.8 (95%CI: 0.03; 0.12). Finally, the correlation between ADL and SRH is higher than the correlation observed for the presence of at least one health condition. In Denmark, for instance, the partial correlation reaches 0.49 (95%CI: 0.47: 0.50).

**Figure 2.**
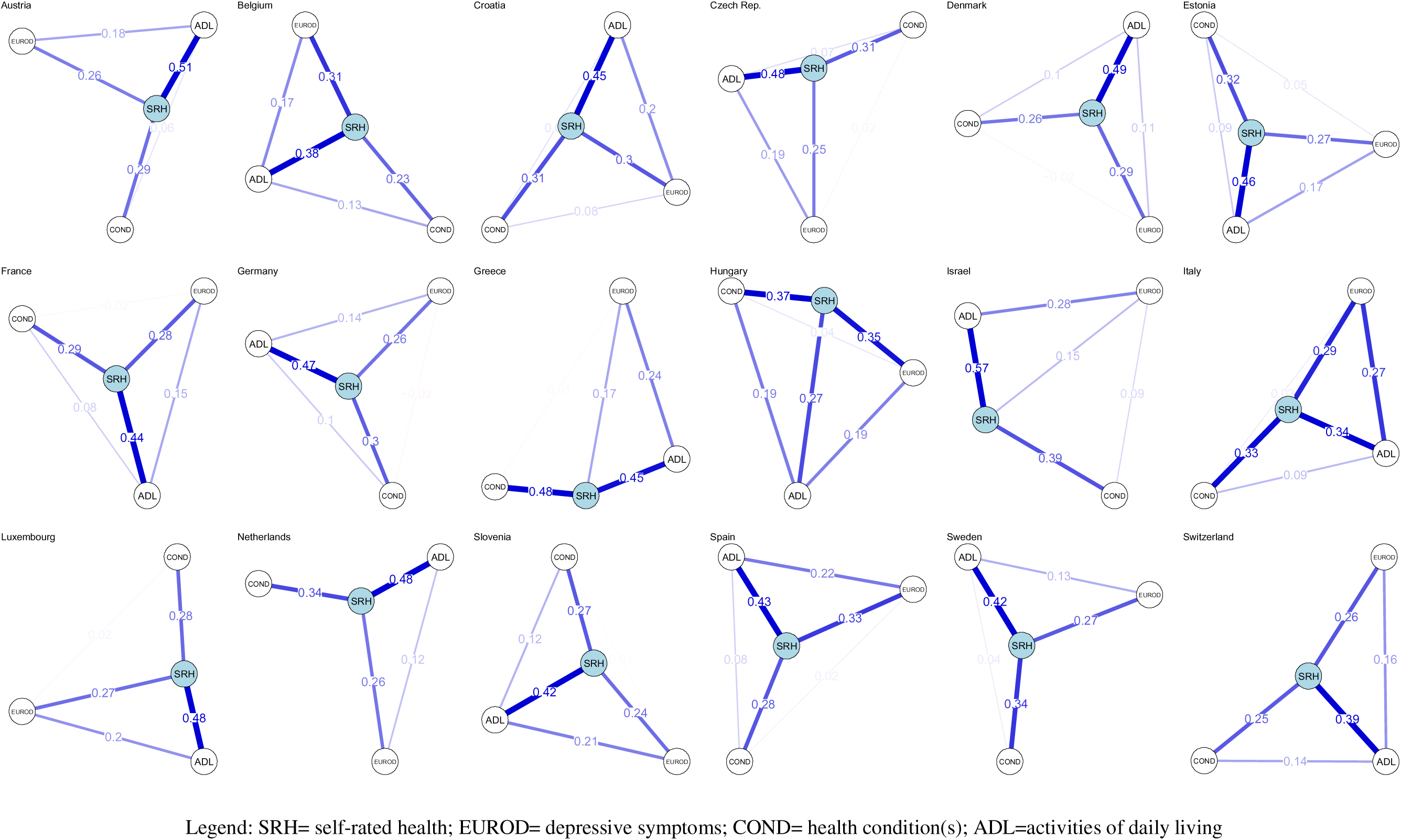
Partial correlations networks including depressive symptoms (EURO-D), activity of daily living (ADL), Chronic condition and self-rated health (SRH), by country. Complete cases.

### Blinder Oaxaca decomposition

Main estimates from the Binder Oaxaca decomposition are shown in <u>figure 3</u> whilst full results are in <u>supplementary files S.6</u>.

**Figure 3.**
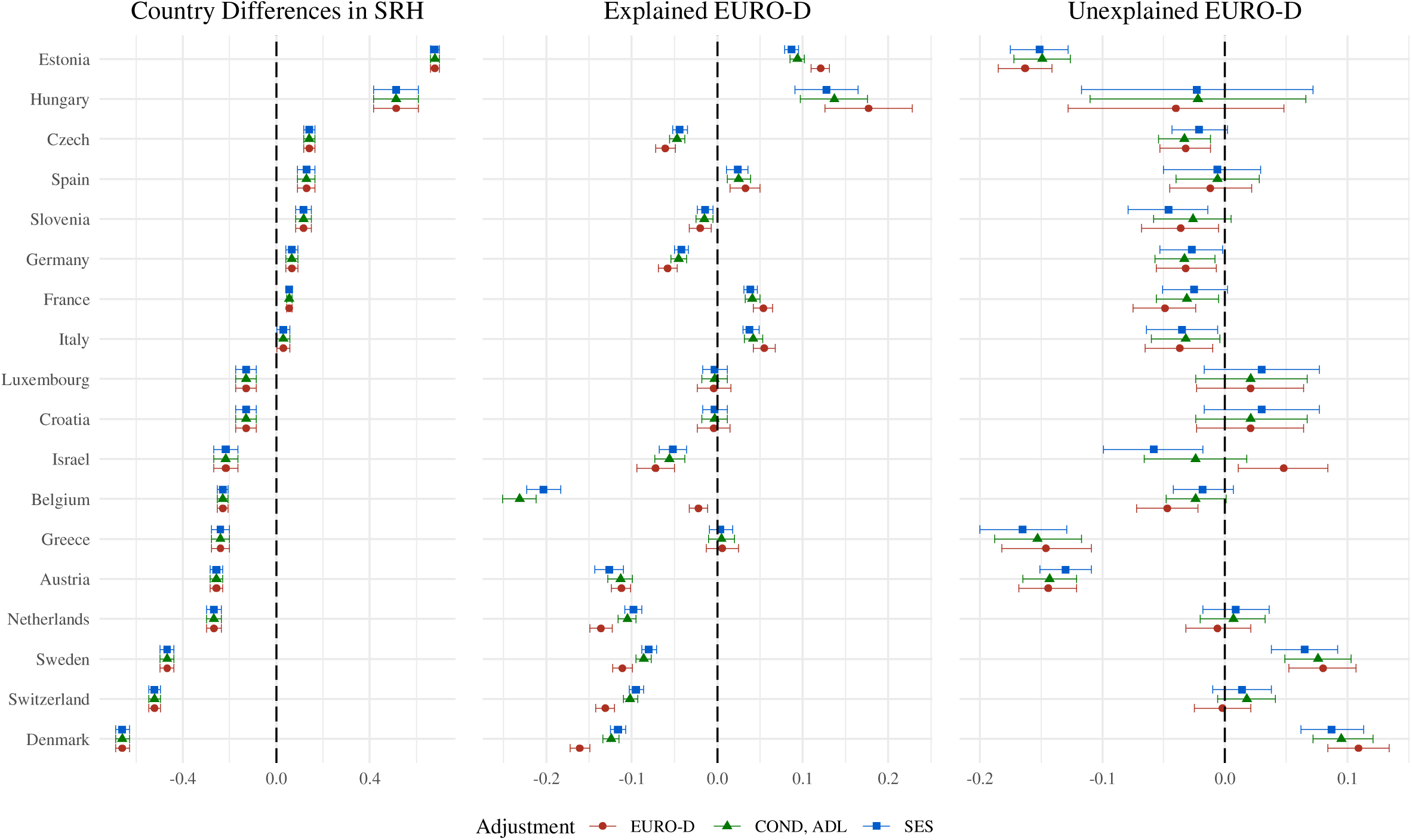
Country differences in self-rated health (SRH) decomposed into total difference (left), the part explained by depressive symptoms (EUROD-D) levels (centre), and the part explained by differences in the SRH-depressive symptoms (EURO-D) association (right).

Looking at <u>figure 3</u>, we observe that lower depression prevalence contributes to their SRH advantage in Nordic countries, as shown by negative explained components (Denmark: - 0.161 [95%CI: −0.172 to −0.149]; Sweden: −0.111 [95%CI: −0.122 to −0.099]). However, the positive EURO-D unexplained components (Denmark: 0.109 [95%CI: 0.084 to 0.134]; Sweden: 0.080 [95%CI: 0.052 to 0.107]) indicate that the relationship between depressive symptoms and SRH is actually stronger in these countries than in the reference population. This means that while Scandinavians benefit from lower overall depression rates, each unit increase in depressive symptoms has a larger negative impact on their health perceptions compared to other countries.

Eastern European countries show a different configuration. Estonia’s positive EURO-D explained component (0.121 [95%CI: 0.110 to 0.131]) confirms that higher depression prevalence contributes to poorer SRH, while its negative unexplained component (−0.163 [95%CI: −0.185 to −0.141]) reveals that the depression-SRH association is weaker than in the reference population. This suggests that although depression is more common in Estonia, its impact on health perceptions is somewhat attenuated compared to other countries. Hungary presents a similar pattern with depression prevalence playing a major role (explained: 0.177 [95%CI: 0.126 to 0.228]) but the depression-SRH relationship being slightly weaker than reference (unexplained: −0.040 [95%CI: −0.128 to 0.048]). These findings may indicate that in Eastern European contexts, other health challenges partially overshadow the impact of depressive symptoms on overall health evaluations.

Mediterranean countries demonstrate yet another distinct pattern. Greece shows minimal contribution from depression prevalence differences (explained: 0.006 [95%CI: −0.013 to 0.025]) but a substantial negative EURO-D unexplained component (−0.146 [95%CI: −0.182 to −0.109]), indicating that the association between depressive symptoms and SRH is weaker than in the reference population. Spain presents a similar but milder version of this pattern. These results suggest that while depression levels in Southern Europe may not differ dramatically from the reference, their impact on health perceptions is less pronounced.

The stability of these EURO-D specific patterns across adjustment models strengthens their interpretation. Switzerland consistently shows near-zero unexplained components, indicating its depression-SRH relationship closely matches the reference population once prevalence differences are accounted for. The Netherlands demonstrates small unexplained components across adjustments, suggesting that while depression prevalence matters for SRH differences, the strength of its association with health perceptions aligns with the reference.

<u>Figure 4</u> displays country variations in how depressive symptoms (EURO-D) influence self-rated health (SRH), decomposed into explained (observed EURO-D differences) and unexplained (divergent associations) components. Three distinct patterns emerge. Countries including Austria, Belgium and Germany show both significantly fewer depressive symptoms (negative explained component) and a negative depression-SRH association (negative unexplained component). This indicates their populations experience better mental health while simultaneously discounting depressive symptoms when evaluating overall health. Countries such as Estonia and Hungary present higher depressive symptoms (positive explained component) coupled with a negative depression-SRH association (negative unexplained component), revealing that despite greater symptom prevalence, depressive symptoms carry less weight in health assessments. Countries including Denmark, Sweden or Switzerland demonstrate fewer depressive symptoms (negative explained component) but a positive depression-SRH association (positive unexplained component), suggesting that while depressive symptoms are generally better, residual symptoms disproportionately affect health ratings. The empty quadrant where both components would be positive confirms no country suffers from the dual disadvantage of both elevated depressive symptoms and their amplified incorporation into health evaluations.

**Figure 4.**
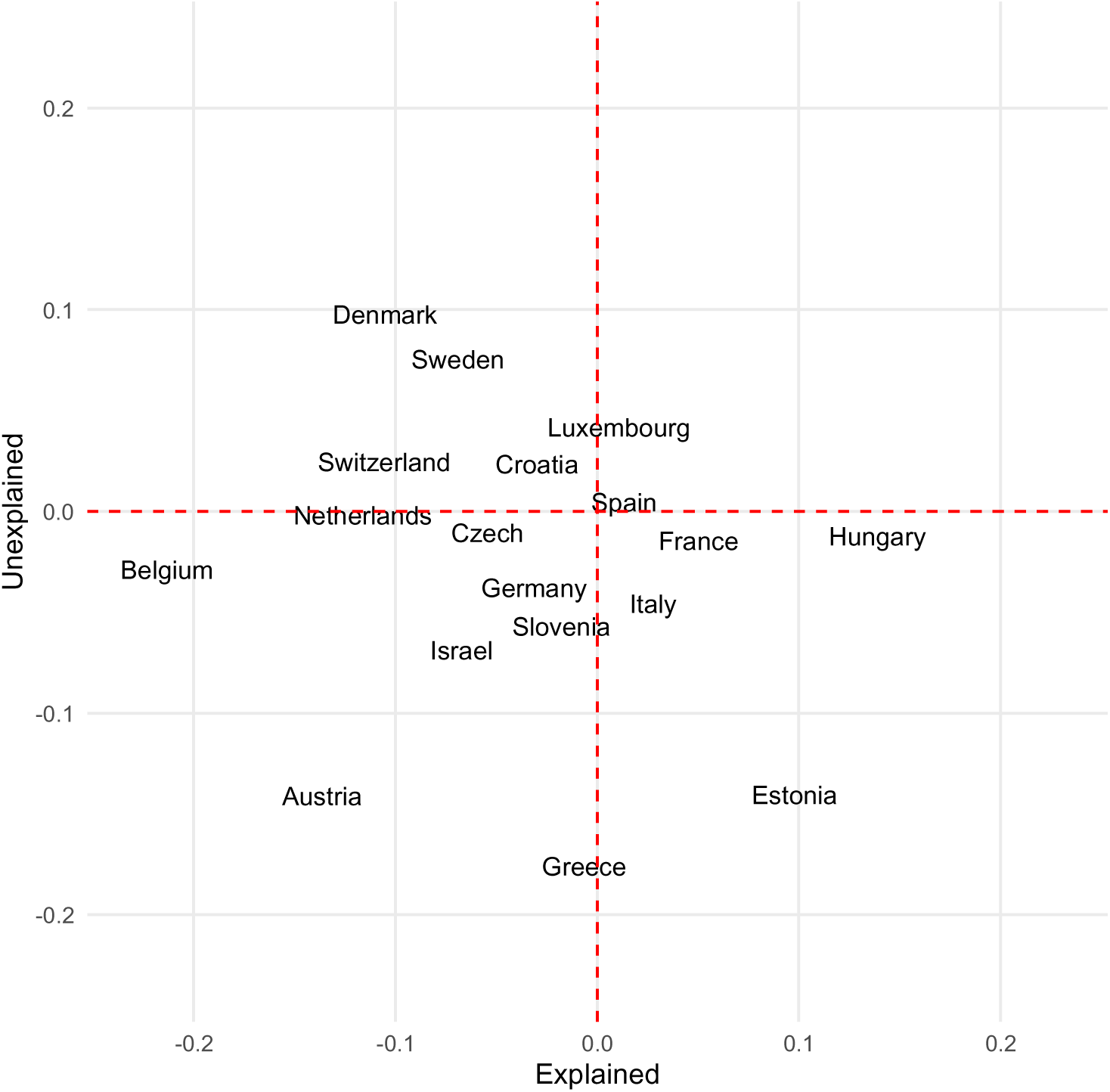
Country variations in self-rated health explained by depressive symptoms (EUROD-D) levels (x-axis), and by differences in the SRH-depressive symptoms (EURO-D) association (y-axis).

### Stratification by gender

We repeated the fully adjusted analyses with stratification by gender.

Pseudo R-square and partial correlations stratification by gender are shown in <u>supplementary files S.1.3 and S.2.4</u>, respectively. No differences are observed across genders when looking at R-square analyses. Partial correlations reveal minimal gender differences in the strength of partial correlations. While some country-specific variations were observed, the overall pattern was consistent between men and women, suggesting that the underlying associations among these health measures are largely gender-invariant.

Blinder-Oaxaca estimates, including the main difference and the explained and unexplained EURO-D components, are shown in <u>Figure 5</u>. The main results are in <u>Supplementary File S.3.4</u>. What can be observed is that males tend to report poorer self-rated health (i.e., higher difference scores) in several countries, including Estonia, the Czech Republic, Germany, Israel, Austria, and Denmark. However, most of this difference is explained by country-specific EURO-D scores, indicating that the higher prevalence of depressive symptoms among male respondents translates into poorer self-rated health. We observe little difference in the unexplained EURO-D component, which confirms the presence of country-specific differences that apply to both genders.

**Figure 5.**
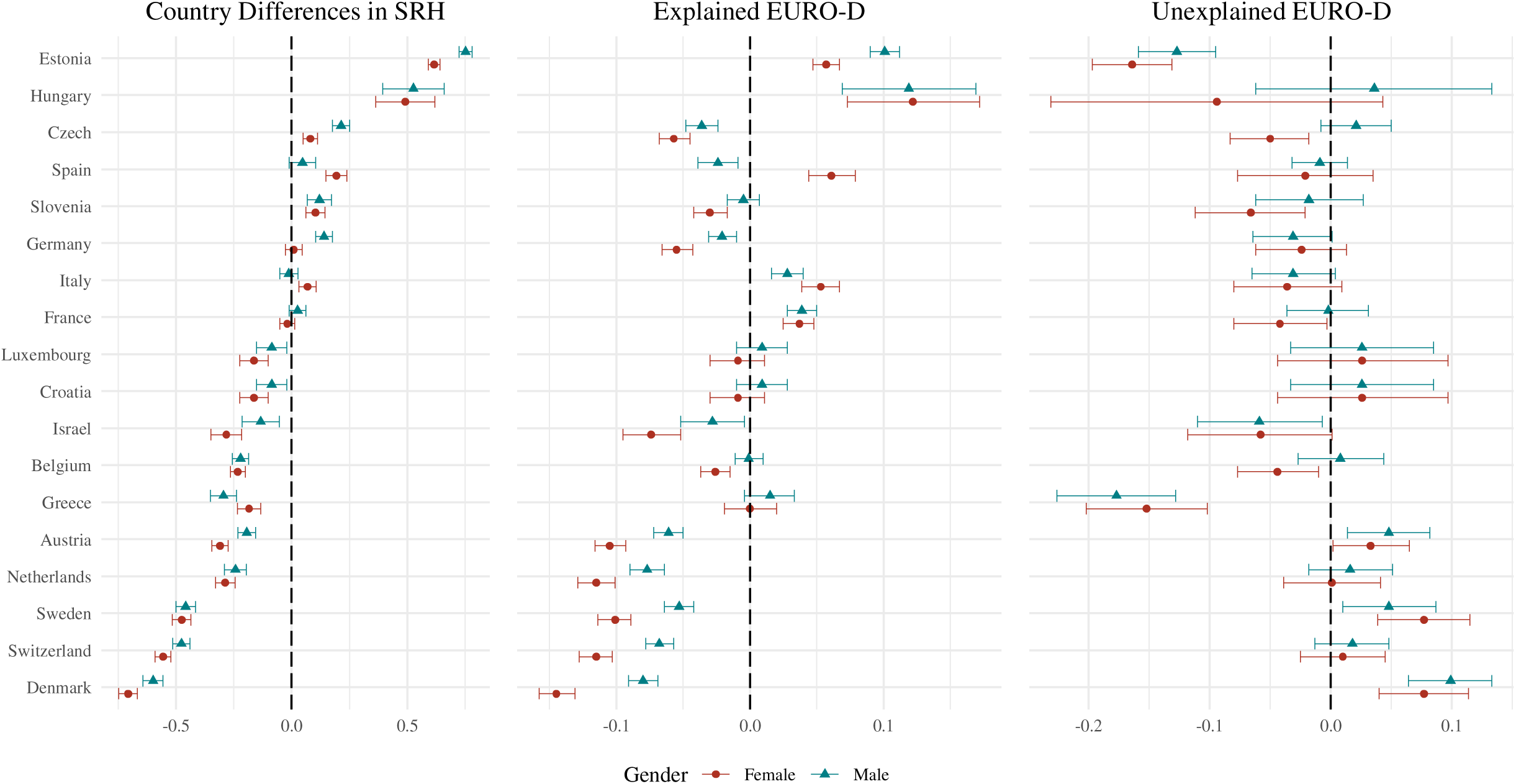
Country differences in self-rated health (SRH) decomposed into total difference (left), the part explained by depressive symptoms (EUROD-D) levels (centre), and the part explained by differences in the SRH-depressive symptoms (EURO-D) association (right), by gender

### Sensitivity analyses

We ran the main decomposition model using a sample restricted to respondents’ answers from the last available wave, in order to avoid duplication of respondents as in the pooled analyses. <u>Supplementary File S.3.6.</u> compares the main differences, as well as the explained and unexplained EURO-D differences, between the pooled dataset and the restricted dataset. No major differences are observed across estimates, except for the Austrian EURO-D unexplained component, which is −0.130 (95% CI: −0.151; −0.109) in the pooled model and 0.043 (95% CI: 0.007; 0.080) in the restricted dataset. Finally, we replicated the main model using interval regressions and plotted the main country differences – as shown in <u>Supplementary File S.3.7.</u> – with results very similar to those of the linear model, indicating that the linear Blinder-Oaxaca decomposition is reliable.

## Discussion

To our knowledge, this is the first study to systematically examine whether self-rated health incorporates depressive symptoms and whether the strength of this relationship varies across countries. Drawing on data from the Survey of Health, Ageing and Retirement in Europe (SHARE) and multiple analytical approaches, this study provides new evidence on how mental health shapes subjective health assessments in later life.

Our findings make three key contributions. First, depressive symptoms explain nearly as much variation in self-rated health as physical health conditions and functional limitations, confirming that mental health is a central element of how individuals evaluate their overall health. Second, the association between depressive symptoms and self-rated remains robust even after controlling for physical health, suggesting that mental health contributes independently to how individuals perceive and report their health status. Third, decomposition analyses reveal clear cross-national variation in the strength of the depressive symptom–self-rated health association.

We identify three empirical patterns among European countries. Countries such as Sweden and Denmark show fewer depressive symptoms but a stronger-than-reference association with self-rated health, suggesting these populations that experience better mental health yet place greater emphasis on it when rating their overall health. Countries including Germany and Belgium exhibit fewer depressive symptoms and a weaker symptom–self-rated link, indicating that good mental health is less integrated into subjective health evaluations in these countries. Countries such as Estonia and France, have higher depressive symptom levels but a weaker association with self-rated, implying that elevated symptom burdens are under-reflected in health assessments. The theoretically possible fourth type pattern, characterized by both more depressive symptoms and a stronger association with SRH, was not observed, suggesting that no population in our sample both experiences high symptom prevalence and strongly weights depressive symptoms in their health evaluations.

We also find that gender differences in SRH are largely explained by differences in depressive symptom levels rather than by differences in how depressive symptoms are integrated into self-rated health. Men and women appear to weigh depressive symptoms similarly when assessing their health, though average symptom levels differ across countries.

Several limitations should be acknowledged. First, the analyses are limited to respondents aged 55 and older in SHARE. Differences in self-rated health are known to exist across surveys, with self-rated health typically rated more favourably in SHARE compared with other datasets such as the European Social Survey or EU-SILC (Croezen et al., 2016). Similar cross-national differences would likely appear elsewhere, though their magnitude may vary due to population characteristics such as age (Pinquart, 2001). Second, to maintain model parsimony, we included a restricted set of confounders. Additional individual- and contextual variables, such as healthcare access, policy environments, or awareness of mental health, might influence both depressive symptoms and their role in self-rated. Third, the Blinder– Oaxaca decomposition was originally designed for linear outcomes (Blinder, 1973; Jann, 2008). Although non-linear extensions exist (Sinning et al., 2008), packages providing detailed decomposition outputs for interval regressions remain limited. Nonetheless, sensitivity analyses suggest that treating self-rated as a continuous variable yields consistent results, in line with existing research practice.

Our findings align with earlier descriptive evidence of cross-European gradients in depressive symptoms (Ploubidis & Grundy, 2009), with better mental health in Scandinavia, intermediate scores in Western Europe, and poorer outcomes in Southern Europe – patterns that reflect broader socioeconomic differences and heterogeneity in GDP per capita (Jorm & Mulder, 2021). Yet, previous studies have not directly examined how mental health translates into broader health self-evaluations. By isolating the unexplained component of self-rated health variation, our analysis highlights that part of the cross-national differences in self-rated health reflect not only compositional differences in depressive symptom levels but also deeper cultural and contextual factors influencing how health is understood and reported.

## Conclusion

Self-rated health (SRH) can be conceptualised in line with the World Health Organization’s definition of health as encompassing physical, mental, and social wellbeing rather than merely the absence of disease (Quesnel-Vallée, 2007). Our findings provide empirical support for this broader conception: depressive symptoms contribute to self-rated health independently of physical health, confirming that mental health is integral to how individuals perceive their health. However, the strength of this relationship is not uniform across Europe, indicating that the meaning of “health” varies across cultural and national contexts.

These cross-national differences may be rooted in variations in cultural perceptions of mental illness (Altweck et al., 2015; Bass et al., 2007), in the attitudes of medical professionals (Stefanovics et al., 2016), or in broader differences in health literacy. Comparative work on health literacy has documented substantial cross-country variation in individuals’ understanding and use health information (Sørensen et al., 2015), and ecological correlations have been observed between national self-rated health levels and health literacy scores (Portela et al., 2020). Yet, relatively little is known about mental health literacy specifically (Kutcher et al., 2016; O’Connor & Casey, 2015; Tay et al., 2018), highlighting the need for improved data to explore whether self-rated health reflects differences in mental health literacy at individual and national levels.

While perfect conceptual comparability across countries may be unattainable (Sartori, 1991), this should not preclude meaningful international comparison. Our findings offer a note of caution for researchers using self-rated health in cross-national analyses: self-rated health remains a valuable and widely used measure of general health status, but it should not be interpreted as a purely objective or universally consistent indicator in terms of it’s meaning. Because depressive symptoms significantly shape self-rated health – independently of physical health and to varying degrees across contexts – cross-national comparisons of SRH risk conflating differences in health status with differences in the cultural weighting of mental wellbeing. Accounting for mental health or explicitly modelling its contribution to SRH can help disentangle these effects. Ultimately, self-rated health should be understood not only as a reflection of health outcomes but also as a socially and culturally embedded perception of health.

## Data availability

SHARE data are available at https://share-eric.eu/.

## Authors contribution

Conceptualization: JW, PP; Data Curation: JW; Formal Analysis: JW; Funding Acquisition: NA; Investigation: JW; Methodology: JW, JM, PP; Project Administration: JW; Resources: JW; Software: JW; Supervision: JW, JM, PP; Validation: JM; Visualization: JW; Writing – Original Draft Preparation: JW; Writing – Review & Editing: JM, PP

## Supplementary files

Supplementary file S.1. Pseudo R-square analysis

Supplementary file S.1.1. Pseudo R-square comparison across countries (linear regression)
Supplementary file S.1.2. Pseudo R-square comparison across countries (interval regression)
Supplementary file S.1.3. Pseudo R-square comparison across countries (linear regression), stratification by gender

Supplementary file S.2. Partial correlations

Supplementary file S.2.1. Partial correlations for the full population, including 95%CI
Supplementary file S.2.2. Partial correlations by gender, including 95%CI
Supplementary file S.2.3. Partial correlations plot (male only)
Supplementary file S.2.4. Partial correlations plot (female only)

Supplementary file S.3. Blinder Oaxaca decomposition

Supplementary file S.3.1. Blinder Oaxaca Linear results
Supplementary file S.3.2. Explained and unexplained ADL and conditions components
Supplementary file S.3.4. Blinder-Oaxaca decomposition by gender, main estimates
Supplementary file S.3.5. Explained and unexplained ADL and health condition components, stratification by gender
Supplementary file S.3.6. Comparing pooled and unique ID models
Supplementary file S.3.7. Main differences within the linear and interval decomposition models

