## Supplementary file for "Cross-Country Differences in Self-Rated Health: Decomposing the Influence of Depressive Symptoms"

Supplementary files

### Supplementary file S.1. Pseudo R-square analysis

#### Supplementary file S.1.1. Pseudo R-square comparison across countries (linear regression)

|  | Socio-economic | | |  | + Health/ADL | | |  | +EURO-D | | |
| --- | --- | --- | --- | --- | --- | --- | --- | --- | --- | --- | --- |
| Country | R_squared | CI_lower | CI_upper |  | R_squared | CI_lower | CI_upper |  | R_squared | CI_lower | CI_upper |
| Austria | 0.129 | 0.114 | 0.144 |  | 0.270 | 0.255 | 0.287 |  | 0.336 | 0.318 | 0.355 |
| Belgium | 0.113 | 0.098 | 0.129 |  | 0.214 | 0.198 | 0.234 |  | 0.290 | 0.272 | 0.303 |
| Switzerland | 0.117 | 0.102 | 0.135 |  | 0.196 | 0.181 | 0.215 |  | 0.265 | 0.245 | 0.278 |
| Czech Republic | 0.103 | 0.087 | 0.115 |  | 0.245 | 0.231 | 0.258 |  | 0.309 | 0.294 | 0.325 |
| Germany | 0.144 | 0.130 | 0.159 |  | 0.264 | 0.247 | 0.283 |  | 0.320 | 0.302 | 0.338 |
| Denmark | 0.139 | 0.124 | 0.156 |  | 0.245 | 0.227 | 0.262 |  | 0.320 | 0.304 | 0.338 |
| Estonia | 0.161 | 0.151 | 0.176 |  | 0.284 | 0.267 | 0.299 |  | 0.341 | 0.329 | 0.355 |
| Spain | 0.176 | 0.157 | 0.203 |  | 0.312 | 0.285 | 0.342 |  | 0.392 | 0.365 | 0.417 |
| France | 0.150 | 0.137 | 0.165 |  | 0.255 | 0.240 | 0.269 |  | 0.325 | 0.309 | 0.341 |
| Greece | 0.238 | 0.210 | 0.265 |  | 0.378 | 0.351 | 0.406 |  | 0.387 | 0.362 | 0.420 |
| Croatia | 0.142 | 0.111 | 0.173 |  | 0.286 | 0.251 | 0.322 |  | 0.372 | 0.336 | 0.415 |
| Hungary | 0.151 | 0.089 | 0.246 |  | 0.295 | 0.230 | 0.369 |  | 0.389 | 0.324 | 0.450 |
| Israel | 0.290 | 0.261 | 0.326 |  | 0.429 | 0.404 | 0.457 |  | 0.422 | 0.387 | 0.457 |
| Italy | 0.152 | 0.133 | 0.167 |  | 0.261 | 0.245 | 0.279 |  | 0.319 | 0.301 | 0.335 |
| Luxembourg | 0.115 | 0.093 | 0.141 |  | 0.243 | 0.208 | 0.272 |  | 0.328 | 0.289 | 0.354 |
| Netherlands | 0.105 | 0.086 | 0.120 |  | 0.216 | 0.196 | 0.237 |  | 0.278 | 0.257 | 0.300 |
| Sweden | 0.135 | 0.118 | 0.150 |  | 0.249 | 0.229 | 0.269 |  | 0.317 | 0.298 | 0.338 |
| Slovenia | 0.141 | 0.121 | 0.157 |  | 0.245 | 0.223 | 0.266 |  | 0.295 | 0.273 | 0.313 |

#### Supplementary file S.1.2. Pseudo R-square comparison across countries (interval regression)

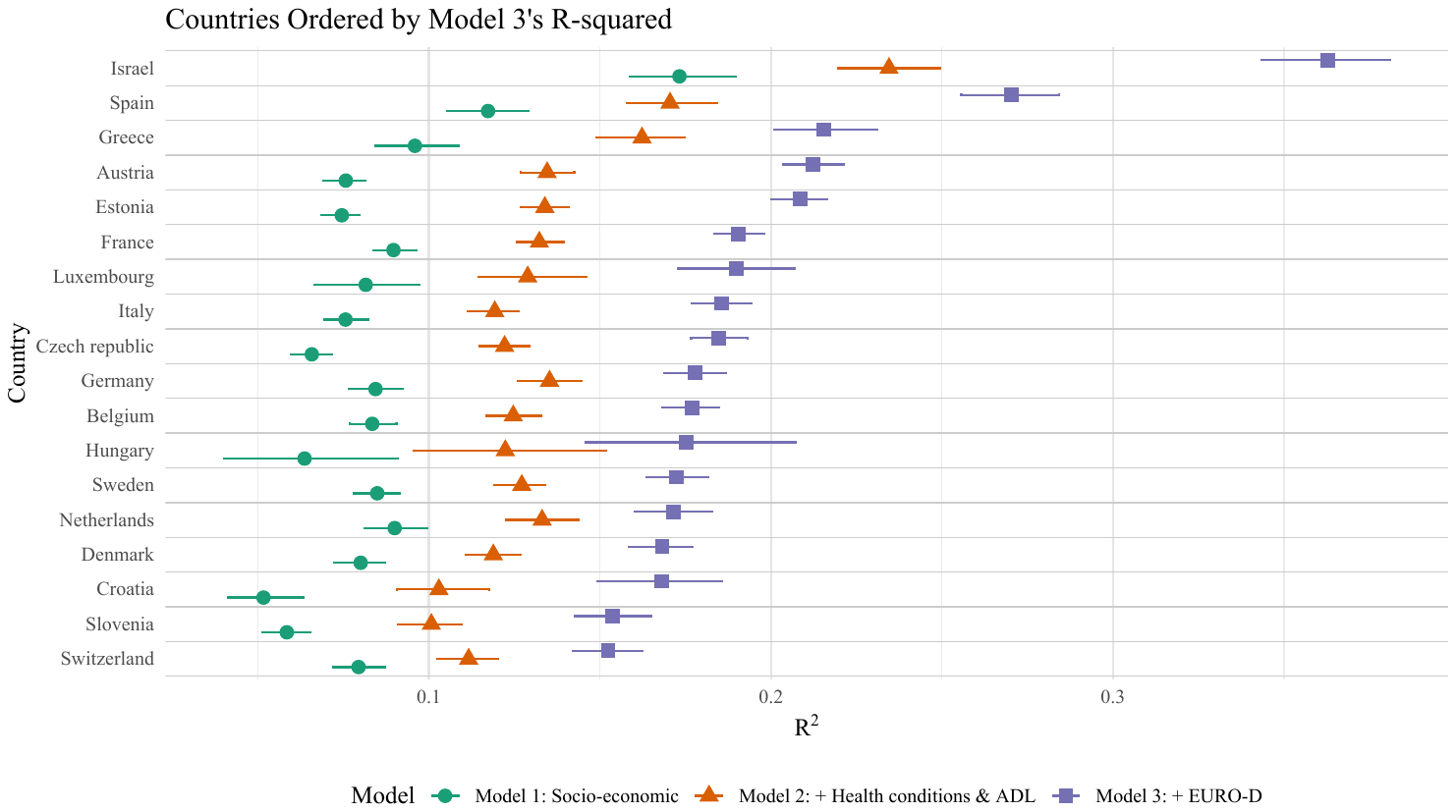

#### Supplementary file S.1.3. Pseudo R-square comparison across countries (linear regression), stratification by gender

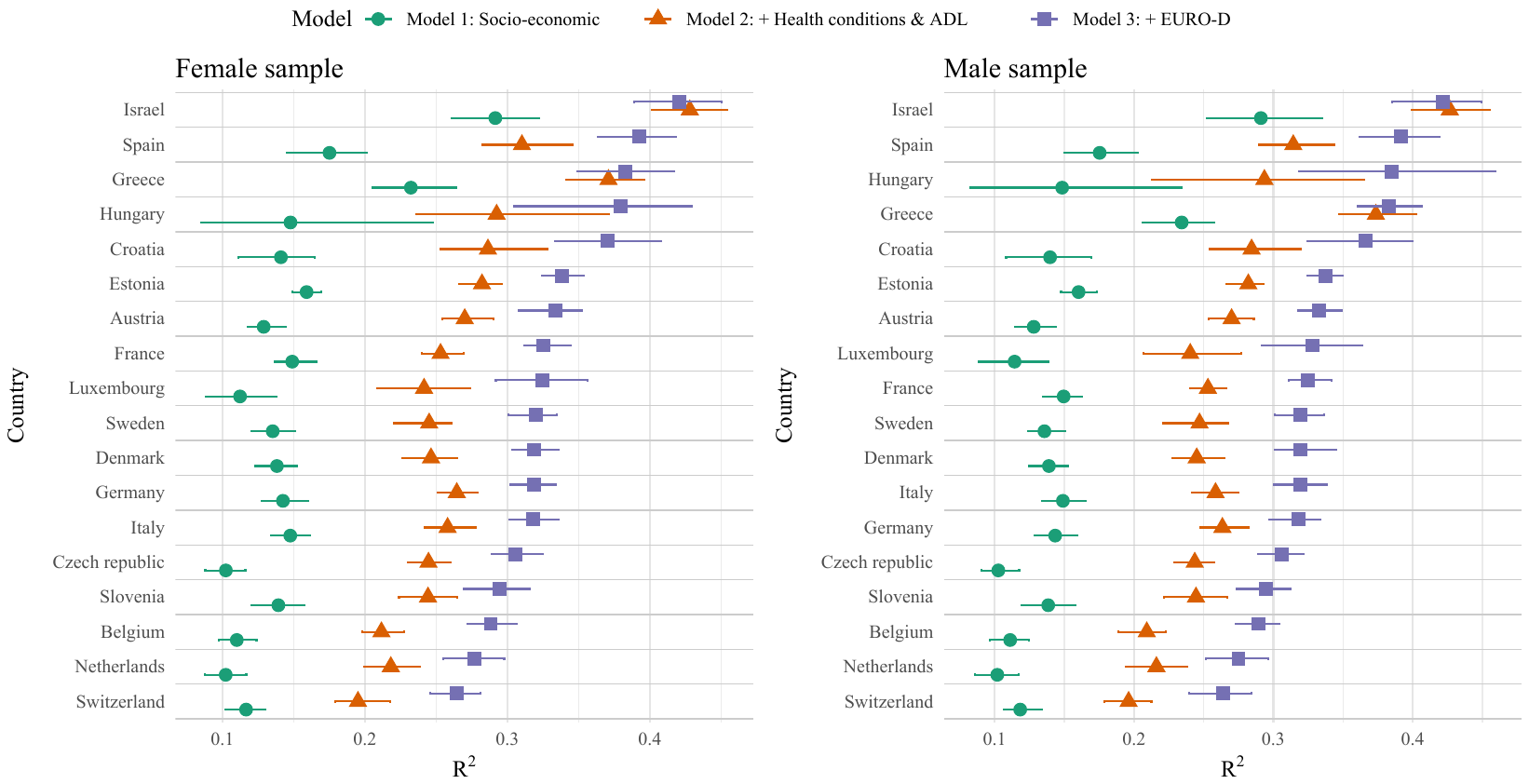

### Supplementary file S.2. Partial correlations

#### Supplementary file S.2.1. Partial correlations for the full population, including 95%CI

| Country | Variable1 | Variable2 | Partial Correlation | CI lower | CI upper |
| --- | --- | --- | --- | --- | --- |
| Austria | SRH | EUROD | 0.262 | 0.244 | 0.279 |
| Austria | SRH | COND | 0.292 | 0.274 | 0.309 |
| Austria | SRH | ADL | 0.515 | 0.501 | 0.529 |
| Austria | EUROD | COND | 0.000 | -0.019 | 0.019 |
| Austria | EUROD | ADL | 0.179 | 0.161 | 0.198 |
| Austria | COND | ADL | 0.055 | 0.036 | 0.074 |
| Belgium | SRH | EUROD | 0.306 | 0.290 | 0.321 |
| Belgium | SRH | COND | 0.228 | 0.212 | 0.244 |
| Belgium | SRH | ADL | 0.381 | 0.366 | 0.395 |
| Belgium | EUROD | COND | 0.000 | -0.017 | 0.017 |
| Belgium | EUROD | ADL | 0.174 | 0.158 | 0.190 |
| Belgium | COND | ADL | 0.133 | 0.116 | 0.150 |
| Croatia | SRH | EUROD | 0.296 | 0.256 | 0.335 |
| Croatia | SRH | COND | 0.312 | 0.273 | 0.350 |
| Croatia | SRH | ADL | 0.449 | 0.414 | 0.483 |
| Croatia | EUROD | COND | 0.076 | 0.033 | 0.119 |
| Croatia | EUROD | ADL | 0.203 | 0.162 | 0.244 |
| Croatia | COND | ADL | 0.052 | 0.009 | 0.094 |
| Czech | SRH | EUROD | 0.253 | 0.237 | 0.268 |
| Czech | SRH | COND | 0.308 | 0.293 | 0.323 |
| Czech | SRH | ADL | 0.479 | 0.466 | 0.492 |
| Czech | EUROD | COND | 0.020 | 0.003 | 0.036 |
| Czech | EUROD | ADL | 0.188 | 0.172 | 0.204 |
| Czech | COND | ADL | 0.067 | 0.051 | 0.084 |
| Denmark | SRH | EUROD | 0.289 | 0.269 | 0.309 |
| Denmark | SRH | COND | 0.264 | 0.244 | 0.284 |
| Denmark | SRH | ADL | 0.488 | 0.471 | 0.504 |
| Denmark | EUROD | COND | -0.021 | -0.043 | 0.001 |
| Denmark | EUROD | ADL | 0.111 | 0.089 | 0.133 |
| Denmark | COND | ADL | 0.095 | 0.074 | 0.117 |
| Estonia | SRH | EUROD | 0.266 | 0.251 | 0.281 |
| Estonia | SRH | COND | 0.322 | 0.308 | 0.336 |
| Estonia | SRH | ADL | 0.459 | 0.447 | 0.472 |
| Estonia | EUROD | COND | 0.045 | 0.029 | 0.061 |
| Estonia | EUROD | ADL | 0.169 | 0.154 | 0.185 |
| Estonia | COND | ADL | 0.094 | 0.079 | 0.110 |
| France | SRH | EUROD | 0.284 | 0.267 | 0.301 |
| France | SRH | COND | 0.291 | 0.275 | 0.308 |
| France | SRH | ADL | 0.440 | 0.425 | 0.454 |
| France | EUROD | COND | -0.015 | -0.033 | 0.003 |
| France | EUROD | ADL | 0.150 | 0.132 | 0.167 |
| France | COND | ADL | 0.082 | 0.064 | 0.100 |
| Germany | SRH | EUROD | 0.260 | 0.242 | 0.279 |
| Germany | SRH | COND | 0.298 | 0.279 | 0.316 |
| Germany | SRH | ADL | 0.470 | 0.454 | 0.485 |
| Germany | EUROD | COND | -0.019 | -0.039 | 0.001 |
| Germany | EUROD | ADL | 0.141 | 0.121 | 0.160 |
| Germany | COND | ADL | 0.096 | 0.076 | 0.116 |
| Greece | SRH | EUROD | 0.165 | 0.135 | 0.195 |
| Greece | SRH | COND | 0.481 | 0.457 | 0.504 |
| Greece | SRH | ADL | 0.454 | 0.429 | 0.478 |
| Greece | EUROD | COND | 0.011 | -0.020 | 0.041 |
| Greece | EUROD | ADL | 0.240 | 0.211 | 0.269 |
| Greece | COND | ADL | 0.000 | -0.031 | 0.031 |
| Hungary | SRH | EUROD | 0.345 | 0.311 | 0.379 |
| Hungary | SRH | COND | 0.374 | 0.340 | 0.407 |
| Hungary | SRH | ADL | 0.265 | 0.229 | 0.301 |
| Hungary | EUROD | COND | 0.038 | -0.001 | 0.077 |
| Hungary | EUROD | ADL | 0.185 | 0.148 | 0.223 |
| Hungary | COND | ADL | 0.192 | 0.154 | 0.229 |
| Israel | SRH | EUROD | 0.153 | 0.121 | 0.184 |
| Israel | SRH | COND | 0.388 | 0.360 | 0.415 |
| Israel | SRH | ADL | 0.573 | 0.551 | 0.594 |
| Israel | EUROD | COND | 0.087 | 0.054 | 0.119 |
| Israel | EUROD | ADL | 0.282 | 0.252 | 0.311 |
| Israel | COND | ADL | 0.000 | -0.032 | 0.032 |
| Italy | SRH | EUROD | 0.293 | 0.276 | 0.310 |
| Italy | SRH | COND | 0.328 | 0.312 | 0.345 |
| Italy | SRH | ADL | 0.340 | 0.324 | 0.356 |
| Italy | EUROD | COND | 0.028 | 0.009 | 0.046 |
| Italy | EUROD | ADL | 0.269 | 0.252 | 0.286 |
| Italy | COND | ADL | 0.091 | 0.073 | 0.110 |
| Luxembourg | SRH | EUROD | 0.270 | 0.234 | 0.305 |
| Luxembourg | SRH | COND | 0.280 | 0.244 | 0.315 |
| Luxembourg | SRH | ADL | 0.476 | 0.446 | 0.505 |
| Luxembourg | EUROD | COND | 0.021 | -0.018 | 0.059 |
| Luxembourg | EUROD | ADL | 0.204 | 0.167 | 0.240 |
| Luxembourg | COND | ADL | 0.000 | -0.038 | 0.038 |
| Netherlands | SRH | EUROD | 0.260 | 0.236 | 0.284 |
| Netherlands | SRH | COND | 0.342 | 0.320 | 0.365 |
| Netherlands | SRH | ADL | 0.479 | 0.460 | 0.499 |
| Netherlands | EUROD | COND | 0.000 | -0.026 | 0.026 |
| Netherlands | EUROD | ADL | 0.117 | 0.091 | 0.142 |
| Netherlands | COND | ADL | 0.000 | -0.026 | 0.026 |
| Slovenia | SRH | EUROD | 0.242 | 0.222 | 0.262 |
| Slovenia | SRH | COND | 0.272 | 0.253 | 0.292 |
| Slovenia | SRH | ADL | 0.419 | 0.401 | 0.437 |
| Slovenia | EUROD | COND | 0.008 | -0.014 | 0.029 |
| Slovenia | EUROD | ADL | 0.209 | 0.188 | 0.229 |
| Slovenia | COND | ADL | 0.125 | 0.104 | 0.146 |
| Spain | SRH | EUROD | 0.329 | 0.313 | 0.344 |
| Spain | SRH | COND | 0.282 | 0.267 | 0.298 |
| Spain | SRH | ADL | 0.426 | 0.412 | 0.440 |
| Spain | EUROD | COND | 0.025 | 0.008 | 0.042 |
| Spain | EUROD | ADL | 0.224 | 0.208 | 0.240 |
| Spain | COND | ADL | 0.077 | 0.060 | 0.094 |
| Sweden | SRH | EUROD | 0.274 | 0.256 | 0.292 |
| Sweden | SRH | COND | 0.336 | 0.318 | 0.353 |
| Sweden | SRH | ADL | 0.424 | 0.408 | 0.441 |
| Sweden | EUROD | COND | 0.000 | -0.020 | 0.020 |
| Sweden | EUROD | ADL | 0.134 | 0.114 | 0.153 |
| Sweden | COND | ADL | 0.035 | 0.015 | 0.055 |
| Switzerland | SRH | EUROD | 0.258 | 0.237 | 0.278 |
| Switzerland | SRH | COND | 0.251 | 0.231 | 0.271 |
| Switzerland | SRH | ADL | 0.389 | 0.370 | 0.407 |
| Switzerland | EUROD | COND | 0.000 | -0.022 | 0.022 |
| Switzerland | EUROD | ADL | 0.163 | 0.142 | 0.183 |
| Switzerland | COND | ADL | 0.137 | 0.116 | 0.158 |

Note: Partial correlations between variables estimated using EBICglasso. Values represent regularized partial correlation coefficients. 95% confidence intervals (CI) were approximated using Fisher’s z-transformation.

#### Supplementary file S.2.2. Partial correlations by gender, including 95%CI

|  |  |  | Male | | |  | Female | | |  | Difference |
| --- | --- | --- | --- | --- | --- | --- | --- | --- | --- | --- | --- |
| Country | Variable1 | Variable2 | Partial correlation | CI - | CI + |  | Partial correlation | CI - | CI + |  |  |
| Austria | SRH | EUROD | 0.353 | 0.327 | 0.379 |  | 0.347 | 0.325 | 0.369 |  | 0.006 |
| Austria | SRH | COND | 0.289 | 0.261 | 0.316 |  | 0.256 | 0.233 | 0.279 |  | 0.033 |
| Austria | SRH | ADL | 0.245 | 0.217 | 0.273 |  | 0.240 | 0.216 | 0.264 |  | 0.005 |
| Austria | EUROD | COND | -0.007 | -0.037 | 0.023 |  | 0.031 | 0.006 | 0.056 |  | -0.038 |
| Austria | EUROD | ADL | 0.215 | 0.187 | 0.243 |  | 0.181 | 0.157 | 0.205 |  | 0.034 |
| Austria | COND | ADL | -0.007 | -0.036 | 0.023 |  | 0.029 | 0.004 | 0.054 |  | -0.036 |
| Belgium | SRH | EUROD | 0.345 | 0.323 | 0.367 |  | 0.343 | 0.322 | 0.363 |  | 0.002 |
| Belgium | SRH | COND | 0.214 | 0.19 | 0.238 |  | 0.196 | 0.174 | 0.218 |  | 0.018 |
| Belgium | SRH | ADL | 0.195 | 0.171 | 0.219 |  | 0.262 | 0.240 | 0.283 |  | -0.067 |
| Belgium | EUROD | COND | 0.01 | -0.015 | 0.035 |  | 0.030 | 0.007 | 0.053 |  | -0.020 |
| Belgium | EUROD | ADL | 0.183 | 0.159 | 0.207 |  | 0.137 | 0.114 | 0.160 |  | 0.046 |
| Belgium | COND | ADL | 0.046 | 0.021 | 0.071 |  | 0.063 | 0.040 | 0.086 |  | -0.017 |
| Croatia | SRH | EUROD | 0.389 | 0.334 | 0.442 |  | 0.381 | 0.330 | 0.430 |  | 0.008 |
| Croatia | SRH | COND | 0.302 | 0.243 | 0.359 |  | 0.262 | 0.207 | 0.316 |  | 0.040 |
| Croatia | SRH | ADL | 0.191 | 0.129 | 0.252 |  | 0.189 | 0.132 | 0.245 |  | 0.002 |
| Croatia | EUROD | COND | 0.042 | -0.022 | 0.105 |  | 0.070 | 0.011 | 0.128 |  | -0.028 |
| Croatia | EUROD | ADL | 0.191 | 0.129 | 0.251 |  | 0.214 | 0.158 | 0.269 |  | -0.023 |
| Croatia | COND | ADL | -0.001 | -0.065 | 0.062 |  | 0.016 | -0.043 | 0.074 |  | -0.017 |
| Czech | SRH | EUROD | 0.328 | 0.305 | 0.351 |  | 0.335 | 0.316 | 0.354 |  | -0.007 |
| Czech | SRH | COND | 0.269 | 0.245 | 0.293 |  | 0.273 | 0.253 | 0.293 |  | -0.004 |
| Czech | SRH | ADL | 0.22 | 0.196 | 0.245 |  | 0.250 | 0.230 | 0.271 |  | -0.030 |
| Czech | EUROD | COND | 0.01 | -0.016 | 0.035 |  | 0.033 | 0.011 | 0.055 |  | -0.023 |
| Czech | EUROD | ADL | 0.219 | 0.194 | 0.244 |  | 0.188 | 0.167 | 0.209 |  | 0.031 |
| Czech | COND | ADL | 0.013 | -0.013 | 0.039 |  | 0.010 | -0.011 | 0.032 |  | 0.003 |
| Denmark | SRH | EUROD | 0.36 | 0.332 | 0.388 |  | 0.361 | 0.335 | 0.387 |  | -0.001 |
| Denmark | SRH | COND | 0.27 | 0.24 | 0.299 |  | 0.245 | 0.217 | 0.273 |  | 0.025 |
| Denmark | SRH | ADL | 0.244 | 0.214 | 0.274 |  | 0.245 | 0.217 | 0.273 |  | -0.001 |
| Denmark | EUROD | COND | -0.025 | -0.057 | 0.006 |  | 0.002 | -0.028 | 0.032 |  | -0.027 |
| Denmark | EUROD | ADL | 0.128 | 0.097 | 0.159 |  | 0.118 | 0.088 | 0.147 |  | 0.010 |
| Denmark | COND | ADL | 0.037 | 0.006 | 0.069 |  | 0.019 | -0.011 | 0.049 |  | 0.018 |
| Estonia | SRH | EUROD | 0.303 | 0.28 | 0.326 |  | 0.322 | 0.304 | 0.341 |  | -0.019 |
| Estonia | SRH | COND | 0.292 | 0.268 | 0.315 |  | 0.279 | 0.260 | 0.298 |  | 0.013 |
| Estonia | SRH | ADL | 0.232 | 0.208 | 0.257 |  | 0.239 | 0.220 | 0.258 |  | -0.007 |
| Estonia | EUROD | COND | 0.037 | 0.011 | 0.063 |  | 0.050 | 0.030 | 0.071 |  | -0.013 |
| Estonia | EUROD | ADL | 0.216 | 0.192 | 0.241 |  | 0.172 | 0.153 | 0.192 |  | 0.044 |
| Estonia | COND | ADL | 0.043 | 0.017 | 0.068 |  | 0.030 | 0.010 | 0.051 |  | 0.013 |
| France | SRH | EUROD | 0.32 | 0.295 | 0.345 |  | 0.343 | 0.322 | 0.364 |  | -0.023 |
| France | SRH | COND | 0.279 | 0.254 | 0.304 |  | 0.243 | 0.220 | 0.266 |  | 0.036 |
| France | SRH | ADL | 0.225 | 0.198 | 0.25 |  | 0.252 | 0.229 | 0.274 |  | -0.027 |
| France | EUROD | COND | 0.02 | -0.008 | 0.047 |  | -0.004 | -0.028 | 0.020 |  | 0.024 |
| France | EUROD | ADL | 0.18 | 0.153 | 0.207 |  | 0.115 | 0.092 | 0.139 |  | 0.065 |
| France | COND | ADL | 0.014 | -0.014 | 0.041 |  | 0.049 | 0.025 | 0.073 |  | -0.035 |
| Germany | SRH | EUROD | 0.298 | 0.272 | 0.324 |  | 0.321 | 0.296 | 0.346 |  | -0.023 |
| Germany | SRH | COND | 0.278 | 0.251 | 0.304 |  | 0.268 | 0.241 | 0.293 |  | 0.010 |
| Germany | SRH | ADL | 0.245 | 0.218 | 0.272 |  | 0.243 | 0.217 | 0.269 |  | 0.002 |
| Germany | EUROD | COND | -0.002 | -0.03 | 0.027 |  | 0.006 | -0.022 | 0.034 |  | -0.008 |
| Germany | EUROD | ADL | 0.162 | 0.134 | 0.189 |  | 0.136 | 0.109 | 0.163 |  | 0.026 |
| Germany | COND | ADL | 0.022 | -0.006 | 0.051 |  | 0.038 | 0.011 | 0.066 |  | -0.016 |
| Greece | SRH | EUROD | 0.216 | 0.172 | 0.26 |  | 0.323 | 0.286 | 0.360 |  | -0.107 |
| Greece | SRH | COND | 0.435 | 0.397 | 0.471 |  | 0.370 | 0.333 | 0.405 |  | 0.065 |
| Greece | SRH | ADL | 0.21 | 0.166 | 0.254 |  | 0.212 | 0.172 | 0.252 |  | -0.002 |
| Greece | EUROD | COND | 0.045 | -0.001 | 0.09 |  | 0.017 | -0.025 | 0.058 |  | 0.028 |
| Greece | EUROD | ADL | 0.176 | 0.132 | 0.22 |  | 0.174 | 0.134 | 0.214 |  | 0.002 |
| Greece | COND | ADL | -0.04 | -0.086 | 0.006 |  | -0.011 | -0.052 | 0.031 |  | -0.029 |
| Hungary | SRH | EUROD | 0.398 | 0.347 | 0.446 |  | 0.361 | 0.314 | 0.405 |  | 0.037 |
| Hungary | SRH | COND | 0.336 | 0.283 | 0.387 |  | 0.318 | 0.271 | 0.365 |  | 0.018 |
| Hungary | SRH | ADL | 0.153 | 0.095 | 0.209 |  | 0.120 | 0.069 | 0.171 |  | 0.033 |
| Hungary | EUROD | COND | 0.006 | -0.052 | 0.065 |  | 0.058 | 0.006 | 0.110 |  | -0.052 |
| Hungary | EUROD | ADL | 0.188 | 0.131 | 0.244 |  | 0.211 | 0.161 | 0.261 |  | -0.023 |
| Hungary | COND | ADL | 0.057 | -0.001 | 0.115 |  | 0.045 | -0.008 | 0.097 |  | 0.012 |
| Israel | SRH | EUROD | 0.303 | 0.257 | 0.346 |  | 0.355 | 0.317 | 0.392 |  | -0.052 |
| Israel | SRH | COND | 0.386 | 0.344 | 0.427 |  | 0.328 | 0.290 | 0.366 |  | 0.058 |
| Israel | SRH | ADL | 0.272 | 0.226 | 0.317 |  | 0.255 | 0.214 | 0.295 |  | 0.017 |
| Israel | EUROD | COND | 0.032 | -0.017 | 0.081 |  | 0.080 | 0.037 | 0.123 |  | -0.048 |
| Israel | EUROD | ADL | 0.328 | 0.284 | 0.371 |  | 0.251 | 0.210 | 0.291 |  | 0.077 |
| Israel | COND | ADL | -0.064 | -0.113 | -0.015 |  | -0.007 | -0.050 | 0.036 |  | -0.057 |
| Italy | SRH | EUROD | 0.319 | 0.295 | 0.343 |  | 0.373 | 0.351 | 0.394 |  | -0.054 |
| Italy | SRH | COND | 0.312 | 0.288 | 0.336 |  | 0.253 | 0.230 | 0.277 |  | 0.059 |
| Italy | SRH | ADL | 0.107 | 0.081 | 0.134 |  | 0.195 | 0.171 | 0.220 |  | -0.088 |
| Italy | EUROD | COND | 0.038 | 0.011 | 0.065 |  | 0.075 | 0.049 | 0.100 |  | -0.037 |
| Italy | EUROD | ADL | 0.301 | 0.276 | 0.325 |  | 0.193 | 0.168 | 0.217 |  | 0.108 |
| Italy | COND | ADL | 0.024 | -0.003 | 0.051 |  | 0.028 | 0.003 | 0.054 |  | -0.004 |
| Luxembourg | SRH | EUROD | 0.333 | 0.283 | 0.381 |  | 0.371 | 0.324 | 0.416 |  | -0.038 |
| Luxembourg | SRH | COND | 0.222 | 0.169 | 0.274 |  | 0.238 | 0.187 | 0.287 |  | -0.016 |
| Luxembourg | SRH | ADL | 0.234 | 0.18 | 0.285 |  | 0.225 | 0.174 | 0.275 |  | 0.009 |
| Luxembourg | EUROD | COND | 0.051 | -0.005 | 0.106 |  | 0.030 | -0.023 | 0.083 |  | 0.021 |
| Luxembourg | EUROD | ADL | 0.219 | 0.165 | 0.271 |  | 0.171 | 0.119 | 0.222 |  | 0.048 |
| Luxembourg | COND | ADL | 0 | -0.055 | 0.056 |  | -0.015 | -0.068 | 0.038 |  | 0.015 |
| Netherlands | SRH | EUROD | 0.306 | 0.272 | 0.34 |  | 0.313 | 0.282 | 0.344 |  | -0.007 |
| Netherlands | SRH | COND | 0.319 | 0.285 | 0.353 |  | 0.273 | 0.241 | 0.305 |  | 0.046 |
| Netherlands | SRH | ADL | 0.152 | 0.115 | 0.188 |  | 0.251 | 0.218 | 0.283 |  | -0.099 |
| Netherlands | EUROD | COND | 0 | -0.038 | 0.037 |  | 0.013 | -0.022 | 0.048 |  | -0.013 |
| Netherlands | EUROD | ADL | 0.156 | 0.119 | 0.192 |  | 0.111 | 0.076 | 0.145 |  | 0.045 |
| Netherlands | COND | ADL | 0.009 | -0.029 | 0.047 |  | 0.004 | -0.030 | 0.039 |  | 0.005 |
| Slovenia | SRH | EUROD | 0.296 | 0.266 | 0.325 |  | 0.321 | 0.295 | 0.346 |  | -0.025 |
| Slovenia | SRH | COND | 0.251 | 0.221 | 0.281 |  | 0.252 | 0.226 | 0.279 |  | -0.001 |
| Slovenia | SRH | ADL | 0.204 | 0.173 | 0.235 |  | 0.215 | 0.188 | 0.242 |  | -0.011 |
| Slovenia | EUROD | COND | -0.005 | -0.037 | 0.028 |  | 0.048 | 0.019 | 0.076 |  | -0.053 |
| Slovenia | EUROD | ADL | 0.205 | 0.174 | 0.236 |  | 0.194 | 0.166 | 0.221 |  | 0.011 |
| Slovenia | COND | ADL | 0.045 | 0.012 | 0.077 |  | 0.027 | -0.001 | 0.056 |  | 0.018 |
| Spain | SRH | EUROD | 0.378 | 0.357 | 0.399 |  | 0.426 | 0.406 | 0.444 |  | -0.048 |
| Spain | SRH | COND | 0.257 | 0.234 | 0.28 |  | 0.246 | 0.224 | 0.268 |  | 0.011 |
| Spain | SRH | ADL | 0.175 | 0.15 | 0.199 |  | 0.199 | 0.176 | 0.221 |  | -0.024 |
| Spain | EUROD | COND | 0.025 | 0 | 0.05 |  | 0.056 | 0.033 | 0.079 |  | -0.031 |
| Spain | EUROD | ADL | 0.234 | 0.211 | 0.258 |  | 0.216 | 0.194 | 0.238 |  | 0.018 |
| Spain | COND | ADL | 0.022 | -0.003 | 0.047 |  | 0.015 | -0.008 | 0.038 |  | 0.007 |
| Sweden | SRH | EUROD | 0.303 | 0.276 | 0.329 |  | 0.345 | 0.321 | 0.369 |  | -0.042 |
| Sweden | SRH | COND | 0.294 | 0.267 | 0.321 |  | 0.292 | 0.267 | 0.317 |  | 0.002 |
| Sweden | SRH | ADL | 0.201 | 0.173 | 0.229 |  | 0.224 | 0.198 | 0.250 |  | -0.023 |
| Sweden | EUROD | COND | 0.014 | -0.015 | 0.043 |  | 0.001 | -0.026 | 0.028 |  | 0.013 |
| Sweden | EUROD | ADL | 0.144 | 0.116 | 0.173 |  | 0.121 | 0.094 | 0.148 |  | 0.023 |
| Sweden | COND | ADL | 0.013 | -0.016 | 0.042 |  | 0.009 | -0.019 | 0.036 |  | 0.004 |
| Switzerland | SRH | EUROD | 0.313 | 0.284 | 0.341 |  | 0.313 | 0.286 | 0.339 |  | 0.000 |
| Switzerland | SRH | COND | 0.246 | 0.216 | 0.276 |  | 0.231 | 0.203 | 0.259 |  | 0.015 |
| Switzerland | SRH | ADL | 0.158 | 0.127 | 0.188 |  | 0.201 | 0.173 | 0.230 |  | -0.043 |
| Switzerland | EUROD | COND | 0.027 | -0.004 | 0.059 |  | 0.053 | 0.023 | 0.082 |  | -0.026 |
| Switzerland | EUROD | ADL | 0.164 | 0.133 | 0.195 |  | 0.110 | 0.081 | 0.139 |  | 0.054 |
| Switzerland | COND | ADL | 0.052 | 0.02 | 0.083 |  | 0.037 | 0.008 | 0.067 |  | 0.015 |

#### Supplementary file S.2.3. Partial correlations plot (male only)

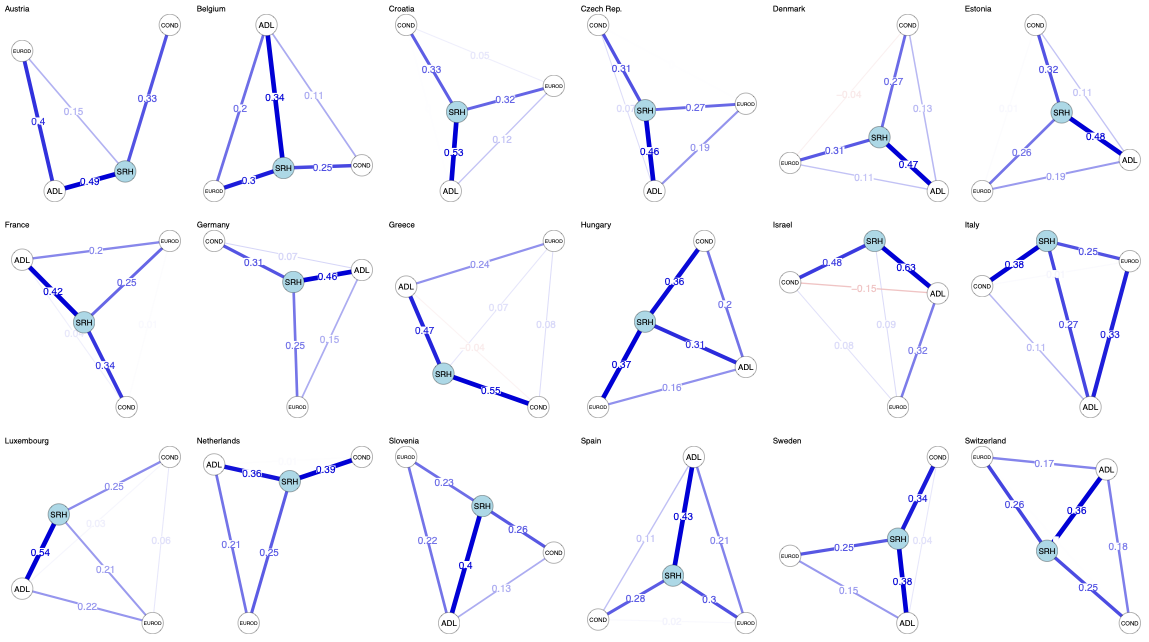

#### Supplementary file S.2.4. Partial correlations plot (female only)

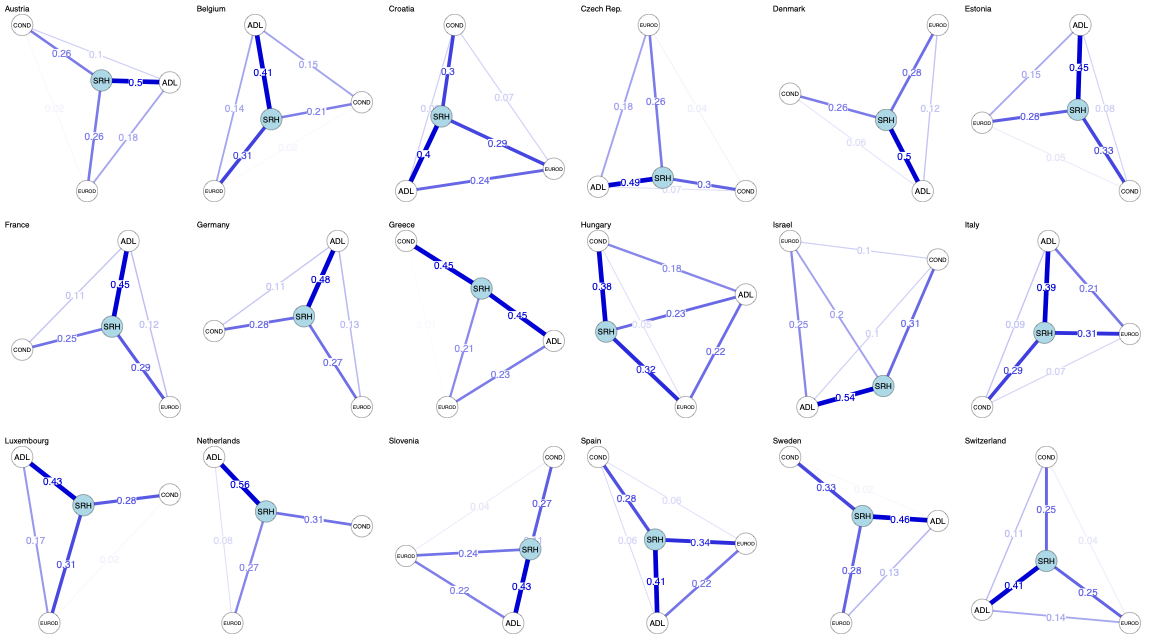

### Supplementary file S.3. Blinder Oaxaca decomposition

#### Supplementary file S.3.1. Blinder Oaxaca Linear results

| Austria | | |  |  |  |  |
| --- | --- | --- | --- | --- | --- | --- |
| Adjustment | EUROD-D | | COND, ADL | | SES | |
| group_1 | 2.987 | *** | 2.987 | *** | 2.987 | *** |
|  | [2.964 3.009] |  | [2.964 3.009] |  | [2.964 3.009] |  |
| group_2 | 3.243 | *** | 3.243 | *** | 3.243 | *** |
|  | [3.230 3.256] |  | [3.230 3.256] |  | [3.230 3.256] |  |
| difference | -0.256 | *** | -0.256 | *** | -0.256 | *** |
|  | [-0.282 -0.230] |  | [-0.282 -0.230] |  | [-0.282 -0.230] |  |
| explained | -0.112 | *** | -0.113 | *** | -0.126 | *** |
|  | [-0.124 -0.101] |  | [-0.128 -0.099] |  | [-0.143 -0.110] |  |
| unexplained | -0.144 | *** | -0.143 | *** | -0.130 | *** |
|  | [-0.168 -0.121] |  | [-0.165 -0.121] |  | [-0.151 -0.109] |  |
| EUROD | -0.114 | *** | -0.088 | *** | -0.082 | *** |
|  | [-0.125 -0.103] |  | [-0.097 -0.080] |  | [-0.091 -0.074] |  |
| wave | 0.002 | * |  |  |  |  |
|  | [0.000 0.004] |  |  |  |  |  |
| wave== 4.0000 | |  | 0.000 |  | 0.000 |  |
|  |  |  | [-0.001 0.002] |  | [-0.000 0.001] |  |
| wave== 6.0000 | |  | 0.001 |  | 0.001 | * |
|  |  |  | [-0.000 0.001] |  | [0.000 0.002] |  |
| ADL |  |  | -0.008 | *** | -0.007 | *** |
|  |  |  | [-0.012 -0.003] |  | [-0.010 -0.003] |  |
| COND |  |  | -0.018 | *** | -0.016 | *** |
|  |  |  | [-0.025 -0.011] |  | [-0.022 -0.010] |  |
| agegroup1 | |  |  |  | 0.000 |  |
|  |  |  |  |  | [-0.001 0.001] |  |
| agegroup3 | |  |  |  | -0.001 | ** |
|  |  |  |  |  | [-0.002 -0.001] |  |
| female |  |  |  |  | -0.001 | ** |
|  |  |  |  |  | [-0.002 -0.000] |  |
| partnerinhh== 3.0000 | | |  |  | 0.001 |  |
|  |  |  |  |  | [-0.000 0.001] |  |
| bieduc |  |  |  |  | -0.019 | *** |
|  |  |  |  |  | [-0.022 -0.016] |  |
| empl==employed or self-employed | | | |  | 0.010 | *** |
|  |  |  |  |  | [0.007 0.013] |  |
| empl==non employed | |  |  |  | -0.007 | *** |
|  |  |  |  |  | [-0.010 -0.005] |  |
| empl==other | |  |  |  | 0.001 | ** |
|  |  |  |  |  | [0.000 0.003] |  |
| RECODE of hin_adjlog (hin_adjlog) | | | |  | -0.006 | * |
|  |  |  |  |  | [-0.012 -0.000] |  |
| housing==NA | |  |  |  | 0.000 |  |
|  |  |  |  |  | [-0.000 0.000] |  |
| EUROD | 0.057 | *** | 0.036 | ** | 0.042 | *** |
|  | [0.033 0.080] |  | [0.013 0.060] |  | [0.018 0.065] |  |
| wave | -0.060 | *** |  |  |  |  |
|  | [-0.090 -0.029] |  |  |  |  |  |
| wave== 4.0000 | |  | -0.062 | *** | -0.056 | *** |
|  |  |  | [-0.079 -0.044] |  | [-0.073 -0.039] |  |
| wave== 6.0000 | |  | -0.006 |  | -0.004 |  |
|  |  |  | [-0.022 0.010] |  | [-0.020 0.011] |  |
| ADL |  |  | 0.015 | *** | 0.013 | *** |
|  |  |  | [0.008 0.022] |  | [0.006 0.021] |  |
| COND |  |  | -0.036 | * | -0.033 | * |
|  |  |  | [-0.067 -0.006] |  | [-0.062 -0.003] |  |
| agegroup1 | |  |  |  | 0.010 |  |
|  |  |  |  |  | [-0.016 0.035] |  |
| agegroup3 | |  |  |  | 0.003 |  |
|  |  |  |  |  | [-0.005 0.010] |  |
| female |  |  |  |  | -0.038 | ** |
|  |  |  |  |  | [-0.065 -0.010] |  |
| partnerinhh== 3.0000 | | |  |  | -0.006 |  |
|  |  |  |  |  | [-0.025 0.013] |  |
| bieduc |  |  |  |  | -0.005 |  |
|  |  |  |  |  | [-0.020 0.010] |  |
| empl==employed or self-employed | | | |  | 0.003 |  |
|  |  |  |  |  | [-0.009 0.015] |  |
| empl==non employed | |  |  |  | 0.005 | * |
|  |  |  |  |  | [0.001 0.010] |  |
| empl==other | |  |  |  | 0.005 |  |
|  |  |  |  |  | [-0.002 0.011] |  |
| RECODE of hin_adjlog (hin_adjlog) | | | |  | -0.742 | *** |
|  |  |  |  |  | [-1.076 -0.409] |  |
| housing==NA | |  |  |  | 0.001 |  |
|  |  |  |  |  | [-0.001 0.003] |  |
| Intercept | -0.141 | *** | -0.089 | *** | 0.673 | *** |
|  | [-0.184 -0.098] |  | [-0.138 -0.040] |  | [0.337 1.009] |  |
| N | 160805 |  | 160805 |  | 160805 |  |
| *** p<.001, ** p<.01, * p<.05 | | |  |  |  |  |

| Belgium | | |  |  |  |  |
| --- | --- | --- | --- | --- | --- | --- |
| Adjustment | EUROD-D | | COND, ADL | | SES | |
| group_1 | 3.014 | *** | 3.014 | *** | 3.014 | *** |
|  | [2.995 3.033] |  | [2.995 3.033] |  | [2.994 3.034] |  |
| group_2 | 3.243 | *** | 3.243 | *** | 3.243 | *** |
|  | [3.230 3.256] |  | [3.230 3.256] |  | [3.230 3.256] |  |
| difference | -0.229 | *** | -0.229 | *** | -0.229 | *** |
|  | [-0.252 -0.206] |  | [-0.252 -0.206] |  | [-0.253 -0.206] |  |
| explained | -0.027 | *** | 0.002 |  | -0.026 | ** |
|  | [-0.038 -0.016] |  | [-0.012 0.017] |  | [-0.044 -0.008] |  |
| unexplained | -0.202 | *** | -0.231 | *** | -0.203 | *** |
|  | [-0.222 -0.181] |  | [-0.251 -0.212] |  | [-0.223 -0.183] |  |
| EUROD | -0.022 | *** | -0.017 | *** | -0.016 | *** |
|  | [-0.033 -0.011] |  | [-0.026 -0.009] |  | [-0.024 -0.008] |  |
| wave== 4.0000 | -0.004 | *** | -0.004 | *** | -0.003 | *** |
|  | [-0.006 -0.003] |  | [-0.005 -0.002] |  | [-0.004 -0.002] |  |
| wave== 6.0000 | -0.001 | * | 0.000 |  | 0.000 |  |
|  | [-0.001 -0.000] |  | [-0.001 0.000] |  | [-0.001 0.000] |  |
| ADL |  |  | 0.025 | *** | 0.021 | *** |
|  |  |  | [0.020 0.030] |  | [0.017 0.026] |  |
| COND |  |  | -0.001 |  | -0.001 |  |
|  |  |  | [-0.008 0.006] |  | [-0.007 0.005] |  |
| agegroup1 | |  |  |  | -0.002 | *** |
|  |  |  |  |  | [-0.004 -0.001] |  |
| agegroup3 | |  |  |  | 0.001 | ** |
|  |  |  |  |  | [0.000 0.002] |  |
| female |  |  |  |  | 0.000 |  |
|  |  |  |  |  | [-0.000 0.001] |  |
| partnerinhh== 3.0000 | | |  |  | -0.001 |  |
|  |  |  |  |  | [-0.002 0.000] |  |
| bieduc |  |  |  |  | -0.021 | *** |
|  |  |  |  |  | [-0.024 -0.018] |  |
| empl==employed or self-employed | | | |  | 0.003 | * |
|  |  |  |  |  | [0.000 0.005] |  |
| empl==non employed | |  |  |  | 0.004 | *** |
|  |  |  |  |  | [0.002 0.005] |  |
| empl==other | |  |  |  | 0.000 |  |
|  |  |  |  |  | [-0.000 0.001] |  |
| RECODE of hin_adjlog (hin_adjlog) | | | |  | -0.011 |  |
|  |  |  |  |  | [-0.022 0.000] |  |
| housing==NA | |  |  |  | 0.000 |  |
|  |  |  |  |  | [-0.001 0.000] |  |
| EUROD | -0.047 | *** | -0.024 |  | -0.018 |  |
|  | [-0.072 -0.022] |  | [-0.048 0.001] |  | [-0.042 0.007] |  |
| wave== 4.0000 | -0.025 | ** | -0.025 | *** | -0.022 | ** |
|  | [-0.040 -0.009] |  | [-0.039 -0.011] |  | [-0.036 -0.009] |  |
| wave== 6.0000 | 0.003 |  | 0.000 |  | 0.004 |  |
|  | [-0.014 0.020] |  | [-0.017 0.016] |  | [-0.012 0.020] |  |
| ADL |  |  | -0.005 |  | -0.006 |  |
|  |  |  | [-0.014 0.004] |  | [-0.015 0.003] |  |
| COND |  |  | -0.156 | *** | -0.138 | *** |
|  |  |  | [-0.186 -0.127] |  | [-0.167 -0.109] |  |
| agegroup1 | |  |  |  | -0.017 |  |
|  |  |  |  |  | [-0.043 0.008] |  |
| agegroup3 | |  |  |  | -0.005 |  |
|  |  |  |  |  | [-0.013 0.003] |  |
| female |  |  |  |  | 0.011 |  |
|  |  |  |  |  | [-0.013 0.035] |  |
| partnerinhh== 3.0000 | | |  |  | -0.015 | * |
|  |  |  |  |  | [-0.030 -0.000] |  |
| bieduc |  |  |  |  | 0.013 |  |
|  |  |  |  |  | [-0.001 0.027] |  |
| empl==employed or self-employed | | | |  | 0.021 | ** |
|  |  |  |  |  | [0.008 0.035] |  |
| empl==non employed | |  |  |  | 0.015 | *** |
|  |  |  |  |  | [0.007 0.022] |  |
| empl==other | |  |  |  | 0.009 | * |
|  |  |  |  |  | [0.002 0.016] |  |
| RECODE of hin_adjlog (hin_adjlog) | | | |  | -0.193 |  |
|  |  |  |  |  | [-0.433 0.047] |  |
| housing==NA | |  |  |  | 0.001 |  |
|  |  |  |  |  | [-0.001 0.004] |  |
| Intercept | -0.134 | *** | -0.021 |  | 0.137 |  |
|  | [-0.173 -0.094] |  | [-0.068 0.027] |  | [-0.101 0.374] |  |
| N | 163540 |  | 163540 |  | 163540 |  |
| *** p<.001, ** p<.01, * p<.05 | | |  |  |  |  |

| Croatia | | |  |  |  |  |
| --- | --- | --- | --- | --- | --- | --- |
| Adjustment | EUROD-D | | COND, ADL | | SES | |
| group_1 | 3.114 | *** | 3.114 | *** | 3.114 | *** |
|  | [3.071 3.157] |  | [3.071 3.157] |  | [3.071 3.157] |  |
| group_2 | 3.243 | *** | 3.243 | *** | 3.243 | *** |
|  | [3.230 3.256] |  | [3.230 3.256] |  | [3.230 3.256] |  |
| difference | -0.129 | *** | -0.129 | *** | -0.129 | *** |
|  | [-0.174 -0.085] |  | [-0.174 -0.085] |  | [-0.174 -0.084] |  |
| explained | -0.045 | *** | -0.015 |  | -0.026 |  |
|  | [-0.067 -0.023] |  | [-0.041 0.011] |  | [-0.059 0.007] |  |
| unexplained | -0.084 | *** | -0.115 | *** | -0.104 | *** |
|  | [-0.124 -0.044] |  | [-0.152 -0.077] |  | [-0.144 -0.063] |  |
| EUROD | -0.004 |  | -0.003 |  | -0.003 |  |
|  | [-0.023 0.016] |  | [-0.018 0.012] |  | [-0.017 0.012] |  |
| wave== 4.0000 | -0.038 | *** | -0.033 | *** | -0.027 | *** |
|  | [-0.048 -0.028] |  | [-0.042 -0.024] |  | [-0.036 -0.018] |  |
| wave== 6.0000 | -0.004 | *** | -0.001 |  | -0.002 | * |
|  | [-0.006 -0.002] |  | [-0.003 0.000] |  | [-0.004 -0.000] |  |
| ADL |  |  | 0.003 |  | 0.003 |  |
|  |  |  | [-0.005 0.012] |  | [-0.004 0.010] |  |
| COND |  |  | 0.019 | *** | 0.017 | *** |
|  |  |  | [0.008 0.031] |  | [0.007 0.027] |  |
| agegroup1 | |  |  |  | -0.005 | *** |
|  |  |  |  |  | [-0.008 -0.003] |  |
| agegroup3 | |  |  |  | 0.000 |  |
|  |  |  |  |  | [-0.002 0.001] |  |
| female |  |  |  |  | 0.000 |  |
|  |  |  |  |  | [-0.000 0.001] |  |
| partnerinhh== 3.0000 | | |  |  | 0.000 |  |
|  |  |  |  |  | [-0.000 0.001] |  |
| bieduc |  |  |  |  | -0.002 |  |
|  |  |  |  |  | [-0.005 0.002] |  |
| empl==employed or self-employed | | | |  | 0.011 | *** |
|  |  |  |  |  | [0.007 0.015] |  |
| empl==non employed | |  |  |  | 0.001 |  |
|  |  |  |  |  | [-0.002 0.003] |  |
| empl==other | |  |  |  | -0.003 | * |
|  |  |  |  |  | [-0.005 -0.001] |  |
| RECODE of hin_adjlog (hin_adjlog) | | | |  | -0.016 |  |
|  |  |  |  |  | [-0.033 0.001] |  |
| housing==NA | |  |  |  | -0.001 |  |
|  |  |  |  |  | [-0.003 0.002] |  |
| EUROD | 0.021 |  | 0.021 |  | 0.030 |  |
|  | [-0.023 0.064] |  | [-0.024 0.067] |  | [-0.017 0.077] |  |
| wave== 4.0000 | 0.000 |  | 0.000 |  | 0.000 |  |
|  | [-0.000 0.000] |  | [-0.000 0.000] |  | [-0.000 0.000] |  |
| wave== 6.0000 | 0.013 |  | 0.023 |  | 0.025 |  |
|  | [-0.021 0.047] |  | [-0.009 0.055] |  | [-0.007 0.057] |  |
| ADL |  |  | 0.010 |  | 0.014 |  |
|  |  |  | [-0.004 0.025] |  | [-0.000 0.028] |  |
| COND |  |  | -0.069 | * | -0.078 | * |
|  |  |  | [-0.129 -0.009] |  | [-0.138 -0.018] |  |
| agegroup1 | |  |  |  | -0.024 |  |
|  |  |  |  |  | [-0.076 0.027] |  |
| agegroup3 | |  |  |  | -0.020 | * |
|  |  |  |  |  | [-0.037 -0.003] |  |
| female |  |  |  |  | -0.063 | ** |
|  |  |  |  |  | [-0.109 -0.016] |  |
| partnerinhh== 3.0000 | | |  |  | 0.022 |  |
|  |  |  |  |  | [-0.012 0.056] |  |
| bieduc |  |  |  |  | -0.021 |  |
|  |  |  |  |  | [-0.043 0.000] |  |
| empl==employed or self-employed | | | |  | 0.021 | * |
|  |  |  |  |  | [0.002 0.040] |  |
| empl==non employed | |  |  |  | 0.013 | * |
|  |  |  |  |  | [0.002 0.025] |  |
| empl==other | |  |  |  | 0.019 |  |
|  |  |  |  |  | [-0.002 0.040] |  |
| RECODE of hin_adjlog (hin_adjlog) | | | |  | -0.046 |  |
|  |  |  |  |  | [-0.473 0.381] |  |
| housing==NA | |  |  |  | 0.001 |  |
|  |  |  |  |  | [-0.007 0.009] |  |
| Intercept | -0.118 | ** | -0.100 | * | 0.003 |  |
|  | [-0.191 -0.045] |  | [-0.190 -0.010] |  | [-0.445 0.451] |  |
| N | 153006 |  | 153006 |  | 153006 |  |
| *** p<.001, ** p<.01, * p<.05 | | |  |  |  |  |

| Czech Republic | | |  |  |  |  |
| --- | --- | --- | --- | --- | --- | --- |
| Adjustment | EUROD-D | | COND, ADL | | SES | |
| group_1 | 3.384 | *** | 3.384 | *** | 3.384 | *** |
|  | [3.364 3.404] |  | [3.364 3.405] |  | [3.364 3.405] |  |
| group_2 | 3.243 | *** | 3.243 | *** | 3.243 | *** |
|  | [3.230 3.256] |  | [3.230 3.256] |  | [3.230 3.256] |  |
| difference | 0.141 | *** | 0.141 | *** | 0.141 | *** |
|  | [0.117 0.165] |  | [0.117 0.165] |  | [0.117 0.165] |  |
| explained | -0.064 | *** | -0.005 |  | 0.021 | * |
|  | [-0.076 -0.052] |  | [-0.019 0.009] |  | [0.004 0.038] |  |
| unexplained | 0.205 | *** | 0.146 | *** | 0.120 | *** |
|  | [0.184 0.226] |  | [0.127 0.166] |  | [0.097 0.143] |  |
| EUROD | -0.061 | *** | -0.047 | *** | -0.044 | *** |
|  | [-0.072 -0.049] |  | [-0.056 -0.038] |  | [-0.052 -0.035] |  |
| wave== 4.0000 | -0.003 | *** | -0.003 | *** | -0.002 | *** |
|  | [-0.005 -0.002] |  | [-0.004 -0.002] |  | [-0.004 -0.001] |  |
| wave== 6.0000 | 0.000 |  | 0.000 |  | 0.000 |  |
|  | [-0.000 0.001] |  | [-0.000 0.000] |  | [-0.000 0.000] |  |
| ADL |  |  | 0.001 |  | 0.000 |  |
|  |  |  | [-0.004 0.005] |  | [-0.004 0.004] |  |
| COND |  |  | 0.044 | *** | 0.039 | *** |
|  |  |  | [0.038 0.051] |  | [0.034 0.045] |  |
| agegroup1 | |  |  |  | 0.001 | * |
|  |  |  |  |  | [0.000 0.002] |  |
| agegroup3 | |  |  |  | -0.002 | *** |
|  |  |  |  |  | [-0.003 -0.001] |  |
| female |  |  |  |  | -0.001 | ** |
|  |  |  |  |  | [-0.002 -0.000] |  |
| partnerinhh== 3.0000 | | |  |  | 0.000 |  |
|  |  |  |  |  | [-0.001 0.000] |  |
| bieduc |  |  |  |  | 0.014 | *** |
|  |  |  |  |  | [0.012 0.017] |  |
| empl==employed or self-employed | | | |  | 0.011 | *** |
|  |  |  |  |  | [0.008 0.014] |  |
| empl==non employed | |  |  |  | -0.007 | *** |
|  |  |  |  |  | [-0.009 -0.005] |  |
| empl==other | |  |  |  | 0.005 | ** |
|  |  |  |  |  | [0.001 0.009] |  |
| RECODE of hin_adjlog (hin_adjlog) | | | |  | 0.007 |  |
|  |  |  |  |  | [-0.000 0.014] |  |
| housing==NA | |  |  |  | 0.000 |  |
|  |  |  |  |  | [-0.000 0.000] |  |
| EUROD | -0.032 | ** | -0.033 | ** | -0.021 |  |
|  | [-0.053 -0.012] |  | [-0.054 -0.012] |  | [-0.043 0.002] |  |
| wave== 4.0000 | -0.016 | * | -0.013 |  | -0.005 |  |
|  | [-0.031 -0.000] |  | [-0.028 0.002] |  | [-0.019 0.010] |  |
| wave== 6.0000 | 0.023 | ** | 0.011 |  | 0.017 | * |
|  | [0.006 0.039] |  | [-0.004 0.026] |  | [0.002 0.032] |  |
| ADL |  |  | 0.007 |  | 0.009 | * |
|  |  |  | [-0.000 0.014] |  | [0.001 0.016] |  |
| COND |  |  | -0.046 | ** | -0.039 | * |
|  |  |  | [-0.080 -0.013] |  | [-0.074 -0.005] |  |
| agegroup1 | |  |  |  | -0.003 |  |
|  |  |  |  |  | [-0.027 0.021] |  |
| agegroup3 | |  |  |  | -0.001 |  |
|  |  |  |  |  | [-0.008 0.007] |  |
| female |  |  |  |  | -0.048 | *** |
|  |  |  |  |  | [-0.073 -0.024] |  |
| partnerinhh== 3.0000 | | |  |  | -0.005 |  |
|  |  |  |  |  | [-0.021 0.011] |  |
| bieduc |  |  |  |  | 0.004 |  |
|  |  |  |  |  | [-0.004 0.013] |  |
| empl==employed or self-employed | | | |  | 0.002 |  |
|  |  |  |  |  | [-0.008 0.013] |  |
| empl==non employed | |  |  |  | 0.006 | *** |
|  |  |  |  |  | [0.003 0.010] |  |
| empl==other | |  |  |  | 0.000 |  |
|  |  |  |  |  | [-0.002 0.001] |  |
| RECODE of hin_adjlog (hin_adjlog) | | | |  | -0.087 |  |
|  |  |  |  |  | [-0.321 0.147] |  |
| housing==NA | |  |  |  | 0.002 | * |
|  |  |  |  |  | [0.000 0.004] |  |
| Intercept | 0.230 | *** | 0.221 | *** | 0.288 | * |
|  | [0.190 0.271] |  | [0.171 0.270] |  | [0.041 0.535] |  |
| N | 163952 |  | 163952 |  | 163952 |  |
| *** p<.001, ** p<.01, * p<.05 | | |  |  |  |  |

| Denmark | | |  |  |  |  |
| --- | --- | --- | --- | --- | --- | --- |
| Adjustment | EUROD-D | | COND, ADL | | SES | |
| group_1 | 2.584 | *** | 2.584 | *** | 2.584 | *** |
|  | [2.558 2.611] |  | [2.558 2.611] |  | [2.558 2.611] |  |
| group_2 | 3.243 | *** | 3.243 | *** | 3.243 | *** |
|  | [3.230 3.256] |  | [3.230 3.256] |  | [3.230 3.256] |  |
| difference | -0.659 | *** | -0.659 | *** | -0.659 | *** |
|  | [-0.688 -0.629] |  | [-0.688 -0.629] |  | [-0.688 -0.629] |  |
| explained | -0.167 | *** | -0.173 | *** | -0.216 | *** |
|  | [-0.179 -0.155] |  | [-0.187 -0.158] |  | [-0.232 -0.199] |  |
| unexplained | -0.492 | *** | -0.486 | *** | -0.443 | *** |
|  | [-0.518 -0.465] |  | [-0.511 -0.461] |  | [-0.468 -0.418] |  |
| EUROD | -0.161 | *** | -0.124 | *** | -0.116 | *** |
|  | [-0.172 -0.149] |  | [-0.134 -0.115] |  | [-0.125 -0.107] |  |
| wave== 4.0000 | -0.006 | *** | -0.005 | *** | -0.004 | *** |
|  | [-0.008 -0.004] |  | [-0.007 -0.003] |  | [-0.006 -0.003] |  |
| wave== 6.0000 | 0.000 |  | 0.000 |  | 0.000 |  |
|  | [-0.000 0.000] |  | [-0.000 0.000] |  | [-0.000 0.000] |  |
| ADL |  |  | -0.015 | *** | -0.013 | *** |
|  |  |  | [-0.020 -0.010] |  | [-0.017 -0.009] |  |
| COND |  |  | -0.028 | *** | -0.024 | *** |
|  |  |  | [-0.035 -0.020] |  | [-0.031 -0.018] |  |
| agegroup1 | |  |  |  | 0.002 | ** |
|  |  |  |  |  | [0.000 0.003] |  |
| agegroup3 | |  |  |  | 0.001 | * |
|  |  |  |  |  | [0.000 0.002] |  |
| female |  |  |  |  | 0.000 |  |
|  |  |  |  |  | [-0.000 0.001] |  |
| partnerinhh== 3.0000 | | |  |  | 0.000 |  |
|  |  |  |  |  | [-0.000 0.000] |  |
| bieduc |  |  |  |  | -0.040 | *** |
|  |  |  |  |  | [-0.045 -0.035] |  |
| empl==employed or self-employed | | | |  | -0.018 | *** |
|  |  |  |  |  | [-0.021 -0.014] |  |
| empl==non employed | |  |  |  | -0.002 | * |
|  |  |  |  |  | [-0.003 -0.000] |  |
| empl==other | |  |  |  | 0.005 | ** |
|  |  |  |  |  | [0.001 0.008] |  |
| RECODE of hin_adjlog (hin_adjlog) | | | |  | -0.006 |  |
|  |  |  |  |  | [-0.013 0.000] |  |
| housing==NA | |  |  |  | 0.000 |  |
|  |  |  |  |  | [-0.001 0.000] |  |
| EUROD | 0.109 | *** | 0.097 | *** | 0.087 | *** |
|  | [0.084 0.134] |  | [0.072 0.121] |  | [0.062 0.113] |  |
| wave== 4.0000 | -0.029 | ** | -0.015 |  | -0.012 |  |
|  | [-0.048 -0.009] |  | [-0.034 0.003] |  | [-0.030 0.005] |  |
| wave== 6.0000 | 0.011 |  | 0.014 |  | 0.015 |  |
|  | [-0.009 0.030] |  | [-0.004 0.032] |  | [-0.003 0.032] |  |
| ADL |  |  | 0.029 | *** | 0.024 | *** |
|  |  |  | [0.021 0.038] |  | [0.016 0.033] |  |
| COND |  |  | -0.034 | * | -0.048 | ** |
|  |  |  | [-0.066 -0.002] |  | [-0.081 -0.016] |  |
| agegroup1 | |  |  |  | 0.015 |  |
|  |  |  |  |  | [-0.018 0.047] |  |
| agegroup3 | |  |  |  | 0.004 |  |
|  |  |  |  |  | [-0.008 0.016] |  |
| female |  |  |  |  | -0.031 | * |
|  |  |  |  |  | [-0.059 -0.004] |  |
| partnerinhh== 3.0000 | | |  |  | 0.007 |  |
|  |  |  |  |  | [-0.013 0.028] |  |
| bieduc |  |  |  |  | 0.010 |  |
|  |  |  |  |  | [-0.011 0.030] |  |
| empl==employed or self-employed | | | |  | 0.027 | * |
|  |  |  |  |  | [0.006 0.048] |  |
| empl==non employed | |  |  |  | 0.015 | *** |
|  |  |  |  |  | [0.008 0.023] |  |
| empl==other | |  |  |  | 0.000 |  |
|  |  |  |  |  | [-0.003 0.004] |  |
| RECODE of hin_adjlog (hin_adjlog) | | | |  | -1.283 | *** |
|  |  |  |  |  | [-1.815 -0.752] |  |
| housing==NA | |  |  |  | 0.001 |  |
|  |  |  |  |  | [-0.002 0.005] |  |
| Intercept | -0.583 | *** | -0.577 | *** | 0.726 | ** |
|  | [-0.630 -0.537] |  | [-0.629 -0.525] |  | [0.194 1.258] |  |
| N | 158391 |  | 158391 |  | 158391 |  |
| *** p<.001, ** p<.01, * p<.05 | | |  |  |  |  |

| Estonia | | |  |  |  |  |
| --- | --- | --- | --- | --- | --- | --- |
| Adjustment | EUROD-D | | COND, ADL | | SES | |
| group_1 | 3.922 | *** | 3.922 | *** | 3.922 | *** |
|  | [3.908 3.936] |  | [3.908 3.936] |  | [3.908 3.936] |  |
| group_2 | 3.243 | *** | 3.243 | *** | 3.243 | *** |
|  | [3.230 3.256] |  | [3.230 3.256] |  | [3.230 3.256] |  |
| difference | 0.679 | *** | 0.679 | *** | 0.679 | *** |
|  | [0.660 0.698] |  | [0.660 0.698] |  | [0.660 0.698] |  |
| explained | 0.122 | *** | 0.173 | *** | 0.143 | *** |
|  | [0.111 0.133] |  | [0.160 0.186] |  | [0.123 0.163] |  |
| unexplained | 0.557 | *** | 0.506 | *** | 0.536 | *** |
|  | [0.540 0.574] |  | [0.490 0.522] |  | [0.512 0.560] |  |
| EUROD | 0.121 | *** | 0.094 | *** | 0.087 | *** |
|  | [0.110 0.131] |  | [0.085 0.102] |  | [0.079 0.095] |  |
| wave== 4.0000 | 0.000 |  | 0.000 |  | 0.000 |  |
|  | [-0.001 0.001] |  | [-0.001 0.001] |  | [-0.001 0.001] |  |
| wave== 6.0000 | 0.001 | ** | 0.000 |  | 0.000 | * |
|  | [0.000 0.001] |  | [-0.000 0.001] |  | [0.000 0.001] |  |
| ADL |  |  | 0.036 | *** | 0.031 | *** |
|  |  |  | [0.031 0.042] |  | [0.027 0.036] |  |
| COND |  |  | 0.042 | *** | 0.037 | *** |
|  |  |  | [0.037 0.048] |  | [0.032 0.042] |  |
| agegroup1 | |  |  |  | 0.008 | *** |
|  |  |  |  |  | [0.006 0.011] |  |
| agegroup3 | |  |  |  | 0.004 | *** |
|  |  |  |  |  | [0.002 0.005] |  |
| female |  |  |  |  | -0.004 | *** |
|  |  |  |  |  | [-0.007 -0.002] |  |
| partnerinhh== 3.0000 | | |  |  | 0.003 |  |
|  |  |  |  |  | [-0.001 0.006] |  |
| bieduc |  |  |  |  | -0.031 | *** |
|  |  |  |  |  | [-0.035 -0.027] |  |
| empl==employed or self-employed | | | |  | -0.008 | *** |
|  |  |  |  |  | [-0.010 -0.006] |  |
| empl==non employed | |  |  |  | 0.000 |  |
|  |  |  |  |  | [-0.001 0.001] |  |
| empl==other | |  |  |  | 0.005 | ** |
|  |  |  |  |  | [0.001 0.009] |  |
| RECODE of hin_adjlog (hin_adjlog) | | | |  | 0.010 |  |
|  |  |  |  |  | [-0.000 0.020] |  |
| housing==NA | |  |  |  | 0.000 |  |
|  |  |  |  |  | [-0.000 0.001] |  |
| EUROD | -0.163 | *** | -0.149 | *** | -0.151 | *** |
|  | [-0.185 -0.141] |  | [-0.172 -0.126] |  | [-0.175 -0.128] |  |
| wave== 4.0000 | -0.039 | *** | -0.041 | *** | -0.034 | *** |
|  | [-0.053 -0.025] |  | [-0.054 -0.028] |  | [-0.047 -0.021] |  |
| wave== 6.0000 | 0.017 | ** | 0.009 |  | 0.010 |  |
|  | [0.005 0.028] |  | [-0.002 0.020] |  | [-0.001 0.021] |  |
| ADL |  |  | -0.024 | *** | -0.024 | *** |
|  |  |  | [-0.032 -0.016] |  | [-0.032 -0.015] |  |
| COND |  |  | -0.091 | *** | -0.086 | *** |
|  |  |  | [-0.118 -0.063] |  | [-0.113 -0.058] |  |
| agegroup1 | |  |  |  | 0.007 |  |
|  |  |  |  |  | [-0.010 0.023] |  |
| agegroup3 | |  |  |  | 0.003 |  |
|  |  |  |  |  | [-0.005 0.011] |  |
| female |  |  |  |  | -0.032 | ** |
|  |  |  |  |  | [-0.054 -0.010] |  |
| partnerinhh== 3.0000 | | |  |  | -0.028 | ** |
|  |  |  |  |  | [-0.046 -0.010] |  |
| bieduc |  |  |  |  | 0.025 | *** |
|  |  |  |  |  | [0.013 0.038] |  |
| empl==employed or self-employed | | | |  | -0.009 |  |
|  |  |  |  |  | [-0.021 0.002] |  |
| empl==non employed | |  |  |  | -0.003 |  |
|  |  |  |  |  | [-0.008 0.002] |  |
| empl==other | |  |  |  | -0.002 | * |
|  |  |  |  |  | [-0.003 -0.000] |  |
| RECODE of hin_adjlog (hin_adjlog) | | | |  | -0.109 |  |
|  |  |  |  |  | [-0.317 0.099] |  |
| housing==NA | |  |  |  | 0.000 |  |
|  |  |  |  |  | [-0.001 0.000] |  |
| Intercept | 0.742 | *** | 0.802 | *** | 0.970 | *** |
|  | [0.708 0.776] |  | [0.762 0.842] |  | [0.746 1.194] |  |
| N | 165347 |  | 165347 |  | 165347 |  |
| *** p<.001, ** p<.01, * p<.05 | | |  |  |  |  |

| France | | |  |  |  |  |
| --- | --- | --- | --- | --- | --- | --- |
| Adjustment | EUROD-D | | COND, ADL | | SES | |
| group_1 | 3.245 | *** | 3.245 | *** | 3.245 | *** |
|  | [3.225 3.265] |  | [3.225 3.265] |  | [3.225 3.265] |  |
| group_2 | 3.243 | *** | 3.243 | *** | 3.243 | *** |
|  | [3.230 3.256] |  | [3.230 3.256] |  | [3.230 3.256] |  |
| difference | 0.002 |  | 0.002 |  | 0.002 |  |
|  | [-0.022 0.026] |  | [-0.022 0.026] |  | [-0.022 0.026] |  |
| explained | 0.056 | *** | 0.033 | *** | 0.031 | *** |
|  | [0.045 0.068] |  | [0.020 0.047] |  | [0.014 0.047] |  |
| unexplained | -0.055 | *** | -0.032 | ** | -0.029 | ** |
|  | [-0.076 -0.033] |  | [-0.052 -0.011] |  | [-0.049 -0.008] |  |
| EUROD | 0.054 | *** | 0.041 | *** | 0.039 | *** |
|  | [0.042 0.065] |  | [0.033 0.050] |  | [0.031 0.047] |  |
| wave== 4.0000 | 0.001 |  | 0.001 |  | 0.001 |  |
|  | [-0.000 0.002] |  | [-0.000 0.002] |  | [-0.000 0.002] |  |
| wave== 6.0000 | 0.002 | *** | 0.001 |  | 0.001 | * |
|  | [0.001 0.003] |  | [-0.000 0.002] |  | [0.000 0.002] |  |
| ADL |  |  | 0.009 | *** | 0.007 | *** |
|  |  |  | [0.004 0.013] |  | [0.003 0.011] |  |
| COND |  |  | -0.018 | *** | -0.016 | *** |
|  |  |  | [-0.025 -0.012] |  | [-0.022 -0.010] |  |
| agegroup1 | |  |  |  | -0.001 |  |
|  |  |  |  |  | [-0.002 0.000] |  |
| agegroup3 | |  |  |  | 0.003 | *** |
|  |  |  |  |  | [0.002 0.004] |  |
| female |  |  |  |  | 0.000 |  |
|  |  |  |  |  | [-0.001 0.000] |  |
| partnerinhh== 3.0000 | | |  |  | 0.000 |  |
|  |  |  |  |  | [-0.000 0.000] |  |
| bieduc |  |  |  |  | 0.003 | * |
|  |  |  |  |  | [0.000 0.005] |  |
| empl==employed or self-employed | | | |  | 0.003 | ** |
|  |  |  |  |  | [0.001 0.005] |  |
| empl==non employed | |  |  |  | -0.003 | *** |
|  |  |  |  |  | [-0.004 -0.001] |  |
| empl==other | |  |  |  | 0.003 | ** |
|  |  |  |  |  | [0.001 0.006] |  |
| RECODE of hin_adjlog (hin_adjlog) | | | |  | -0.009 | * |
|  |  |  |  |  | [-0.017 -0.001] |  |
| housing==NA | |  |  |  | 0.000 |  |
|  |  |  |  |  | [-0.000 0.000] |  |
| EUROD | -0.049 | *** | -0.031 | * | -0.025 |  |
|  | [-0.075 -0.024] |  | [-0.056 -0.005] |  | [-0.051 0.002] |  |
| wave== 4.0000 | -0.034 | *** | -0.038 | *** | -0.039 | *** |
|  | [-0.052 -0.017] |  | [-0.054 -0.021] |  | [-0.055 -0.023] |  |
| wave== 6.0000 | 0.006 |  | -0.001 |  | -0.003 |  |
|  | [-0.009 0.021] |  | [-0.014 0.013] |  | [-0.017 0.010] |  |
| ADL |  |  | 0.009 | * | 0.002 |  |
|  |  |  | [0.001 0.016] |  | [-0.006 0.010] |  |
| COND |  |  | -0.057 | *** | -0.065 | *** |
|  |  |  | [-0.085 -0.029] |  | [-0.093 -0.038] |  |
| agegroup1 | |  |  |  | -0.057 | *** |
|  |  |  |  |  | [-0.082 -0.032] |  |
| agegroup3 | |  |  |  | 0.011 | * |
|  |  |  |  |  | [0.002 0.021] |  |
| female |  |  |  |  | -0.013 |  |
|  |  |  |  |  | [-0.036 0.011] |  |
| partnerinhh== 3.0000 | | |  |  | -0.008 |  |
|  |  |  |  |  | [-0.023 0.008] |  |
| bieduc |  |  |  |  | -0.006 |  |
|  |  |  |  |  | [-0.016 0.005] |  |
| empl==employed or self-employed | | | |  | 0.025 | *** |
|  |  |  |  |  | [0.013 0.037] |  |
| empl==non employed | |  |  |  | 0.008 | ** |
|  |  |  |  |  | [0.002 0.013] |  |
| empl==other | |  |  |  | -0.001 |  |
|  |  |  |  |  | [-0.006 0.003] |  |
| RECODE of hin_adjlog (hin_adjlog) | | | |  | -0.472 | *** |
|  |  |  |  |  | [-0.716 -0.228] |  |
| housing==NA | |  |  |  | -0.001 |  |
|  |  |  |  |  | [-0.002 0.001] |  |
| Intercept | 0.024 |  | 0.086 | *** | 0.615 | *** |
|  | [-0.019 0.066] |  | [0.037 0.134] |  | [0.367 0.862] |  |
| N | 161856 |  | 161856 |  | 161856 |  |
| *** p<.001, ** p<.01, * p<.05 | | |  |  |  |  |

| Germany | | |  |  |  |  |
| --- | --- | --- | --- | --- | --- | --- |
| Adjustment | EUROD-D | | COND, ADL | | SES | |
| group_1 | 3.310 | *** | 3.310 | *** | 3.310 | *** |
|  | [3.288 3.332] |  | [3.288 3.332] |  | [3.288 3.332] |  |
| group_2 | 3.243 | *** | 3.243 | *** | 3.243 | *** |
|  | [3.230 3.256] |  | [3.230 3.256] |  | [3.230 3.256] |  |
| difference | 0.067 | *** | 0.067 | *** | 0.067 | *** |
|  | [0.042 0.093] |  | [0.042 0.093] |  | [0.042 0.093] |  |
| explained | -0.062 | *** | -0.029 | *** | -0.067 | *** |
|  | [-0.074 -0.051] |  | [-0.043 -0.014] |  | [-0.083 -0.051] |  |
| unexplained | 0.130 | *** | 0.096 | *** | 0.135 | *** |
|  | [0.107 0.152] |  | [0.075 0.117] |  | [0.114 0.155] |  |
| EUROD | -0.058 | *** | -0.045 | *** | -0.042 | *** |
|  | [-0.069 -0.047] |  | [-0.054 -0.036] |  | [-0.050 -0.034] |  |
| wave== 4.0000 | -0.005 | *** | -0.004 | *** | -0.003 | *** |
|  | [-0.007 -0.003] |  | [-0.006 -0.002] |  | [-0.005 -0.002] |  |
| wave== 6.0000 | 0.001 | * | 0.000 |  | 0.000 |  |
|  | [0.000 0.001] |  | [-0.000 0.000] |  | [-0.000 0.001] |  |
| ADL |  |  | 0.006 | * | 0.005 | * |
|  |  |  | [0.001 0.012] |  | [0.001 0.010] |  |
| COND |  |  | 0.014 | *** | 0.012 | *** |
|  |  |  | [0.007 0.021] |  | [0.006 0.018] |  |
| agegroup1 | |  |  |  | 0.000 |  |
|  |  |  |  |  | [-0.002 0.001] |  |
| agegroup3 | |  |  |  | -0.002 | ** |
|  |  |  |  |  | [-0.003 -0.001] |  |
| female |  |  |  |  | 0.001 | ** |
|  |  |  |  |  | [0.000 0.002] |  |
| partnerinhh== 3.0000 | | |  |  | 0.000 |  |
|  |  |  |  |  | [-0.000 0.000] |  |
| bieduc |  |  |  |  | -0.025 | *** |
|  |  |  |  |  | [-0.028 -0.021] |  |
| empl==employed or self-employed | | | |  | -0.010 | *** |
|  |  |  |  |  | [-0.013 -0.008] |  |
| empl==non employed | |  |  |  | 0.000 |  |
|  |  |  |  |  | [-0.002 0.001] |  |
| empl==other | |  |  |  | 0.003 | *** |
|  |  |  |  |  | [0.001 0.005] |  |
| RECODE of hin_adjlog (hin_adjlog) | | | |  | -0.007 | * |
|  |  |  |  |  | [-0.013 -0.001] |  |
| housing==NA | |  |  |  | 0.000 |  |
|  |  |  |  |  | [-0.001 0.000] |  |
| EUROD | -0.032 | * | -0.033 | ** | -0.027 | * |
|  | [-0.056 -0.007] |  | [-0.057 -0.008] |  | [-0.053 -0.002] |  |
| wave== 4.0000 | -0.030 | ** | -0.026 | ** | -0.026 | ** |
|  | [-0.049 -0.012] |  | [-0.043 -0.010] |  | [-0.042 -0.010] |  |
| wave== 6.0000 | 0.007 |  | 0.011 |  | 0.010 |  |
|  | [-0.009 0.022] |  | [-0.003 0.025] |  | [-0.004 0.024] |  |
| ADL |  |  | 0.010 | * | 0.007 |  |
|  |  |  | [0.002 0.019] |  | [-0.002 0.015] |  |
| COND |  |  | -0.037 | * | -0.036 | * |
|  |  |  | [-0.070 -0.004] |  | [-0.069 -0.003] |  |
| agegroup1 | |  |  |  | 0.007 |  |
|  |  |  |  |  | [-0.023 0.037] |  |
| agegroup3 | |  |  |  | 0.005 |  |
|  |  |  |  |  | [-0.003 0.013] |  |
| female |  |  |  |  | -0.027 | * |
|  |  |  |  |  | [-0.051 -0.004] |  |
| partnerinhh== 3.0000 | | |  |  | -0.006 |  |
|  |  |  |  |  | [-0.024 0.012] |  |
| bieduc |  |  |  |  | 0.012 |  |
|  |  |  |  |  | [-0.003 0.027] |  |
| empl==employed or self-employed | | | |  | 0.018 | * |
|  |  |  |  |  | [0.002 0.035] |  |
| empl==non employed | |  |  |  | 0.008 | * |
|  |  |  |  |  | [0.002 0.015] |  |
| empl==other | |  |  |  | -0.011 | *** |
|  |  |  |  |  | [-0.017 -0.005] |  |
| RECODE of hin_adjlog (hin_adjlog) | | | |  | -0.640 | *** |
|  |  |  |  |  | [-0.946 -0.334] |  |
| housing==NA | |  |  |  | 0.000 |  |
|  |  |  |  |  | [-0.003 0.003] |  |
| Intercept | 0.185 | *** | 0.170 | *** | 0.842 | *** |
|  | [0.143 0.226] |  | [0.121 0.220] |  | [0.532 1.152] |  |
| N | 159901 |  | 159901 |  | 159901 |  |
| *** p<.001, ** p<.01, * p<.05 | | |  |  |  |  |

| Greece | | |  |  |  |  |
| --- | --- | --- | --- | --- | --- | --- |
| Adjustment | EUROD-D | | COND, ADL | | SES | |
| group_1 | 3.004 | *** | 3.004 | *** | 3.004 | *** |
|  | [2.968 3.041] |  | [2.968 3.040] |  | [2.969 3.040] |  |
| group_2 | 3.243 | *** | 3.243 | *** | 3.243 | *** |
|  | [3.230 3.256] |  | [3.230 3.256] |  | [3.230 3.256] |  |
| difference | -0.239 | *** | -0.239 | *** | -0.239 | *** |
|  | [-0.277 -0.200] |  | [-0.277 -0.201] |  | [-0.277 -0.201] |  |
| explained | -0.059 | *** | -0.055 | *** | -0.050 | *** |
|  | [-0.083 -0.035] |  | [-0.080 -0.029] |  | [-0.077 -0.022] |  |
| unexplained | -0.180 | *** | -0.184 | *** | -0.189 | *** |
|  | [-0.217 -0.142] |  | [-0.217 -0.151] |  | [-0.222 -0.156] |  |
| EUROD | 0.006 |  | 0.005 |  | 0.004 |  |
|  | [-0.013 0.025] |  | [-0.010 0.020] |  | [-0.009 0.018] |  |
| wave== 4.0000 | -0.038 | *** | -0.033 | *** | -0.027 | *** |
|  | [-0.048 -0.028] |  | [-0.042 -0.024] |  | [-0.036 -0.018] |  |
| wave== 6.0000 | -0.027 | *** | -0.011 |  | -0.015 | * |
|  | [-0.041 -0.014] |  | [-0.023 0.002] |  | [-0.027 -0.002] |  |
| ADL |  |  | -0.027 | *** | -0.023 | *** |
|  |  |  | [-0.033 -0.021] |  | [-0.028 -0.018] |  |
| COND |  |  | 0.011 | * | 0.010 | * |
|  |  |  | [0.001 0.022] |  | [0.001 0.019] |  |
| agegroup1 | |  |  |  | -0.004 | *** |
|  |  |  |  |  | [-0.006 -0.002] |  |
| agegroup3 | |  |  |  | 0.000 |  |
|  |  |  |  |  | [-0.001 0.001] |  |
| female |  |  |  |  | 0.001 |  |
|  |  |  |  |  | [-0.000 0.001] |  |
| partnerinhh== 3.0000 | | |  |  | -0.001 |  |
|  |  |  |  |  | [-0.002 0.000] |  |
| bieduc |  |  |  |  | -0.003 |  |
|  |  |  |  |  | [-0.006 0.001] |  |
| empl==employed or self-employed | | | |  | 0.001 |  |
|  |  |  |  |  | [-0.003 0.005] |  |
| empl==non employed | |  |  |  | -0.001 |  |
|  |  |  |  |  | [-0.004 0.001] |  |
| empl==other | |  |  |  | -0.005 | ** |
|  |  |  |  |  | [-0.008 -0.001] |  |
| RECODE of hin_adjlog (hin_adjlog) | | | |  | 0.013 |  |
|  |  |  |  |  | [-0.001 0.026] |  |
| housing==NA | |  |  |  | 0.000 |  |
|  |  |  |  |  | [-0.001 0.001] |  |
| EUROD | -0.146 | *** | -0.153 | *** | -0.165 | *** |
|  | [-0.182 -0.109] |  | [-0.188 -0.117] |  | [-0.200 -0.129] |  |
| wave== 4.0000 | 0.000 | *** | 0.000 | * | 0.000 | ** |
|  | [0.000 0.000] |  | [0.000 0.000] |  | [0.000 0.000] |  |
| wave== 6.0000 | 0.041 | *** | 0.016 |  | 0.022 | * |
|  | [0.021 0.062] |  | [-0.002 0.035] |  | [0.003 0.042] |  |
| ADL |  |  | 0.012 | ** | 0.007 |  |
|  |  |  | [0.004 0.020] |  | [-0.001 0.015] |  |
| COND |  |  | 0.165 | *** | 0.110 | *** |
|  |  |  | [0.117 0.213] |  | [0.061 0.159] |  |
| agegroup1 | |  |  |  | -0.105 | *** |
|  |  |  |  |  | [-0.145 -0.065] |  |
| agegroup3 | |  |  |  | 0.010 |  |
|  |  |  |  |  | [-0.001 0.021] |  |
| female |  |  |  |  | 0.097 | *** |
|  |  |  |  |  | [0.058 0.135] |  |
| partnerinhh== 3.0000 | | |  |  | 0.019 |  |
|  |  |  |  |  | [-0.003 0.041] |  |
| bieduc |  |  |  |  | 0.014 |  |
|  |  |  |  |  | [-0.003 0.031] |  |
| empl==employed or self-employed | | | |  | -0.002 |  |
|  |  |  |  |  | [-0.022 0.018] |  |
| empl==non employed | |  |  |  | -0.004 |  |
|  |  |  |  |  | [-0.013 0.005] |  |
| empl==other | |  |  |  | -0.015 |  |
|  |  |  |  |  | [-0.035 0.005] |  |
| RECODE of hin_adjlog (hin_adjlog) | | | |  | 0.176 |  |
|  |  |  |  |  | [-0.034 0.386] |  |
| housing==NA | |  |  |  | -0.001 |  |
|  |  |  |  |  | [-0.001 0.000] |  |
| Intercept | -0.075 | ** | -0.225 | *** | -0.353 | ** |
|  | [-0.122 -0.029] |  | [-0.285 -0.164] |  | [-0.596 -0.110] |  |
| N | 154230 |  | 154230 |  | 154230 |  |
| *** p<.001, ** p<.01, * p<.05 | | |  |  |  |  |

| Hungary | | |  |  |  |  |
| --- | --- | --- | --- | --- | --- | --- |
| Adjustment | EUROD-D | | COND, ADL | | SES | |
| group_1 | 3.756 | *** | 3.756 | *** | 3.756 | *** |
|  | [3.661 3.851] |  | [3.662 3.850] |  | [3.664 3.848] |  |
| group_2 | 3.243 | *** | 3.243 | *** | 3.243 | *** |
|  | [3.230 3.256] |  | [3.230 3.256] |  | [3.230 3.256] |  |
| difference | 0.513 | *** | 0.513 | *** | 0.513 | *** |
|  | [0.417 0.609] |  | [0.418 0.608] |  | [0.420 0.606] |  |
| explained | 0.267 | *** | 0.292 | *** | 0.308 | *** |
|  | [0.213 0.322] |  | [0.235 0.350] |  | [0.248 0.367] |  |
| unexplained | 0.246 | *** | 0.220 | *** | 0.205 | *** |
|  | [0.159 0.332] |  | [0.139 0.302] |  | [0.124 0.286] |  |
| EUROD | 0.177 | *** | 0.137 | *** | 0.128 | *** |
|  | [0.126 0.228] |  | [0.097 0.176] |  | [0.091 0.165] |  |
| wave== 4.0000 | 0.077 | *** | 0.067 | *** | 0.055 | *** |
|  | [0.057 0.096] |  | [0.048 0.085] |  | [0.037 0.073] |  |
| wave== 6.0000 | 0.014 | *** | 0.006 |  | 0.008 | * |
|  | [0.007 0.021] |  | [-0.001 0.012] |  | [0.001 0.014] |  |
| ADL |  |  | 0.020 | * | 0.017 | * |
|  |  |  | [0.002 0.038] |  | [0.001 0.032] |  |
| COND |  |  | 0.063 | *** | 0.056 | *** |
|  |  |  | [0.044 0.083] |  | [0.039 0.073] |  |
| agegroup1 | |  |  |  | -0.003 |  |
|  |  |  |  |  | [-0.007 0.001] |  |
| agegroup3 | |  |  |  | -0.001 |  |
|  |  |  |  |  | [-0.004 0.001] |  |
| female |  |  |  |  | -0.002 | * |
|  |  |  |  |  | [-0.004 -0.000] |  |
| partnerinhh== 3.0000 | | |  |  | 0.002 |  |
|  |  |  |  |  | [-0.001 0.004] |  |
| bieduc |  |  |  |  | 0.008 | ** |
|  |  |  |  |  | [0.002 0.014] |  |
| empl==employed or self-employed | | | |  | 0.021 | *** |
|  |  |  |  |  | [0.014 0.027] |  |
| empl==non employed | |  |  |  | 0.009 | ** |
|  |  |  |  |  | [0.003 0.015] |  |
| empl==other | |  |  |  | 0.004 | ** |
|  |  |  |  |  | [0.001 0.007] |  |
| RECODE of hin_adjlog (hin_adjlog) | | | |  | 0.007 |  |
|  |  |  |  |  | [-0.001 0.016] |  |
| housing==NA | |  |  |  | 0.000 |  |
|  |  |  |  |  | [-0.001 0.000] |  |
| EUROD | -0.040 |  | -0.022 |  | -0.023 |  |
|  | [-0.128 0.048] |  | [-0.110 0.066] |  | [-0.117 0.072] |  |
| wave== 4.0000 | -0.114 | *** | -0.100 | *** | -0.082 | *** |
|  | [-0.144 -0.085] |  | [-0.128 -0.072] |  | [-0.109 -0.055] |  |
| wave== 6.0000 | 0.000 |  | 0.000 |  | 0.000 |  |
|  | [-0.000 0.000] |  | [-0.000 0.000] |  | [-0.000 0.000] |  |
| ADL |  |  | -0.038 |  | -0.025 |  |
|  |  |  | [-0.078 0.002] |  | [-0.056 0.007] |  |
| COND |  |  | 0.129 |  | 0.168 | * |
|  |  |  | [-0.027 0.286] |  | [0.024 0.313] |  |
| agegroup1 | |  |  |  | 0.077 |  |
|  |  |  |  |  | [-0.020 0.174] |  |
| agegroup3 | |  |  |  | -0.004 |  |
|  |  |  |  |  | [-0.043 0.035] |  |
| female |  |  |  |  | -0.050 |  |
|  |  |  |  |  | [-0.144 0.043] |  |
| partnerinhh== 3.0000 | | |  |  | -0.025 |  |
|  |  |  |  |  | [-0.097 0.048] |  |
| bieduc |  |  |  |  | -0.014 |  |
|  |  |  |  |  | [-0.045 0.017] |  |
| empl==employed or self-employed | | | |  | -0.027 |  |
|  |  |  |  |  | [-0.056 0.001] |  |
| empl==non employed | |  |  |  | -0.029 | * |
|  |  |  |  |  | [-0.054 -0.004] |  |
| empl==other | |  |  |  | 0.006 |  |
|  |  |  |  |  | [-0.009 0.022] |  |
| RECODE of hin_adjlog (hin_adjlog) | | | |  | 1.090 |  |
|  |  |  |  |  | [-0.127 2.307] |  |
| housing==NA | |  |  |  | -0.001 |  |
|  |  |  |  |  | [-0.007 0.005] |  |
| Intercept | 0.400 | *** | 0.251 | ** | -0.857 |  |
|  | [0.262 0.538] |  | [0.061 0.440] |  | [-2.097 0.384] |  |
| N | 152924 |  | 152924 |  | 152924 |  |
| *** p<.001, ** p<.01, * p<.05 | | |  |  |  |  |

| Israel | | |  |  |  |  |
| --- | --- | --- | --- | --- | --- | --- |
| Adjustment | EUROD-D | | COND, ADL | | SES | |
| group_1 | 3.027 | *** | 3.027 | *** | 3.027 | *** |
|  | [2.977 3.077] |  | [2.977 3.077] |  | [2.978 3.077] |  |
| group_2 | 3.243 | *** | 3.243 | *** | 3.243 | *** |
|  | [3.230 3.256] |  | [3.230 3.256] |  | [3.230 3.256] |  |
| difference | -0.216 | *** | -0.216 | *** | -0.216 | *** |
|  | [-0.267 -0.164] |  | [-0.267 -0.164] |  | [-0.267 -0.164] |  |
| explained | -0.114 | *** | -0.099 | *** | -0.149 | *** |
|  | [-0.139 -0.090] |  | [-0.129 -0.070] |  | [-0.181 -0.118] |  |
| unexplained | -0.101 | *** | -0.117 | *** | -0.066 | ** |
|  | [-0.147 -0.056] |  | [-0.158 -0.075] |  | [-0.107 -0.026] |  |
| EUROD | -0.072 | *** | -0.056 | *** | -0.052 | *** |
|  | [-0.094 -0.050] |  | [-0.073 -0.038] |  | [-0.068 -0.036] |  |
| wave== 4.0000 | -0.038 | *** | -0.033 | *** | -0.027 | *** |
|  | [-0.047 -0.028] |  | [-0.042 -0.024] |  | [-0.036 -0.018] |  |
| wave== 6.0000 | -0.005 | *** | -0.002 |  | -0.003 | * |
|  | [-0.007 -0.002] |  | [-0.004 0.000] |  | [-0.005 -0.000] |  |
| ADL |  |  | -0.007 |  | -0.006 |  |
|  |  |  | [-0.015 0.000] |  | [-0.013 0.000] |  |
| COND |  |  | -0.001 |  | -0.001 |  |
|  |  |  | [-0.014 0.012] |  | [-0.013 0.010] |  |
| agegroup1 | |  |  |  | 0.002 | * |
|  |  |  |  |  | [0.000 0.005] |  |
| agegroup3 | |  |  |  | -0.001 |  |
|  |  |  |  |  | [-0.002 0.001] |  |
| female |  |  |  |  | 0.000 |  |
|  |  |  |  |  | [-0.001 0.000] |  |
| partnerinhh== 3.0000 | | |  |  | -0.001 |  |
|  |  |  |  |  | [-0.003 0.000] |  |
| bieduc |  |  |  |  | -0.038 | *** |
|  |  |  |  |  | [-0.044 -0.031] |  |
| empl==employed or self-employed | | | |  | -0.022 | *** |
|  |  |  |  |  | [-0.028 -0.017] |  |
| empl==non employed | |  |  |  | 0.001 |  |
|  |  |  |  |  | [-0.002 0.003] |  |
| empl==other | |  |  |  | 0.000 |  |
|  |  |  |  |  | [-0.001 0.001] |  |
| RECODE of hin_adjlog (hin_adjlog) | | | |  | -0.001 |  |
|  |  |  |  |  | [-0.003 0.001] |  |
| housing==NA | |  |  |  | 0.000 |  |
|  |  |  |  |  | [-0.000 0.000] |  |
| EUROD | 0.048 | * | -0.024 |  | -0.058 | ** |
|  | [0.011 0.084] |  | [-0.066 0.018] |  | [-0.099 -0.018] |  |
| wave== 4.0000 | 0.000 |  | 0.000 |  | 0.000 |  |
|  | [-0.000 0.000] |  | [-0.000 0.000] |  | [-0.000 0.000] |  |
| wave== 6.0000 | -0.017 |  | -0.011 |  | -0.029 |  |
|  | [-0.057 0.024] |  | [-0.049 0.026] |  | [-0.066 0.007] |  |
| ADL |  |  | 0.027 | *** | 0.019 | ** |
|  |  |  | [0.013 0.040] |  | [0.006 0.032] |  |
| COND |  |  | 0.087 | ** | 0.078 | ** |
|  |  |  | [0.027 0.147] |  | [0.019 0.137] |  |
| agegroup1 | |  |  |  | -0.040 |  |
|  |  |  |  |  | [-0.086 0.006] |  |
| agegroup3 | |  |  |  | 0.021 | * |
|  |  |  |  |  | [0.005 0.037] |  |
| female |  |  |  |  | -0.017 |  |
|  |  |  |  |  | [-0.065 0.030] |  |
| partnerinhh== 3.0000 | | |  |  | -0.003 |  |
|  |  |  |  |  | [-0.032 0.026] |  |
| bieduc |  |  |  |  | 0.038 | * |
|  |  |  |  |  | [0.005 0.071] |  |
| empl==employed or self-employed | | | |  | -0.007 |  |
|  |  |  |  |  | [-0.040 0.025] |  |
| empl==non employed | |  |  |  | 0.022 | *** |
|  |  |  |  |  | [0.010 0.035] |  |
| empl==other | |  |  |  | 0.015 |  |
|  |  |  |  |  | [-0.003 0.034] |  |
| RECODE of hin_adjlog (hin_adjlog) | | | |  | -0.103 |  |
|  |  |  |  |  | [-0.437 0.230] |  |
| housing==NA | |  |  |  | 0.000 |  |
|  |  |  |  |  | [-0.003 0.003] |  |
| Intercept | -0.132 | *** | -0.196 | *** | -0.001 |  |
|  | [-0.207 -0.057] |  | [-0.277 -0.114] |  | [-0.366 0.363] |  |
| N | 153832 |  | 153832 |  | 153832 |  |
| *** p<.001, ** p<.01, * p<.05 | | |  |  |  |  |

| Italy | | |  |  |  |  |
| --- | --- | --- | --- | --- | --- | --- |
| Adjustment | EUROD-D | | COND, ADL | | SES | |
| group_1 | 3.273 | *** | 3.273 | *** | 3.273 | *** |
|  | [3.248 3.298] |  | [3.249 3.297] |  | [3.249 3.297] |  |
| group_2 | 3.243 | *** | 3.243 | *** | 3.243 | *** |
|  | [3.230 3.256] |  | [3.230 3.256] |  | [3.230 3.256] |  |
| difference | 0.030 | * | 0.030 | * | 0.030 | * |
|  | [0.002 0.058] |  | [0.002 0.057] |  | [0.002 0.058] |  |
| explained | 0.044 | *** | 0.013 |  | 0.033 | *** |
|  | [0.031 0.058] |  | [-0.003 0.029] |  | [0.016 0.050] |  |
| unexplained | -0.014 |  | 0.017 |  | -0.003 |  |
|  | [-0.039 0.010] |  | [-0.006 0.040] |  | [-0.026 0.020] |  |
| EUROD | 0.055 | *** | 0.042 | *** | 0.039 | *** |
|  | [0.042 0.068] |  | [0.032 0.053] |  | [0.030 0.049] |  |
| wave== 4.0000 | -0.009 | *** | -0.008 | *** | -0.006 | *** |
|  | [-0.011 -0.006] |  | [-0.010 -0.005] |  | [-0.008 -0.004] |  |
| wave== 6.0000 | -0.002 | *** | -0.001 |  | -0.001 | * |
|  | [-0.003 -0.001] |  | [-0.001 0.000] |  | [-0.002 -0.000] |  |
| ADL |  |  | -0.005 | * | -0.004 | * |
|  |  |  | [-0.009 -0.000] |  | [-0.008 -0.000] |  |
| COND |  |  | -0.017 | *** | -0.015 | *** |
|  |  |  | [-0.024 -0.009] |  | [-0.021 -0.008] |  |
| agegroup1 | |  |  |  | -0.001 |  |
|  |  |  |  |  | [-0.002 0.000] |  |
| agegroup3 | |  |  |  | -0.002 | *** |
|  |  |  |  |  | [-0.003 -0.001] |  |
| female |  |  |  |  | 0.000 |  |
|  |  |  |  |  | [-0.000 0.001] |  |
| partnerinhh== 3.0000 | | |  |  | -0.001 |  |
|  |  |  |  |  | [-0.001 0.000] |  |
| bieduc |  |  |  |  | 0.026 | *** |
|  |  |  |  |  | [0.022 0.029] |  |
| empl==employed or self-employed | | | |  | -0.001 |  |
|  |  |  |  |  | [-0.003 0.002] |  |
| empl==non employed | |  |  |  | -0.004 | *** |
|  |  |  |  |  | [-0.005 -0.002] |  |
| empl==other | |  |  |  | -0.002 | * |
|  |  |  |  |  | [-0.004 -0.000] |  |
| RECODE of hin_adjlog (hin_adjlog) | | | |  | 0.002 |  |
|  |  |  |  |  | [-0.001 0.006] |  |
| housing==NA | |  |  |  | 0.000 |  |
|  |  |  |  |  | [-0.000 0.001] |  |
| EUROD | -0.037 | ** | -0.032 | * | -0.035 | * |
|  | [-0.065 -0.010] |  | [-0.060 -0.004] |  | [-0.064 -0.006] |  |
| wave== 4.0000 | -0.024 | ** | -0.021 | ** | -0.023 | ** |
|  | [-0.039 -0.008] |  | [-0.036 -0.006] |  | [-0.037 -0.008] |  |
| wave== 6.0000 | -0.008 |  | -0.002 |  | 0.000 |  |
|  | [-0.029 0.014] |  | [-0.022 0.018] |  | [-0.020 0.019] |  |
| ADL |  |  | -0.010 | * | -0.012 | ** |
|  |  |  | [-0.018 -0.002] |  | [-0.020 -0.004] |  |
| COND |  |  | -0.024 |  | -0.025 |  |
|  |  |  | [-0.057 0.009] |  | [-0.058 0.008] |  |
| agegroup1 | |  |  |  | -0.041 | ** |
|  |  |  |  |  | [-0.068 -0.014] |  |
| agegroup3 | |  |  |  | 0.005 |  |
|  |  |  |  |  | [-0.003 0.013] |  |
| female |  |  |  |  | 0.040 | ** |
|  |  |  |  |  | [0.012 0.068] |  |
| partnerinhh== 3.0000 | | |  |  | 0.013 |  |
|  |  |  |  |  | [-0.006 0.032] |  |
| bieduc |  |  |  |  | 0.004 |  |
|  |  |  |  |  | [-0.005 0.013] |  |
| empl==employed or self-employed | | | |  | 0.016 |  |
|  |  |  |  |  | [-0.000 0.032] |  |
| empl==non employed | |  |  |  | -0.002 |  |
|  |  |  |  |  | [-0.008 0.005] |  |
| empl==other | |  |  |  | 0.011 |  |
|  |  |  |  |  | [-0.000 0.023] |  |
| RECODE of hin_adjlog (hin_adjlog) | | | |  | 0.368 | ** |
|  |  |  |  |  | [0.110 0.625] |  |
| housing==NA | |  |  |  | 0.000 |  |
|  |  |  |  |  | [-0.001 0.001] |  |
| Intercept | 0.054 | * | 0.106 | *** | -0.322 | * |
|  | [0.002 0.107] |  | [0.051 0.161] |  | [-0.592 -0.051] |  |
| N | 161178 |  | 161178 |  | 161178 |  |
| *** p<.001, ** p<.01, * p<.05 | | |  |  |  |  |

| Luxembourg | | |  |  |  |  |
| --- | --- | --- | --- | --- | --- | --- |
| Adjustment | EUROD-D | | COND, ADL | | SES | |
| group_1 | 3.114 | *** | 3.114 | *** | 3.114 | *** |
|  | [3.071 3.157] |  | [3.071 3.157] |  | [3.071 3.157] |  |
| group_2 | 3.243 | *** | 3.243 | *** | 3.243 | *** |
|  | [3.230 3.256] |  | [3.230 3.256] |  | [3.230 3.256] |  |
| difference | -0.129 | *** | -0.129 | *** | -0.129 | *** |
|  | [-0.174 -0.085] |  | [-0.174 -0.085] |  | [-0.174 -0.084] |  |
| explained | -0.045 | *** | -0.015 |  | -0.026 |  |
|  | [-0.067 -0.023] |  | [-0.041 0.011] |  | [-0.059 0.007] |  |
| unexplained | -0.084 | *** | -0.115 | *** | -0.104 | *** |
|  | [-0.124 -0.044] |  | [-0.152 -0.077] |  | [-0.144 -0.063] |  |
| EUROD | -0.004 |  | -0.003 |  | -0.003 |  |
|  | [-0.023 0.016] |  | [-0.018 0.012] |  | [-0.017 0.012] |  |
| wave== 4.0000 | -0.038 | *** | -0.033 | *** | -0.027 | *** |
|  | [-0.048 -0.028] |  | [-0.042 -0.024] |  | [-0.036 -0.018] |  |
| wave== 6.0000 | -0.004 | *** | -0.001 |  | -0.002 | * |
|  | [-0.006 -0.002] |  | [-0.003 0.000] |  | [-0.004 -0.000] |  |
| ADL |  |  | 0.003 |  | 0.003 |  |
|  |  |  | [-0.005 0.012] |  | [-0.004 0.010] |  |
| COND |  |  | 0.019 | *** | 0.017 | *** |
|  |  |  | [0.008 0.031] |  | [0.007 0.027] |  |
| agegroup1 | |  |  |  | -0.005 | *** |
|  |  |  |  |  | [-0.008 -0.003] |  |
| agegroup3 | |  |  |  | 0.000 |  |
|  |  |  |  |  | [-0.002 0.001] |  |
| female |  |  |  |  | 0.000 |  |
|  |  |  |  |  | [-0.000 0.001] |  |
| partnerinhh== 3.0000 | | |  |  | 0.000 |  |
|  |  |  |  |  | [-0.000 0.001] |  |
| bieduc |  |  |  |  | -0.002 |  |
|  |  |  |  |  | [-0.005 0.002] |  |
| empl==employed or self-employed | | | |  | 0.011 | *** |
|  |  |  |  |  | [0.007 0.015] |  |
| empl==non employed | |  |  |  | 0.001 |  |
|  |  |  |  |  | [-0.002 0.003] |  |
| empl==other | |  |  |  | -0.003 | * |
|  |  |  |  |  | [-0.005 -0.001] |  |
| RECODE of hin_adjlog (hin_adjlog) | | | |  | -0.016 |  |
|  |  |  |  |  | [-0.033 0.001] |  |
| housing==NA | |  |  |  | -0.001 |  |
|  |  |  |  |  | [-0.003 0.002] |  |
| EUROD | 0.021 |  | 0.021 |  | 0.030 |  |
|  | [-0.023 0.064] |  | [-0.024 0.067] |  | [-0.017 0.077] |  |
| wave== 4.0000 | 0.000 |  | 0.000 |  | 0.000 |  |
|  | [-0.000 0.000] |  | [-0.000 0.000] |  | [-0.000 0.000] |  |
| wave== 6.0000 | 0.013 |  | 0.023 |  | 0.025 |  |
|  | [-0.021 0.047] |  | [-0.009 0.055] |  | [-0.007 0.057] |  |
| ADL |  |  | 0.010 |  | 0.014 |  |
|  |  |  | [-0.004 0.025] |  | [-0.000 0.028] |  |
| COND |  |  | -0.069 | * | -0.078 | * |
|  |  |  | [-0.129 -0.009] |  | [-0.138 -0.018] |  |
| agegroup1 | |  |  |  | -0.024 |  |
|  |  |  |  |  | [-0.076 0.027] |  |
| agegroup3 | |  |  |  | -0.020 | * |
|  |  |  |  |  | [-0.037 -0.003] |  |
| female |  |  |  |  | -0.063 | ** |
|  |  |  |  |  | [-0.109 -0.016] |  |
| partnerinhh== 3.0000 | | |  |  | 0.022 |  |
|  |  |  |  |  | [-0.012 0.056] |  |
| bieduc |  |  |  |  | -0.021 |  |
|  |  |  |  |  | [-0.043 0.000] |  |
| empl==employed or self-employed | | | |  | 0.021 | * |
|  |  |  |  |  | [0.002 0.040] |  |
| empl==non employed | |  |  |  | 0.013 | * |
|  |  |  |  |  | [0.002 0.025] |  |
| empl==other | |  |  |  | 0.019 |  |
|  |  |  |  |  | [-0.002 0.040] |  |
| RECODE of hin_adjlog (hin_adjlog) | | | |  | -0.046 |  |
|  |  |  |  |  | [-0.473 0.381] |  |
| housing==NA | |  |  |  | 0.001 |  |
|  |  |  |  |  | [-0.007 0.009] |  |
| Intercept | -0.118 | ** | -0.100 | * | 0.003 |  |
|  | [-0.191 -0.045] |  | [-0.190 -0.010] |  | [-0.445 0.451] |  |
| N | 153006 |  | 153006 |  | 153006 |  |
| *** p<.001, ** p<.01, * p<.05 | | |  |  |  |  |

| Netherlands | | |  |  |  |  |
| --- | --- | --- | --- | --- | --- | --- |
| Adjustment | EUROD-D | | COND, ADL | | SES | |
| group_1 | 2.976 | *** | 2.976 | *** | 2.976 | *** |
|  | [2.947 3.005] |  | [2.947 3.005] |  | [2.947 3.005] |  |
| group_2 | 3.243 | *** | 3.243 | *** | 3.243 | *** |
|  | [3.230 3.256] |  | [3.230 3.256] |  | [3.230 3.256] |  |
| difference | -0.267 | *** | -0.267 | *** | -0.267 | *** |
|  | [-0.298 -0.235] |  | [-0.298 -0.235] |  | [-0.299 -0.235] |  |
| explained | -0.116 | *** | -0.169 | *** | -0.174 | *** |
|  | [-0.131 -0.101] |  | [-0.187 -0.152] |  | [-0.193 -0.154] |  |
| unexplained | -0.151 | *** | -0.098 | *** | -0.093 | *** |
|  | [-0.181 -0.121] |  | [-0.126 -0.070] |  | [-0.120 -0.065] |  |
| EUROD | -0.136 | *** | -0.105 | *** | -0.098 | *** |
|  | [-0.149 -0.123] |  | [-0.116 -0.095] |  | [-0.108 -0.088] |  |
| wave== 4.0000 | 0.005 | *** | 0.005 | *** | 0.004 | *** |
|  | [0.003 0.007] |  | [0.003 0.007] |  | [0.002 0.005] |  |
| wave== 6.0000 | 0.015 | *** | 0.006 |  | 0.008 | * |
|  | [0.008 0.021] |  | [-0.000 0.012] |  | [0.002 0.015] |  |
| ADL |  |  | -0.021 | *** | -0.018 | *** |
|  |  |  | [-0.026 -0.015] |  | [-0.023 -0.013] |  |
| COND |  |  | -0.054 | *** | -0.048 | *** |
|  |  |  | [-0.063 -0.045] |  | [-0.056 -0.040] |  |
| agegroup1 | |  |  |  | -0.002 | ** |
|  |  |  |  |  | [-0.004 -0.001] |  |
| agegroup3 | |  |  |  | 0.000 |  |
|  |  |  |  |  | [-0.001 0.001] |  |
| female |  |  |  |  | 0.000 |  |
|  |  |  |  |  | [-0.001 0.001] |  |
| partnerinhh== 3.0000 | | |  |  | 0.000 |  |
|  |  |  |  |  | [-0.000 0.000] |  |
| bieduc |  |  |  |  | -0.008 | *** |
|  |  |  |  |  | [-0.011 -0.005] |  |
| empl==employed or self-employed | | | |  | -0.005 | *** |
|  |  |  |  |  | [-0.008 -0.002] |  |
| empl==non employed | |  |  |  | 0.003 | ** |
|  |  |  |  |  | [0.001 0.005] |  |
| empl==other | |  |  |  | -0.001 | * |
|  |  |  |  |  | [-0.002 -0.000] |  |
| RECODE of hin_adjlog (hin_adjlog) | | | |  | -0.008 |  |
|  |  |  |  |  | [-0.017 0.000] |  |
| housing==NA | |  |  |  | 0.000 |  |
|  |  |  |  |  | [-0.001 0.001] |  |
| EUROD | -0.006 |  | 0.007 |  | 0.009 |  |
|  | [-0.032 0.021] |  | [-0.020 0.033] |  | [-0.018 0.036] |  |
| wave== 4.0000 | -0.045 | *** | -0.040 | *** | -0.033 | ** |
|  | [-0.068 -0.021] |  | [-0.061 -0.018] |  | [-0.054 -0.011] |  |
| wave== 6.0000 | -0.001 | *** | -0.001 | *** | -0.001 | *** |
|  | [-0.001 -0.000] |  | [-0.001 -0.000] |  | [-0.001 -0.000] |  |
| ADL |  |  | 0.010 | * | 0.009 | * |
|  |  |  | [0.002 0.017] |  | [0.001 0.017] |  |
| COND |  |  | -0.012 |  | -0.008 |  |
|  |  |  | [-0.044 0.021] |  | [-0.040 0.025] |  |
| agegroup1 | |  |  |  | -0.011 |  |
|  |  |  |  |  | [-0.049 0.028] |  |
| agegroup3 | |  |  |  | -0.008 |  |
|  |  |  |  |  | [-0.020 0.005] |  |
| female |  |  |  |  | -0.002 |  |
|  |  |  |  |  | [-0.034 0.031] |  |
| partnerinhh== 3.0000 | | |  |  | -0.012 |  |
|  |  |  |  |  | [-0.036 0.012] |  |
| bieduc |  |  |  |  | 0.016 |  |
|  |  |  |  |  | [-0.001 0.032] |  |
| empl==employed or self-employed | | | |  | 0.027 | ** |
|  |  |  |  |  | [0.007 0.047] |  |
| empl==non employed | |  |  |  | 0.013 | ** |
|  |  |  |  |  | [0.003 0.022] |  |
| empl==other | |  |  |  | -0.001 |  |
|  |  |  |  |  | [-0.013 0.012] |  |
| RECODE of hin_adjlog (hin_adjlog) | | | |  | -0.457 |  |
|  |  |  |  |  | [-0.927 0.014] |  |
| housing==NA | |  |  |  | 0.000 |  |
|  |  |  |  |  | [-0.004 0.004] |  |
| Intercept | -0.100 | *** | -0.062 | * | 0.364 |  |
|  | [-0.146 -0.053] |  | [-0.115 -0.009] |  | [-0.109 0.837] |  |
| N | 156097 |  | 156097 |  | 156097 |  |
| *** p<.001, ** p<.01, * p<.05 | | |  |  |  |  |

| Slovenia | | |  |  |  |  |
| --- | --- | --- | --- | --- | --- | --- |
| Adjustment | EUROD-D | | COND, ADL | | SES | |
| group_1 | 3.360 | *** | 3.360 | *** | 3.360 | *** |
|  | [3.328 3.391] |  | [3.329 3.390] |  | [3.330 3.389] |  |
| group_2 | 3.243 | *** | 3.243 | *** | 3.243 | *** |
|  | [3.230 3.256] |  | [3.230 3.256] |  | [3.230 3.256] |  |
| difference | 0.117 | *** | 0.117 | *** | 0.117 | *** |
|  | [0.083 0.150] |  | [0.083 0.150] |  | [0.084 0.149] |  |
| explained | -0.016 | * | 0.006 |  | 0.046 | *** |
|  | [-0.029 -0.002] |  | [-0.010 0.023] |  | [0.028 0.064] |  |
| unexplained | 0.132 | *** | 0.110 | *** | 0.070 | *** |
|  | [0.101 0.164] |  | [0.081 0.140] |  | [0.040 0.100] |  |
| EUROD | -0.020 | ** | -0.015 | ** | -0.014 | ** |
|  | [-0.033 -0.007] |  | [-0.025 -0.005] |  | [-0.023 -0.005] |  |
| wave== 4.0000 | 0.003 | ** | 0.003 | ** | 0.002 | ** |
|  | [0.001 0.005] |  | [0.001 0.005] |  | [0.001 0.004] |  |
| wave== 6.0000 | 0.001 | * | 0.000 |  | 0.000 |  |
|  | [0.000 0.001] |  | [-0.000 0.001] |  | [-0.000 0.001] |  |
| ADL |  |  | -0.004 |  | -0.003 |  |
|  |  |  | [-0.010 0.002] |  | [-0.008 0.002] |  |
| COND |  |  | 0.022 | *** | 0.019 | *** |
|  |  |  | [0.014 0.030] |  | [0.012 0.027] |  |
| agegroup1 | |  |  |  | 0.003 | *** |
|  |  |  |  |  | [0.001 0.005] |  |
| agegroup3 | |  |  |  | 0.003 | *** |
|  |  |  |  |  | [0.001 0.004] |  |
| female |  |  |  |  | -0.002 | ** |
|  |  |  |  |  | [-0.003 -0.001] |  |
| partnerinhh== 3.0000 | | |  |  | 0.001 |  |
|  |  |  |  |  | [-0.001 0.003] |  |
| bieduc |  |  |  |  | 0.008 | *** |
|  |  |  |  |  | [0.005 0.011] |  |
| empl==employed or self-employed | | | |  | 0.026 | *** |
|  |  |  |  |  | [0.022 0.031] |  |
| empl==non employed | |  |  |  | -0.004 | *** |
|  |  |  |  |  | [-0.006 -0.002] |  |
| empl==other | |  |  |  | 0.002 | ** |
|  |  |  |  |  | [0.001 0.004] |  |
| RECODE of hin_adjlog (hin_adjlog) | | | |  | 0.004 |  |
|  |  |  |  |  | [-0.000 0.008] |  |
| housing==NA | |  |  |  | 0.000 |  |
|  |  |  |  |  | [-0.000 0.001] |  |
| EUROD | -0.036 | * | -0.026 |  | -0.046 | ** |
|  | [-0.068 -0.005] |  | [-0.058 0.005] |  | [-0.079 -0.014] |  |
| wave== 4.0000 | -0.024 |  | -0.021 |  | 0.002 |  |
|  | [-0.054 0.007] |  | [-0.049 0.008] |  | [-0.025 0.029] |  |
| wave== 6.0000 | 0.045 | *** | 0.028 | *** | 0.032 | *** |
|  | [0.028 0.062] |  | [0.012 0.044] |  | [0.016 0.048] |  |
| ADL |  |  | 0.002 |  | 0.004 |  |
|  |  |  | [-0.008 0.012] |  | [-0.006 0.014] |  |
| COND |  |  | -0.063 | ** | -0.069 | ** |
|  |  |  | [-0.106 -0.021] |  | [-0.111 -0.027] |  |
| agegroup1 | |  |  |  | -0.072 | *** |
|  |  |  |  |  | [-0.108 -0.036] |  |
| agegroup3 | |  |  |  | -0.007 |  |
|  |  |  |  |  | [-0.021 0.006] |  |
| female |  |  |  |  | 0.006 |  |
|  |  |  |  |  | [-0.034 0.047] |  |
| partnerinhh== 3.0000 | | |  |  | -0.028 |  |
|  |  |  |  |  | [-0.059 0.003] |  |
| bieduc |  |  |  |  | -0.017 | ** |
|  |  |  |  |  | [-0.030 -0.005] |  |
| empl==employed or self-employed | | | |  | 0.007 |  |
|  |  |  |  |  | [-0.001 0.015] |  |
| empl==non employed | |  |  |  | -0.002 |  |
|  |  |  |  |  | [-0.008 0.003] |  |
| empl==other | |  |  |  | 0.009 |  |
|  |  |  |  |  | [-0.000 0.019] |  |
| RECODE of hin_adjlog (hin_adjlog) | | | |  | -0.681 | *** |
|  |  |  |  |  | [-0.945 -0.418] |  |
| housing==NA | |  |  |  | 0.000 |  |
|  |  |  |  |  | [-0.000 0.001] |  |
| Intercept | 0.147 | *** | 0.191 | *** | 0.932 | *** |
|  | [0.097 0.198] |  | [0.130 0.251] |  | [0.651 1.213] |  |
| N | 158707 |  | 158707 |  | 158707 |  |
| *** p<.001, ** p<.01, * p<.05 | | |  |  |  |  |

| Spain | | |  |  |  |  |
| --- | --- | --- | --- | --- | --- | --- |
| Adjustment | EUROD-D | | COND, ADL | | SES | |
| group_1 | 3.372 | *** | 3.372 | *** | 3.372 | *** |
|  | [3.337 3.407] |  | [3.338 3.406] |  | [3.337 3.406] |  |
| group_2 | 3.243 | *** | 3.243 | *** | 3.243 | *** |
|  | [3.230 3.256] |  | [3.230 3.256] |  | [3.230 3.256] |  |
| difference | 0.129 | *** | 0.129 | *** | 0.129 | *** |
|  | [0.092 0.166] |  | [0.092 0.165] |  | [0.092 0.166] |  |
| explained | 0.024 | ** | 0.047 | *** | 0.096 | *** |
|  | [0.006 0.042] |  | [0.025 0.070] |  | [0.072 0.120] |  |
| unexplained | 0.104 | *** | 0.081 | *** | 0.033 | * |
|  | [0.072 0.137] |  | [0.052 0.111] |  | [0.004 0.062] |  |
| EUROD | 0.033 | *** | 0.025 | *** | 0.024 | *** |
|  | [0.015 0.050] |  | [0.012 0.039] |  | [0.011 0.036] |  |
| wave== 4.0000 | -0.006 | *** | -0.006 | *** | -0.005 | *** |
|  | [-0.008 -0.004] |  | [-0.008 -0.004] |  | [-0.006 -0.003] |  |
| wave== 6.0000 | -0.002 | *** | -0.001 | * | -0.001 | * |
|  | [-0.003 -0.001] |  | [-0.002 -0.000] |  | [-0.002 -0.000] |  |
| ADL |  |  | 0.009 | ** | 0.008 | ** |
|  |  |  | [0.003 0.015] |  | [0.003 0.013] |  |
| COND |  |  | 0.019 | *** | 0.017 | *** |
|  |  |  | [0.010 0.029] |  | [0.008 0.026] |  |
| agegroup1 | |  |  |  | 0.005 | *** |
|  |  |  |  |  | [0.003 0.008] |  |
| agegroup3 | |  |  |  | 0.005 | *** |
|  |  |  |  |  | [0.003 0.007] |  |
| female |  |  |  |  | 0.000 |  |
|  |  |  |  |  | [-0.001 0.001] |  |
| partnerinhh== 3.0000 | | |  |  | 0.000 |  |
|  |  |  |  |  | [-0.000 0.000] |  |
| bieduc |  |  |  |  | 0.027 | *** |
|  |  |  |  |  | [0.023 0.030] |  |
| empl==employed or self-employed | | | |  | 0.010 | *** |
|  |  |  |  |  | [0.006 0.014] |  |
| empl==non employed | |  |  |  | 0.006 | *** |
|  |  |  |  |  | [0.003 0.009] |  |
| empl==other | |  |  |  | -0.005 | * |
|  |  |  |  |  | [-0.009 -0.000] |  |
| RECODE of hin_adjlog (hin_adjlog) | | | |  | 0.005 |  |
|  |  |  |  |  | [-0.000 0.010] |  |
| housing==NA | |  |  |  | 0.000 |  |
|  |  |  |  |  | [-0.000 0.001] |  |
| EUROD | -0.012 |  | -0.006 |  | -0.006 |  |
|  | [-0.045 0.022] |  | [-0.040 0.028] |  | [-0.040 0.029] |  |
| wave== 4.0000 | -0.025 | ** | -0.020 | * | -0.017 | * |
|  | [-0.043 -0.006] |  | [-0.037 -0.003] |  | [-0.033 -0.001] |  |
| wave== 6.0000 | -0.032 | * | -0.026 |  | -0.015 |  |
|  | [-0.062 -0.001] |  | [-0.053 0.001] |  | [-0.041 0.011] |  |
| ADL |  |  | -0.011 | * | -0.006 |  |
|  |  |  | [-0.020 -0.001] |  | [-0.016 0.003] |  |
| COND |  |  | -0.021 |  | -0.027 |  |
|  |  |  | [-0.075 0.032] |  | [-0.077 0.022] |  |
| agegroup1 | |  |  |  | -0.024 |  |
|  |  |  |  |  | [-0.055 0.008] |  |
| agegroup3 | |  |  |  | -0.008 |  |
|  |  |  |  |  | [-0.019 0.003] |  |
| female |  |  |  |  | 0.023 |  |
|  |  |  |  |  | [-0.017 0.063] |  |
| partnerinhh== 3.0000 | | |  |  | -0.033 | ** |
|  |  |  |  |  | [-0.055 -0.011] |  |
| bieduc |  |  |  |  | 0.012 | ** |
|  |  |  |  |  | [0.003 0.021] |  |
| empl==employed or self-employed | | | |  | -0.011 |  |
|  |  |  |  |  | [-0.031 0.010] |  |
| empl==non employed | |  |  |  | -0.013 | * |
|  |  |  |  |  | [-0.025 -0.001] |  |
| empl==other | |  |  |  | 0.015 |  |
|  |  |  |  |  | [-0.005 0.035] |  |
| RECODE of hin_adjlog (hin_adjlog) | | | |  | 0.131 |  |
|  |  |  |  |  | [-0.244 0.506] |  |
| housing==NA | |  |  |  | 0.000 |  |
|  |  |  |  |  | [-0.001 0.001] |  |
| Intercept | 0.173 | *** | 0.165 | *** | 0.012 |  |
|  | [0.115 0.231] |  | [0.088 0.242] |  | [-0.385 0.409] |  |
| N | 163007 |  | 163007 |  | 163007 |  |
| *** p<.001, ** p<.01, * p<.05 | | |  |  |  |  |
| Sweden | | |  |  |  |  |
| Adjustment | EUROD-D | | COND, ADL | | SES | |
| group_1 | 2.776 | *** | 2.776 | *** | 2.776 | *** |
|  | [2.749 2.802] |  | [2.749 2.802] |  | [2.749 2.802] |  |
| group_2 | 3.243 | *** | 3.243 | *** | 3.243 | *** |
|  | [3.230 3.256] |  | [3.230 3.256] |  | [3.230 3.256] |  |
| difference | -0.467 | *** | -0.467 | *** | -0.467 | *** |
|  | [-0.497 -0.438] |  | [-0.497 -0.438] |  | [-0.497 -0.438] |  |
| explained | -0.117 | *** | -0.138 | *** | -0.178 | *** |
|  | [-0.129 -0.105] |  | [-0.153 -0.124] |  | [-0.196 -0.160] |  |
| unexplained | -0.350 | *** | -0.329 | *** | -0.289 | *** |
|  | [-0.377 -0.324] |  | [-0.354 -0.305] |  | [-0.314 -0.265] |  |
| EUROD | -0.111 | *** | -0.086 | *** | -0.080 | *** |
|  | [-0.122 -0.099] |  | [-0.095 -0.077] |  | [-0.088 -0.071] |  |
| wave== 4.0000 | -0.006 | *** | -0.006 | *** | -0.005 | *** |
|  | [-0.009 -0.004] |  | [-0.008 -0.004] |  | [-0.006 -0.003] |  |
| wave== 6.0000 | 0.000 |  | 0.000 |  | 0.000 |  |
|  | [-0.000 0.000] |  | [-0.000 0.000] |  | [-0.000 0.000] |  |
| ADL |  |  | -0.013 | *** | -0.011 | *** |
|  |  |  | [-0.017 -0.008] |  | [-0.015 -0.007] |  |
| COND |  |  | -0.034 | *** | -0.030 | *** |
|  |  |  | [-0.041 -0.027] |  | [-0.037 -0.024] |  |
| agegroup1 | |  |  |  | 0.006 | *** |
|  |  |  |  |  | [0.004 0.009] |  |
| agegroup3 | |  |  |  | 0.002 | *** |
|  |  |  |  |  | [0.001 0.004] |  |
| female |  |  |  |  | 0.000 |  |
|  |  |  |  |  | [-0.000 0.001] |  |
| partnerinhh== 3.0000 | | |  |  | 0.001 |  |
|  |  |  |  |  | [-0.000 0.002] |  |
| bieduc |  |  |  |  | -0.035 | *** |
|  |  |  |  |  | [-0.039 -0.030] |  |
| empl==employed or self-employed | | | |  | -0.019 | *** |
|  |  |  |  |  | [-0.022 -0.015] |  |
| empl==non employed | |  |  |  | -0.007 | *** |
|  |  |  |  |  | [-0.009 -0.005] |  |
| empl==other | |  |  |  | 0.005 | ** |
|  |  |  |  |  | [0.002 0.009] |  |
| RECODE of hin_adjlog (hin_adjlog) | | | |  | -0.008 |  |
|  |  |  |  |  | [-0.015 0.000] |  |
| housing==NA | |  |  |  | 0.000 |  |
|  |  |  |  |  | [-0.000 0.000] |  |
| EUROD | 0.080 | *** | 0.076 | *** | 0.065 | *** |
|  | [0.052 0.107] |  | [0.049 0.103] |  | [0.038 0.092] |  |
| wave== 4.0000 | 0.008 |  | 0.013 |  | 0.008 |  |
|  | [-0.012 0.028] |  | [-0.006 0.032] |  | [-0.010 0.026] |  |
| wave== 6.0000 | 0.042 | *** | 0.041 | *** | 0.032 | *** |
|  | [0.023 0.061] |  | [0.024 0.058] |  | [0.016 0.049] |  |
| ADL |  |  | 0.014 | ** | 0.012 | ** |
|  |  |  | [0.005 0.022] |  | [0.004 0.021] |  |
| COND |  |  | 0.032 | * | 0.013 |  |
|  |  |  | [0.000 0.064] |  | [-0.019 0.044] |  |
| agegroup1 | |  |  |  | 0.066 | *** |
|  |  |  |  |  | [0.034 0.098] |  |
| agegroup3 | |  |  |  | 0.014 | * |
|  |  |  |  |  | [0.002 0.025] |  |
| female |  |  |  |  | 0.025 |  |
|  |  |  |  |  | [-0.003 0.053] |  |
| partnerinhh== 3.0000 | | |  |  | -0.015 |  |
|  |  |  |  |  | [-0.038 0.008] |  |
| bieduc |  |  |  |  | -0.001 |  |
|  |  |  |  |  | [-0.020 0.018] |  |
| empl==employed or self-employed | | | |  | -0.026 | * |
|  |  |  |  |  | [-0.050 -0.003] |  |
| empl==non employed | |  |  |  | 0.009 | *** |
|  |  |  |  |  | [0.004 0.013] |  |
| empl==other | |  |  |  | -0.001 |  |
|  |  |  |  |  | [-0.003 0.001] |  |
| RECODE of hin_adjlog (hin_adjlog) | | | |  | -0.954 | *** |
|  |  |  |  |  | [-1.390 -0.517] |  |
| housing==NA | |  |  |  | -0.001 |  |
|  |  |  |  |  | [-0.003 0.002] |  |
| Intercept | -0.480 | *** | -0.505 | *** | 0.464 | * |
|  | [-0.526 -0.434] |  | [-0.558 -0.453] |  | [0.018 0.911] |  |
| N | 159748 |  | 159748 |  | 159748 |  |
| *** p<.001, ** p<.01, * p<.05 | | |  |  |  |  |

| Switzerland | | |  |  |  |  |
| --- | --- | --- | --- | --- | --- | --- |
| Adjustment | EUROD-D | | COND, ADL | | SES | |
| group_1 | 2.722 | *** | 2.722 | *** | 2.722 | *** |
|  | [2.701 2.744] |  | [2.701 2.744] |  | [2.701 2.744] |  |
| group_2 | 3.243 | *** | 3.243 | *** | 3.243 | *** |
|  | [3.230 3.256] |  | [3.230 3.256] |  | [3.230 3.256] |  |
| difference | -0.521 | *** | -0.521 | *** | -0.521 | *** |
|  | [-0.546 -0.496] |  | [-0.546 -0.496] |  | [-0.546 -0.496] |  |
| explained | -0.132 | *** | -0.207 | *** | -0.250 | *** |
|  | [-0.143 -0.121] |  | [-0.221 -0.193] |  | [-0.271 -0.229] |  |
| unexplained | -0.389 | *** | -0.314 | *** | -0.271 | *** |
|  | [-0.412 -0.366] |  | [-0.336 -0.292] |  | [-0.295 -0.247] |  |
| EUROD | -0.131 | *** | -0.102 | *** | -0.095 | *** |
|  | [-0.142 -0.120] |  | [-0.110 -0.093] |  | [-0.103 -0.086] |  |
| wave== 4.0000 | -0.001 |  | -0.001 |  | 0.000 |  |
|  | [-0.002 0.001] |  | [-0.002 0.001] |  | [-0.001 0.001] |  |
| wave== 6.0000 | 0.000 |  | 0.000 |  | 0.000 |  |
|  | [-0.000 0.001] |  | [-0.000 0.000] |  | [-0.000 0.000] |  |
| ADL |  |  | -0.028 | *** | -0.024 | *** |
|  |  |  | [-0.032 -0.023] |  | [-0.028 -0.020] |  |
| COND |  |  | -0.077 | *** | -0.068 | *** |
|  |  |  | [-0.085 -0.070] |  | [-0.075 -0.061] |  |
| agegroup1 | |  |  |  | 0.002 | ** |
|  |  |  |  |  | [0.000 0.003] |  |
| agegroup3 | |  |  |  | 0.001 | * |
|  |  |  |  |  | [0.000 0.002] |  |
| female |  |  |  |  | 0.000 |  |
|  |  |  |  |  | [-0.000 0.001] |  |
| partnerinhh== 3.0000 | | |  |  | 0.000 |  |
|  |  |  |  |  | [-0.001 0.000] |  |
| bieduc |  |  |  |  | -0.022 | *** |
|  |  |  |  |  | [-0.026 -0.019] |  |
| empl==employed or self-employed | | | |  | -0.025 | *** |
|  |  |  |  |  | [-0.029 -0.020] |  |
| empl==non employed | |  |  |  | -0.007 | *** |
|  |  |  |  |  | [-0.009 -0.005] |  |
| empl==other | |  |  |  | 0.002 | ** |
|  |  |  |  |  | [0.001 0.004] |  |
| RECODE of hin_adjlog (hin_adjlog) | | | |  | -0.014 |  |
|  |  |  |  |  | [-0.028 0.000] |  |
| housing==NA | |  |  |  | 0.000 |  |
|  |  |  |  |  | [-0.000 0.000] |  |
| EUROD | -0.002 |  | 0.018 |  | 0.014 |  |
|  | [-0.025 0.021] |  | [-0.006 0.041] |  | [-0.010 0.038] |  |
| wave== 4.0000 | -0.048 | *** | -0.050 | *** | -0.042 | *** |
|  | [-0.066 -0.029] |  | [-0.068 -0.033] |  | [-0.059 -0.025] |  |
| wave== 6.0000 | 0.021 | * | 0.013 |  | 0.012 |  |
|  | [0.003 0.039] |  | [-0.004 0.030] |  | [-0.005 0.028] |  |
| ADL |  |  | 0.003 |  | 0.000 |  |
|  |  |  | [-0.003 0.009] |  | [-0.006 0.007] |  |
| COND |  |  | -0.088 | *** | -0.087 | *** |
|  |  |  | [-0.112 -0.064] |  | [-0.111 -0.063] |  |
| agegroup1 | |  |  |  | 0.010 |  |
|  |  |  |  |  | [-0.018 0.038] |  |
| agegroup3 | |  |  |  | 0.009 |  |
|  |  |  |  |  | [-0.001 0.019] |  |
| female |  |  |  |  | -0.009 |  |
|  |  |  |  |  | [-0.035 0.016] |  |
| partnerinhh== 3.0000 | | |  |  | -0.003 |  |
|  |  |  |  |  | [-0.020 0.015] |  |
| bieduc |  |  |  |  | 0.017 | * |
|  |  |  |  |  | [0.002 0.032] |  |
| empl==employed or self-employed | | | |  | 0.034 | *** |
|  |  |  |  |  | [0.014 0.054] |  |
| empl==non employed | |  |  |  | 0.010 | *** |
|  |  |  |  |  | [0.005 0.014] |  |
| empl==other | |  |  |  | 0.001 |  |
|  |  |  |  |  | [-0.005 0.007] |  |
| RECODE of hin_adjlog (hin_adjlog) | | | |  | -0.541 | *** |
|  |  |  |  |  | [-0.822 -0.260] |  |
| housing==NA | |  |  |  | 0.001 |  |
|  |  |  |  |  | [-0.001 0.004] |  |
| Intercept | -0.361 | *** | -0.209 | *** | 0.302 | * |
|  | [-0.405 -0.318] |  | [-0.256 -0.162] |  | [0.023 0.582] |  |
| N | 158493 |  | 158493 |  | 158493 |  |
| *** p<.001, ** p<.01, * p<.05 | | |  |  |  |  |

#### Supplementary file S.3.2. Explained and unexplained ADL and conditions components

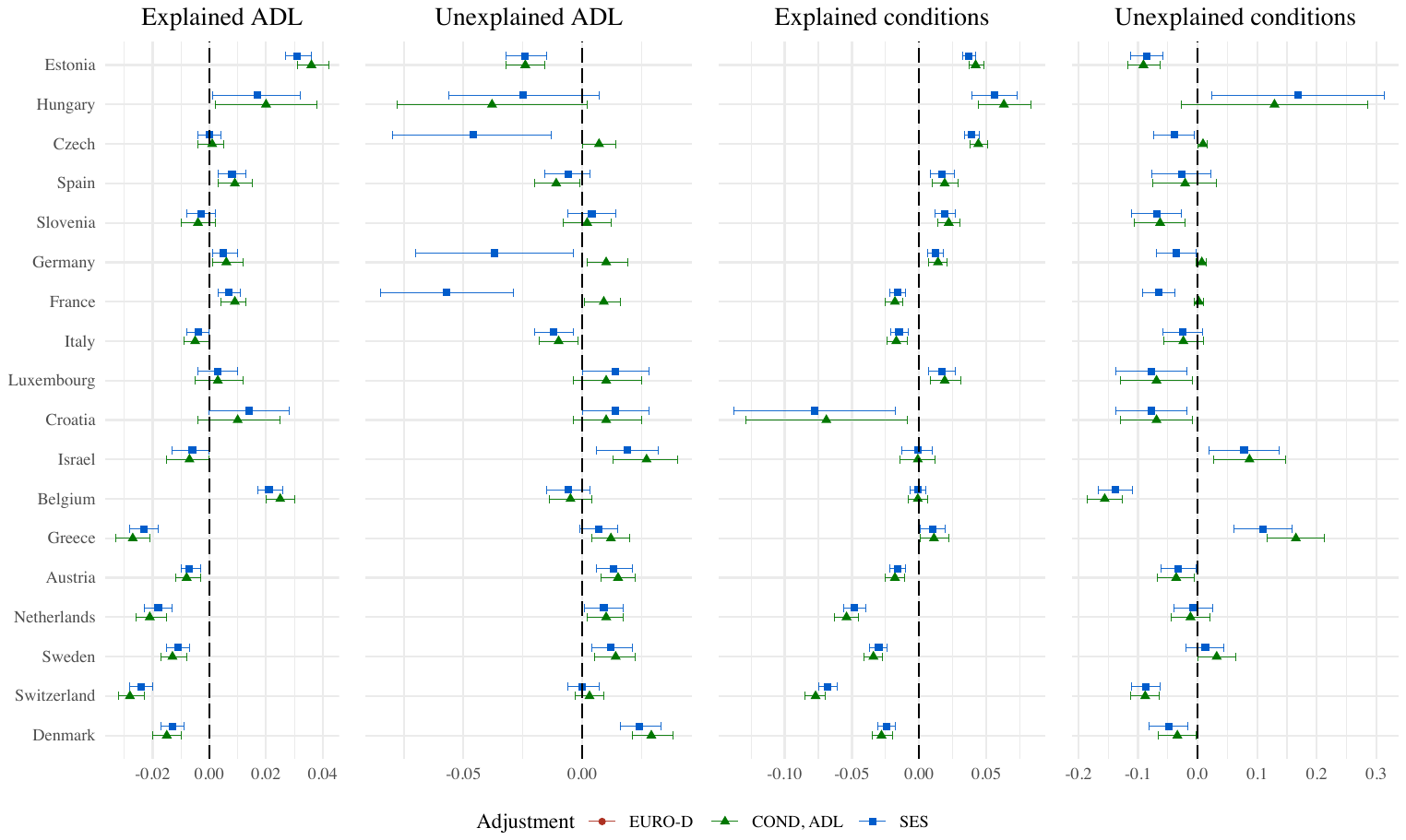

#### Supplementary file S.3.4. Blinder-Oaxaca decomposition by gender, main estimates

#### Main difference and explained and unexplained EURO-D

|  |  | Main difference | | | Explained EURO-D | | | Unexplained EURO-D | | |
| --- | --- | --- | --- | --- | --- | --- | --- | --- | --- | --- |
| Country | Gender | Coef. | 95%CI - | 95%CI + | Coef. | 95%CI - | 95%CI + | Coef. | 95%CI - | 95%CI + |
| Austria | Female | -0.309 | -0.344 | -0.274 | -0.105 | -0.116 | -0.093 | 0.033 | 0.002 | 0.065 |
| Austria | Male | -0.194 | -0.233 | -0.155 | -0.061 | -0.072 | -0.050 | 0.048 | 0.014 | 0.082 |
| Belgium | Female | -0.233 | -0.265 | -0.201 | -0.026 | -0.037 | -0.015 | -0.044 | -0.077 | -0.01 |
| Belgium | Male | -0.221 | -0.256 | -0.186 | -0.001 | -0.011 | 0.01 | 0.008 | -0.027 | 0.044 |
| Croatia | Female | -0.163 | -0.225 | -0.101 | -0.009 | -0.03 | 0.011 | 0.026 | -0.044 | 0.097 |
| Croatia | Male | -0.086 | -0.151 | -0.021 | 0.009 | -0.01 | 0.028 | 0.026 | -0.033 | 0.085 |
| Czech | Female | 0.081 | 0.049 | 0.113 | -0.057 | -0.068 | -0.045 | -0.05 | -0.083 | -0.018 |
| Czech | Male | 0.214 | 0.177 | 0.251 | -0.036 | -0.048 | -0.024 | 0.021 | -0.008 | 0.05 |
| Denmark | Female | -0.707 | -0.747 | -0.666 | -0.145 | -0.158 | -0.131 | 0.077 | 0.04 | 0.114 |
| Denmark | Male | -0.599 | -0.642 | -0.557 | -0.08 | -0.091 | -0.069 | 0.099 | 0.064 | 0.133 |
| Estonia | Female | 0.616 | 0.591 | 0.642 | 0.057 | 0.047 | 0.067 | -0.164 | -0.197 | -0.131 |
| Estonia | Male | 0.752 | 0.723 | 0.78 | 0.101 | 0.09 | 0.112 | -0.127 | -0.159 | -0.095 |
| France | Female | -0.019 | -0.051 | 0.013 | 0.037 | 0.025 | 0.048 | -0.042 | -0.08 | -0.003 |
| France | Male | 0.026 | -0.01 | 0.062 | 0.039 | 0.028 | 0.05 | -0.002 | -0.036 | 0.031 |
| Germany | Female | 0.009 | -0.026 | 0.045 | -0.055 | -0.066 | -0.043 | -0.024 | -0.062 | 0.013 |
| Germany | Male | 0.14 | 0.103 | 0.177 | -0.021 | -0.031 | -0.01 | -0.031 | -0.064 | 0.001 |
| Greece | Female | -0.184 | -0.234 | -0.134 | 0 | -0.019 | 0.02 | -0.152 | -0.202 | -0.102 |
| Greece | Male | -0.295 | -0.352 | -0.239 | 0.015 | -0.004 | 0.033 | -0.177 | -0.226 | -0.128 |
| Hungary | Female | 0.491 | 0.364 | 0.619 | 0.122 | 0.073 | 0.172 | -0.094 | -0.231 | 0.043 |
| Hungary | Male | 0.527 | 0.395 | 0.659 | 0.119 | 0.069 | 0.169 | 0.036 | -0.062 | 0.133 |
| Israel | Female | -0.282 | -0.35 | -0.215 | -0.074 | -0.095 | -0.052 | -0.058 | -0.118 | 0.001 |
| Israel | Male | -0.133 | -0.213 | -0.054 | -0.028 | -0.052 | -0.004 | -0.059 | -0.11 | -0.007 |
| Italy | Female | 0.069 | 0.032 | 0.107 | 0.053 | 0.039 | 0.067 | -0.036 | -0.08 | 0.009 |
| Italy | Male | -0.012 | -0.052 | 0.028 | 0.028 | 0.016 | 0.04 | -0.031 | -0.065 | 0.004 |
| Luxembourg | Female | -0.163 | -0.225 | -0.101 | -0.009 | -0.03 | 0.011 | 0.026 | -0.044 | 0.097 |
| Luxembourg | Male | -0.086 | -0.151 | -0.021 | 0.009 | -0.01 | 0.028 | 0.026 | -0.033 | 0.085 |
| Netherlands | Female | -0.287 | -0.329 | -0.244 | -0.115 | -0.129 | -0.101 | 0.001 | -0.039 | 0.041 |
| Netherlands | Male | -0.243 | -0.29 | -0.195 | -0.077 | -0.09 | -0.064 | 0.016 | -0.018 | 0.051 |
| Slovenia | Female | 0.103 | 0.062 | 0.144 | -0.03 | -0.042 | -0.017 | -0.066 | -0.112 | -0.021 |
| Slovenia | Male | 0.121 | 0.068 | 0.173 | -0.005 | -0.017 | 0.007 | -0.018 | -0.062 | 0.027 |
| Spain | Female | 0.194 | 0.149 | 0.24 | 0.061 | 0.044 | 0.079 | -0.021 | -0.077 | 0.035 |
| Spain | Male | 0.047 | -0.011 | 0.105 | -0.024 | -0.039 | -0.009 | -0.009 | -0.032 | 0.014 |
| Sweden | Female | -0.475 | -0.515 | -0.435 | -0.101 | -0.114 | -0.089 | 0.077 | 0.039 | 0.115 |
| Sweden | Male | -0.458 | -0.5 | -0.415 | -0.053 | -0.064 | -0.042 | 0.048 | 0.01 | 0.087 |
| Switzerland | Female | -0.556 | -0.59 | -0.522 | -0.115 | -0.128 | -0.103 | 0.01 | -0.025 | 0.045 |
| Switzerland | Male | -0.476 | -0.513 | -0.44 | -0.068 | -0.078 | -0.057 | 0.018 | -0.013 | 0.048 |

#### Explained and unexplained ADL and health conditions

|  |  | Explained ADL | | | Explained health conditions | | | Unexplained EURO-D | | | Unexplained health conditions | | |
| --- | --- | --- | --- | --- | --- | --- | --- | --- | --- | --- | --- | --- | --- |
| Country | Gender | Coef. | 95%CI - | 95%CI + | Coef. | 95%CI - | 95%CI + | Coef. | 95%CI - | 95%CI + | Coef. | 95%CI - | 95%CI + |
| Austria | Female | -0.011 | -0.016 | -0.006 | -0.021 | -0.029 | -0.014 | 0.012 | 0.002 | 0.023 | -0.021 | -0.029 | -0.014 |
| Austria | Male | -0.001 | -0.007 | 0.005 | -0.007 | -0.018 | 0.003 | 0.014 | 0.005 | 0.025 | -0.007 | -0.018 | 0.003 |
| Belgium | Female | 0.023 | 0.016 | 0.029 | 0 | -0.008 | 0.007 | 0 | -0.013 | 0.014 | -0.128 | -0.167 | -0.09 |
| Belgium | Male | 0.02 | 0.014 | 0.026 | -0.002 | -0.011 | 0.008 | -0.012 | -0.023 | -0.001 | -0.151 | -0.195 | -0.016 |
| Croatia | Female | 0.006 | -0.005 | 0.017 | 0.004 | -0.009 | 0.017 | 0.011 | -0.011 | 0.033 | -0.041 | -0.12 | 0.038 |
| Croatia | Male | -0.001 | -0.011 | 0.009 | 0.035 | 0.019 | 0.051 | 0.016 | 0 | 0.033 | -0.134 | -0.226 | -0.043 |
| Czech | Female | -0.004 | -0.01 | 0.001 | 0.034 | 0.027 | 0.041 | 0.01 | 0 | 0.02 | -0.026 | -0.071 | 0.019 |
| Czech | Male | 0.006 | 0 | 0.012 | 0.047 | 0.037 | 0.056 | 0.004 | -0.006 | 0.015 | -0.059 | -0.113 | -0.006 |
| Denmark | Female | -0.02 | -0.026 | -0.014 | -0.025 | -0.033 | -0.017 | 0.022 | 0.01 | 0.034 | -0.039 | -0.084 | 0.006 |
| Denmark | Male | -0.004 | -0.01 | 0.002 | -0.023 | -0.034 | -0.013 | 0.026 | 0.014 | 0.038 | -0.06 | -0.106 | -0.013 |
| Estonia | Female | 0.03 | 0.024 | 0.036 | 0.044 | 0.038 | 0.05 | -0.025 | -0.038 | -0.013 | -0.066 | -0.105 | -0.028 |
| Estonia | Male | 0.029 | 0.022 | 0.035 | 0.017 | 0.009 | 0.026 | -0.019 | -0.028 | -0.009 | -0.102 | -0.14 | -0.063 |
| France | Female | 0.005 | -0.001 | 0.01 | -0.016 | -0.023 | -0.009 | 0.005 | -0.006 | 0.016 | -0.073 | -0.109 | -0.036 |
| France | Male | 0.011 | 0.005 | 0.017 | -0.016 | -0.025 | -0.006 | -0.003 | -0.014 | 0.008 | -0.052 | -0.093 | -0.011 |
| Germany | Female | 0 | -0.006 | 0.007 | 0.006 | -0.001 | 0.014 | 0.006 | -0.006 | 0.018 | -0.03 | -0.075 | 0.015 |
| Germany | Male | 0.012 | 0.006 | 0.019 | 0.02 | 0.011 | 0.03 | 0.008 | -0.004 | 0.019 | -0.042 | -0.092 | 0.005 |
| Greece | Female | -0.021 | -0.028 | -0.013 | 0.002 | -0.01 | 0.013 | 0.002 | -0.01 | 0.013 | 0.08 | 0.015 | 0.145 |
| Greece | Male | -0.026 | -0.032 | -0.019 | 0.012 | 0.002 | 0.022 | 0.012 | 0.002 | 0.022 | 0.141 | 0.068 | 0.214 |
| Hungary | Female | 0.013 | -0.008 | 0.034 | 0.065 | 0.046 | 0.084 | -0.014 | -0.056 | 0.027 | 0.243 | 0.024 | 0.463 |
| Hungary | Male | 0.021 | -0.002 | 0.044 | 0.037 | 0.006 | 0.069 | -0.036 | -0.072 | 0 | 0.093 | -0.062 | 0.248 |
| Israel | Female | -0.014 | -0.023 | -0.005 | -0.009 | 0.023 | 0.004 | 0.014 | -0.003 | 0.031 | 0.068 | -0.01 | 0.146 |
| Israel | Male | 0.003 | -0.007 | 0.013 | 0.012 | -0.009 | 0.032 | 0.0243 | 0.004 | 0.043 | 0.093 | 0.003 | 0.183 |
| Italy | Female | 0.001 | -0.005 | 0.007 | -0.015 | -0.023 | -0.007 | -0.009 | -0.021 | 0.004 | -0.049 | -0.094 | -0.005 |
| Italy | Male | -0.009 | -0.014 | -0.005 | -0.014 | -0.024 | -0.004 | -0.015 | -0.025 | -0.006 | 0.001 | -0.048 | 0.05 |
| Luxembourg | Female | 0.006 | -0.005 | 0.017 | 0.004 | -0.009 | 0.017 | 0.011 | -0.011 | 0.033 | -0.041 | -0.12 | 0.038 |
| Luxembourg | Male | -0.001 | -0.011 | 0.009 | 0.035 | 0.019 | 0.051 | 0.016 | 0 | 0.033 | -0.134 | -0.226 | -0.043 |
| Netherlands | Female | -0.019 | -0.026 | -0.012 | -0.042 | -0.052 | -0.032 | 0.019 | 0.008 | 0.03 | -0.015 | -0.06 | 0.029 |
| Netherlands | Male | -0.016 | -0.023 | -0.01 | -0.055 | -0.068 | -0.042 | -0.003 | -0.014 | 0.008 | 0.004 | -0.045 | 0.053 |
| Slovenia | Female | -0.009 | -0.016 | -0.003 | 0.019 | 0.011 | 0.028 | 0.007 | -0.007 | 0.02 | -0.045 | -0.099 | 0.01 |
| Slovenia | Male | 0.004 | -0.004 | 0.012 | 0.019 | 0.006 | 0.031 | 0.001 | -0.015 | 0.016 | -0.093 | -0.156 | -0.029 |
| Spain | Female | 0.017 | 0.01 | 0.025 | 0.018 | 0.009 | 0.028 | -0.006 | -0.022 | 0.009 | -0.031 | -0.093 | 0.032 |
| Spain | Male | -0.004 | -0.01 | 0.002 | 0.015 | -0.001 | 0.031 | -0.006 | -0.017 | 0.004 | -0.027 | -0.104 | 0.05 |
| Sweden | Female | -0.016 | -0.022 | -0.01 | -0.033 | -0.041 | -0.024 | 0.019 | 0.007 | 0.03 | 0.024 | -0.02 | 0.067 |
| Sweden | Male | -0.004 | -0.01 | 0.001 | -0.026 | -0.036 | -0.015 | 0.004 | -0.007 | 0.016 | -0.002 | -0.048 | 0.044 |
| Switzerland | Female | -0.027 | -0.033 | -0.021 | -0.08 | -0.089 | -0.07 | 0.005 | -0.004 | 0.015 | -0.082 | -0.113 | -0.051 |
| Switzerland | Male | -0.019 | -0.025 | -0.014 | -0.051 | -0.061 | -0.04 | -0.006 | -0.013 | 0.001 | -0.092 | -0.13 | -0.054 |

#### Supplementary file S.3.5. Explained and unexplained ADL and health condition components, stratification by gender

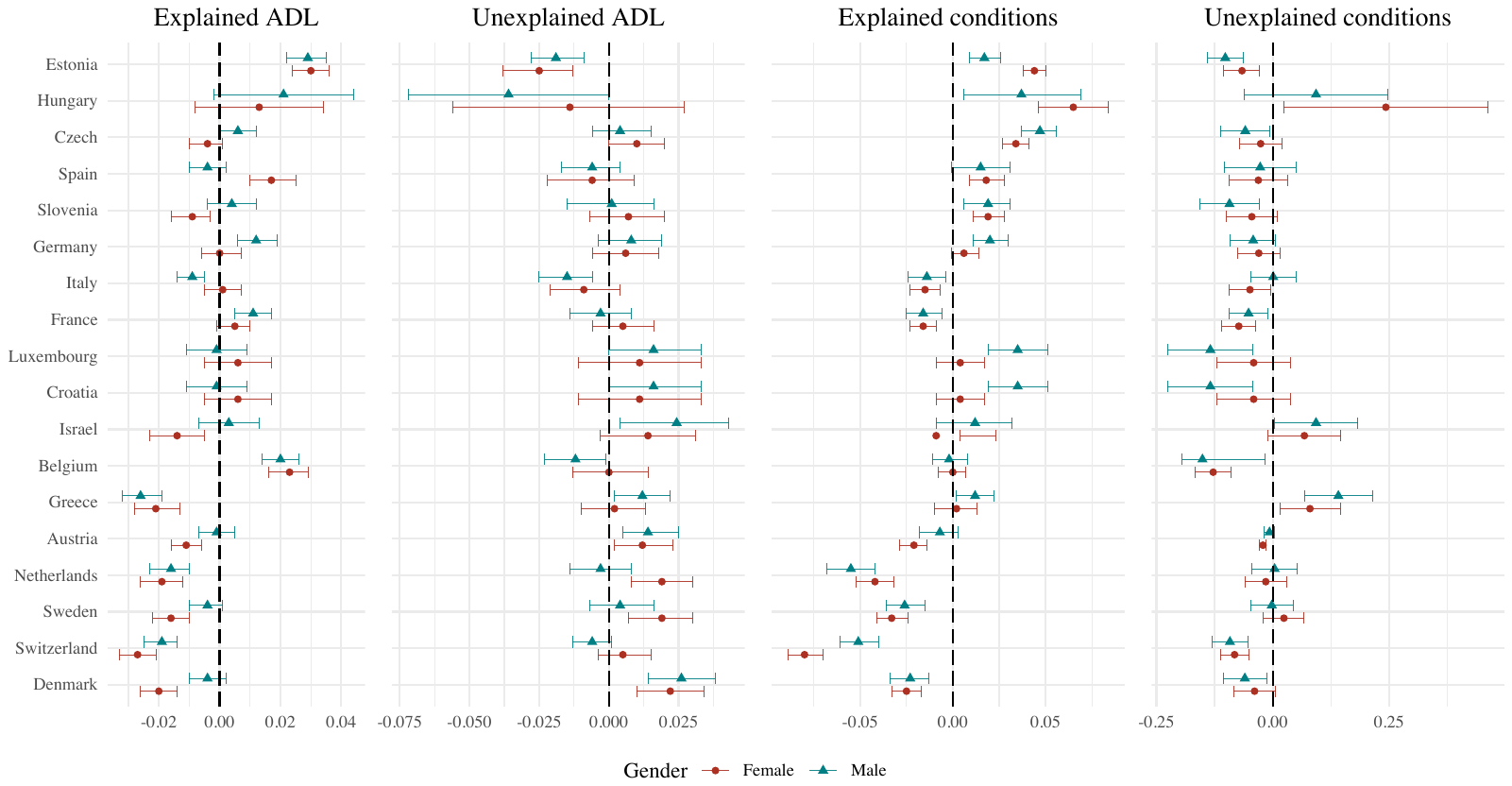

#### Supplementary file S.3.5. Comparing pooled and unique ID models

|  |  | **Main difference** | | | **Explained EURO-D** | | | **Unexplained EURO-D** | | |
| --- | --- | --- | --- | --- | --- | --- | --- | --- | --- | --- |
| Country | Model | Coef. | 95%CI - | 95%CI + | Coef. | 95%CI - | 95%CI + | Coef. | 95%CI - | 95%CI + |
| Austria | Pooled | -0.256 | -0.282 | -0.230 | -0.126 | -0.143 | -0.110 | -0.130 | -0.151 | -0.109 |
| Belgium | Pooled | -0.229 | -0.252 | -0.206 | -0.203 | -0.223 | -0.183 | -0.018 | -0.042 | 0.007 |
| Croatia | Pooled | -0.129 | -0.174 | -0.085 | -0.003 | -0.017 | 0.012 | 0.030 | -0.017 | 0.077 |
| Czech | Pooled | 0.141 | 0.117 | 0.165 | -0.044 | -0.052 | -0.035 | -0.021 | -0.043 | 0.002 |
| Denmark | Pooled | -0.659 | -0.688 | -0.629 | -0.116 | -0.125 | -0.107 | 0.087 | 0.062 | 0.113 |
| Estonia | Pooled | 0.679 | 0.660 | 0.698 | 0.087 | 0.079 | 0.095 | -0.151 | -0.175 | -0.128 |
| France | Pooled | 0.056 | 0.045 | 0.068 | 0.039 | 0.031 | 0.047 | -0.025 | -0.051 | 0.002 |
| Germany | Pooled | 0.067 | 0.042 | 0.093 | -0.042 | -0.050 | -0.034 | -0.027 | -0.053 | -0.002 |
| Greece | Pooled | -0.239 | -0.277 | -0.200 | 0.004 | -0.009 | 0.018 | -0.165 | -0.200 | -0.129 |
| Hungary | Pooled | 0.513 | 0.417 | 0.609 | 0.128 | 0.091 | 0.165 | -0.023 | -0.117 | 0.072 |
| Israel | Pooled | -0.216 | -0.267 | -0.164 | -0.052 | -0.068 | -0.036 | -0.058 | -0.099 | -0.018 |
| Italy | Pooled | 0.030 | 0.002 | 0.058 | 0.038 | 0.030 | 0.049 | -0.035 | -0.064 | -0.006 |
| Luxembourg | Pooled | -0.129 | -0.174 | -0.085 | -0.003 | -0.017 | 0.012 | 0.030 | -0.017 | 0.077 |
| Netherlands | Pooled | -0.267 | -0.298 | -0.235 | -0.098 | -0.108 | -0.088 | 0.009 | -0.018 | 0.036 |
| Slovenia | Pooled | 0.117 | 0.083 | 0.150 | -0.014 | -0.023 | -0.005 | -0.046 | -0.079 | -0.014 |
| Spain | Pooled | 0.129 | 0.092 | 0.166 | 0.024 | 0.011 | 0.036 | -0.006 | -0.050 | 0.029 |
| Sweden | Pooled | -0.467 | -0.497 | -0.438 | -0.080 | -0.088 | -0.071 | 0.065 | 0.038 | 0.092 |
| Switzerland | Pooled | -0.521 | -0.546 | -0.496 | -0.095 | -0.103 | -0.086 | 0.014 | -0.010 | 0.038 |
| Austria | Unique | -0.229 | -0.267 | -0.190 | -0.072 | -0.084 | -0.061 | 0.043 | 0.007 | 0.080 |
| Belgium | Unique | -0.225 | -0.259 | -0.192 | -0.011 | -0.022 | 0.000 | 0.005 | -0.026 | 0.037 |
| Croatia | Unique | -0.144 | -0.200 | -0.089 | -0.007 | -0.025 | 0.010 | 0.031 | -0.028 | 0.089 |
| Czech | Unique | 0.175 | 0.140 | 0.210 | -0.039 | -0.052 | -0.027 | 0.001 | -0.031 | 0.033 |
| Denmark | Unique | -0.612 | -0.654 | -0.570 | -0.109 | -0.121 | -0.098 | 0.117 | 0.079 | 0.154 |
| Estonia | Unique | 0.708 | 0.681 | 0.735 | 0.089 | 0.078 | 0.100 | -0.121 | -0.154 | -0.089 |
| France | Unique | 0.018 | -0.016 | 0.052 | 0.040 | 0.029 | 0.051 | -0.003 | -0.039 | 0.033 |
| Germany | Unique | 0.092 | 0.059 | 0.126 | -0.040 | -0.051 | -0.030 | -0.035 | -0.068 | -0.002 |
| Greece | Unique | -0.243 | -0.283 | -0.204 | 0.003 | -0.011 | 0.017 | -0.162 | -0.199 | -0.125 |
| Hungary | Unique | 0.500 | 0.406 | 0.594 | 0.122 | 0.085 | 0.159 | -0.016 | -0.111 | 0.078 |
| Israel | Unique | -0.232 | -0.297 | -0.167 | -0.039 | -0.061 | -0.018 | -0.060 | -0.113 | -0.008 |
| Italy | Unique | 0.028 | -0.009 | 0.066 | 0.040 | 0.027 | 0.053 | -0.059 | -0.097 | -0.020 |
| Luxembourg | Unique | -0.144 | -0.200 | -0.089 | -0.007 | -0.025 | 0.010 | 0.031 | -0.028 | 0.089 |
| Netherlands | Unique | -0.269 | -0.309 | -0.230 | -0.100 | -0.112 | -0.088 | 0.004 | -0.030 | 0.038 |
| Slovenia | Unique | 0.143 | 0.107 | 0.180 | -0.014 | -0.026 | -0.002 | -0.047 | -0.086 | -0.007 |
| Spain | Unique | 0.049 | -0.012 | 0.110 | 0.008 | -0.011 | 0.028 | 0.020 | -0.034 | 0.074 |
| Sweden | Unique | -0.429 | -0.469 | -0.389 | -0.075 | -0.087 | -0.064 | 0.078 | 0.040 | 0.116 |
| Switzerland | Unique | -0.470 | -0.508 | -0.433 | -0.091 | -0.102 | -0.079 | 0.043 | 0.008 | 0.077 |

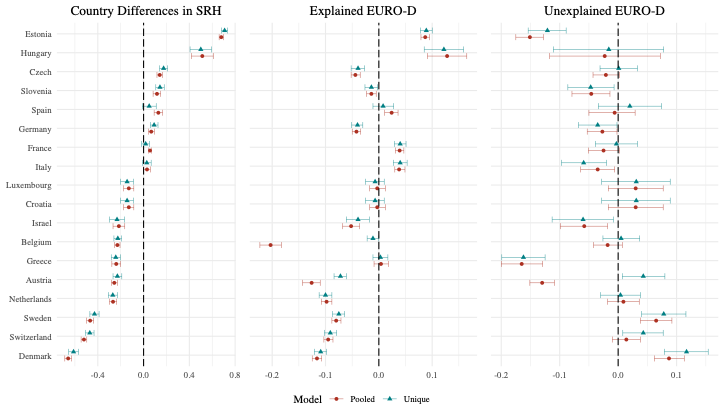

#### Supplementary file S.3.7. Main differences within the linear and interval decomposition models

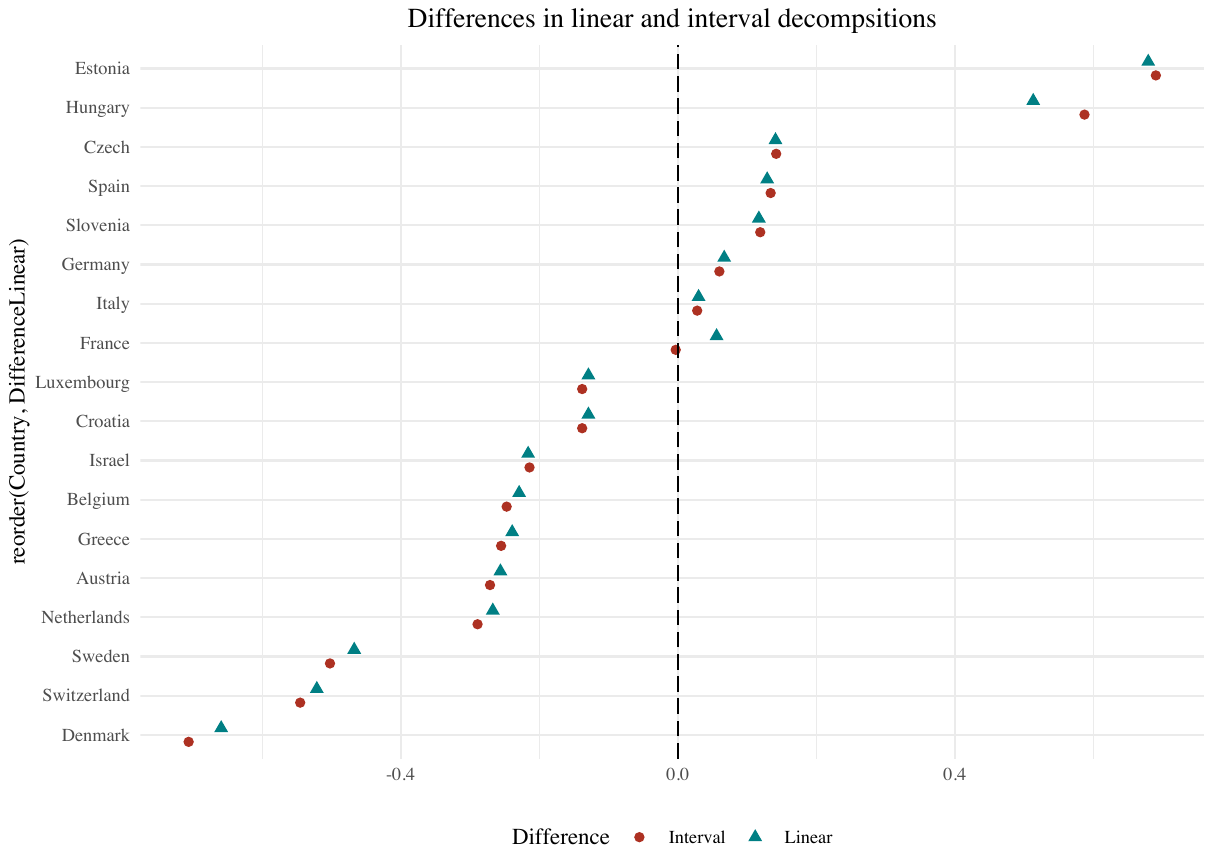
